# Reproducible symptom subtypes of depression identified using unsupervised machine learning

**DOI:** 10.64898/2026.02.13.26346271

**Authors:** David M. Howard, Francisco Diego Rabelo-da-Ponte, Maria Viejo-Romero, Evangelos Vassos, Cathryn M. Lewis

**Affiliations:** Social, Genetic and Developmental Psychiatry Centre, Institute of Psychiatry, Psychology & Neuroscience, King’s College London, London, UK; National Institute for Health Research Maudsley Biomedical Research Centre at South London and Maudsley NHS Foundation Trust and King’s College London, London, UK; Department of Medical & Molecular Genetics, Faculty of Life Sciences and Medicine, King’s College London, London, UK

**Author notes:** Corresponding author: David M. Howard, Social, Genetic and Developmental Psychiatry Centre, Institute of Psychiatry, Psychology & Neuroscience, King’s College London, UK.

## Abstract

Depression is a heterogeneous disorder, often diagnosed based on symptom co-occurrence. However, individuals may present with markedly different symptom profiles, potentially reflecting distinct underlying mechanisms. Identifying common patterns of symptoms using data-driven approaches could help clarify the heterogeneity of depression. Furthermore, examining the sociodemographic and lifestyle characteristics, health status, and polygenic scores of individuals with specific symptom profiles may offer insights into underlying risk factors.

Unsupervised machine learning models were applied to large-scale data from the UK Biobank. Independent groups of individuals were assessed at two time points (the Mental Health Questionnaire: Q1; and the Mental Well-being Questionnaire: Q2) and reporting on historical or current episodes of depression. Two machine learning models, multivariate Bernoulli-mixtures and agglomerative hierarchical clustering, were used to identify common sets of symptoms and cluster individuals by symptom similarity. Consistency of results was examined between Q1 and Q2 and between clustering models. Associations between cluster membership probabilities and sociodemographic and lifestyle factors (sex, age, body mass index, smoking status, ethnicity, and deprivation), eight health conditions, and polygenic scores for bipolar disorder, schizophrenia, and attention-deficit/hyperactivity disorder (ADHD) were examined using regression models.

Symptom clusters were highly consistent across Q1 and Q2 (mean correlation > 0.81) and between machine learning models (Rand Index > 0.83). Clusters aligned with the existing clinical subtypes, atypical and melancholic depression, alongside other potentially novel clusters reflecting a range of different symptom profiles. Atypical clusters (hypersomnia with weight gain) appeared in both Q1 and Q2 and were associated with younger age and higher body mass index. Distinct clusters combining insomnia, weight gain, and having thoughts of death were associated with asthma, suggesting potential inflammatory dysregulation. Further clusters were characterised by psychomotor changes and showed strong associations with Parkinson’s disease, both before and after the mental health questionnaire was conducted.

These findings highlight robust and clinically meaningful symptom subtypes within depression and support the use of data-driven approaches to improve diagnostic refinement and inform personalised treatment strategies.

## Introduction

Depression affects over 320 million people worldwide and is a leading contributor to the global burden of disease (1). Due to its high heterogeneity (2) and the absence of biomarkers, clinical diagnosis is primarily based on symptom presentation as defined in the International Classification of Diseases (11th Revision) (3) or the Diagnostic and Statistical Manual of Mental Disorders-5 (DSM-5) (4). According to DSM-5 criteria, major depressive disorder (MDD) is characterised by a persistent depressed mood and/or loss of interest or pleasure in daily activities, accompanied by at least four additional symptoms, including significant weight change, sleep disturbance (insomnia or hypersomnia), psychomotor changes, fatigue, feelings of worthlessness, difficulty concentrating, and thoughts of death or suicide. These symptoms must co-occur over a two-week period and represent a change from an individual’s usual behaviour and mental state. As a diagnosis requires at least five out of nine symptoms, there are 227 unique symptom combinations that can meet the criteria for MDD (5).

The DSM-5 further defines melancholic and atypical depression as subtypes of the disorder. Melancholic depression is typically associated with reduced appetite or weight loss and insomnia (6, 7), whereas atypical depression involves increased appetite and hypersomnia. Elevated inflammatory markers have been observed in individuals with atypical features, suggesting that immunometabolic dysregulation may influence treatment response (8). Among those with treatment resistant depression (typically defined as a lack of response to two antidepressants taken for sufficient time and dose), symptoms such as insomnia and suicidality are more frequently reported (9). To deliver on the promise of personalised medicine in psychiatry, it is essential to understand how specific symptom profiles relate to the underlying biological heterogeneity of depression to guide more effective and targeted interventions.

Previous studies have had limited success in identifying further depression subtypes. Factor analysis has previously been applied to characterise depressive symptom structures (10–13). However, this approach is limited by its focus on explaining correlations among variables rather than grouping individuals, and its results depend on choices such as rotation method and number of factors (14). Machine learning approaches can overcome some of these limitations and have increasingly been used in depression research (15). However, the focus has primarily been on the classification of case/control status (16–18) or predicting treatment response (19, 20). Although several studies have incorporated depressive symptoms into machine learning analyses (21–23), none has used these methods to identify latent symptom structures within depression. Unsupervised clustering analysis offers the potential to identify symptom patterns without relying on predefined categories, enabling data-driven classification of individuals into groups with similar symptom profiles (24, 25).

The UK Biobank provides a unique opportunity to conduct this research due to its large sample size, extensive phenotyping, genetic data, and repeated mental health assessments (26–28). Depressive symptoms were assessed using both the Composite International Diagnostic Interview – Short Form (capturing worst lifetime episode) and the Patient Health Questionnaire-9 (capturing symptoms over the previous two weeks), enabling evaluation of both lifetime and current depressive symptom patterns at two time points.

In this study, we applied two unsupervised machine learning models to identify clusters of depressive symptoms within the UK Biobank cohort. We then evaluated the consistency of symptom-based classifications across models and across time. Instead of assigning individuals to discrete categories, we used a probability-based grade-of-membership approach to quantify the degree to which each individual aligned with each cluster. Finally, we examined associations between these probabilistic cluster memberships and sociodemographic, lifestyle, clinical, and genetic factors to evaluate the clinical and biological relevance of the identified symptom profiles. This data-driven approach aims to advance our understanding of depression heterogeneity and inform research into potential depressive subtypes.

## Methods

### Study cohort

The UK Biobank is a population-based cohort consisting of over 500,000 individuals (29), accessed under application 82087. Extensive phenotypic, health, lifestyle, and genetic data have been collected for all participants. In addition to the detailed baseline assessments (30), two online questionnaires have collected mental health data. The first mental health questionnaire (Q1) was released in 2017, with responses from 157,366 individuals (26). The second questionnaire, the mental well-being questionnaire (Q2), was released in 2023 with responses from 172,910 individuals (27).

### Depression cases and symptoms

Self-reported depressive symptoms were ascertained from the Composite International Diagnostic Interview – Short Form (CIDI-SF) and the Patient Health Questionnaire (PHQ-9), included in Q1 and Q2. The CIDI-SF assessed symptoms during participants’ worst ever episode of depression, whereas the PHQ-9 assessed symptoms experienced over the last two weeks. The symptom questions used were the same across Q1 and Q2. Only individuals who met criteria for depression using the CIDI-SF or the PHQ-9 and had complete symptom information were taken forward for analysis, with Q1 and Q2 responses assessed separately.

To reduce diagnostic heterogeneity, 4,900 individuals with bipolar disorder or schizophrenia were excluded. These individuals were identified based on self-reported schizophrenia, any other type of psychosis or psychotic illness, mania, hypomania, bipolar or manic-depression (Q1 Data-Field: 20544 / Q2 Data-Field: 29000). Additional bipolar disorder cases were identified from Data-Field: 20126 (31); from reports of mania/excitability (Data-Fields 20501 / 29049) with ≥3 manifestations of manic or irritability (Data-Fields 20548 / 29051); and from reports of extreme irritability (Data-Fields 20502 / 29050) with ≥4 manifestations of mania or irritability (Data-Fields 20548 / 29051) (26).

#### Ever depressed

Individuals were classed as *ever depressed* if they endorsed at least one of two cardinal symptoms and five out of eight depressive symptoms on the CIDI-SF. The two cardinal symptoms were a ‘loss of interest’ (Q1 Data-Field: 20441 / Q2 Data-Field: 29012) and ‘feelings of depression’ (Data-Fields 20446 / 29011). The six additional symptoms included feelings of tiredness (Data-Fields 20449 / 29018), changes in sleep (Data-Fields 20532 / 29021), feelings of inadequacy (Data-Fields 20450 / 29027), difficulty concentrating (Data-Fields 20435 / 29026), weight changes (Data-Fields 20536 / 29021), and thoughts of death (Data-Fields 20437 / 29029). Symptoms had to be present for most or all of the day (Data-Fields 20436 / 29014), and for almost every day or every day (Data-Fields 20439 / 29015), and with an impact on normal roles of either somewhat or a lot (Data-Fields 20440 / 29031).

All symptoms were binarised and were either present or absent. Weight change (Data-Fields 20536 / 29021) was subdivided into the presence or absence of increased weight and the presence or absence of decreased weight. Sleep change was subdivided into the presence or absence of trouble falling asleep (Data-Fields 20533 / 29023), sleeping too much (Data-Fields 20534 / 29025) and waking too early (Data-Fields 20535 and 29024). Symptom prevalences for the *ever depressed* cases is provided in Supplementary Table 1.

#### Currently depressed

Individuals were classed as *currently depressed* if they endorsed at least one of two cardinal symptoms plus five out of nine depressive symptoms on the PHQ-9. The two cardinal symptoms were a ‘lack of interest or pleasure’ (Q1 Data-Field: 20514 / Q2 Data-Field: 29002) and ‘feelings of depression’ (Data-Fields 20510 / 29003) and these symptoms had to be experienced for more than half of the days over the previous two weeks. The remaining seven symptoms were related to over or under sleeping (Data-Fields 20517 / 29004), tiredness or low energy (Data-Fields 20519 / 29005), changes in appetite (Data-Fields 20511 / 29006), feelings of inadequacy (Data-Fields 20507 / 29007), trouble concentrating (Data-Fields 20508 / 29008), psychomotor changes (Data-Fields 20518 / 29009), and suicidal thoughts or self-harm (Data-Fields 20513 / 29010). These seven symptoms were required to be present for more than half of the days over the previous two weeks, except for suicidal thoughts or self-harm, which had to be present for several days over the previous two weeks. All symptoms were coded as either present or absent. Symptom prevalences for the *currently depressed* cases is provided in Supplementary Table 1.

### Groups for analysis

Individuals could meet criteria for *ever depressed* and/or *currently depressed* at either or both the Q1 and Q2 time points. To maximise sample sizes and avoid overlapping samples, independent groups were created (Supplementary Figure 1). Initially, independent groups comprised of 3,096 individuals that were *currently depressed* in Q1, and 3,240 individuals that were *currently depressed* in Q2 were defined. In the remaining participants, individuals who met criteria for *ever depressed* in either Q1 or Q2 were identified. Those who met criteria at both time points were randomly assigned to one of the groups, resulting in 14,864 participants in the Q1 *ever depressed* group and 11,658 in the Q2 group.

### Machine learning models

Two machine learning models were applied to the four groups for analysis.

#### Model 1 - Multivariate Bernoulli mixtures

We applied multivariate Bernoulli-mixtures modelling using the MBM.cluster function from the *comato* package (32) in R version 4.4.0 (33). Probabilistic cluster assignments were obtained using an iterative Expectation-Maximization algorithm. The minimum number of potential clusters was set to 1 and the maximum number of clusters was set to 12 for *currently depressed* and 14 for *ever depressed*. The optimal number of clusters in each group was selected based on the lowest Akaike Information Criterion value. For each cluster, Symptom Scores between 0 and 1 were calculated to represent the probability of symptom presence. Individuals received a set of Probability Scores (summing to 1) reflecting their degree of membership for each cluster. Clusters were named based on their discriminative symptoms. The correlations between the calculated Symptoms Scores and the prevalence of each symptom were calculated.

#### Model 2 - Agglomerative hierarchical clustering

To evaluate the consistency Model 1, we also applied agglomerative hierarchical clustering to the same groups for analysis. The dist function within the built-in *stats* package in R version 4.0.0 (33) was used to create a distance matrix of symptoms using binary distance measures. The distance matrix was then passed to the agnes function within the R *cluster* package (34). The agglomerative coefficient determined the optimum method (average, single, complete, or ward) for clustering. Cluster membership was then assigned with cutree (using the built-in *stats* package in R version 4.0.0), based on the optimum number of clusters identified by the multivariate Bernoulli-mixtures model.

#### Machine learning model comparison

Individuals were assigned to clusters based on their highest Probability Score from Model 1. A statistical comparison of Model 1 assignments with the Model 2 assignments was made using the rand.index function from the R *fossil* package (35). Agreement between models was visualised using Sankey diagrams generated with *dplyr* (36), *ggsankey* (37), and *ggplot2* (38) packages in R version 4.0.0.

### Comparison over time

To assess replicability across time points, correlations between cluster Symptom Scores between Q1 and Q2 were calculated for *ever depressed* and for *currently depressed* groups. A correlation matrix was generated, and the maximum correlation was iteratively selected (without replacement) to identify the best-matched cluster pairs. Q2 clusters were reordered accordingly.

### Regression analyses

Multivariable linear regression models examined associations between multiple factors and individuals’ cluster Probability Scores fitted as the dependent variable. This follows a grade of membership approach (39) rather than discretely assigning individuals to clusters. There were 27 factors (14 sociodemographic and lifestyle factors, 8 health variables, recurrent depression, treatment resistant depression, and 3 polygenic scores) examined as independent variables (Table 1). Regressions were performed using the lm or glm function from the built-in *stats* package in R version 4.0.0 with individuals excluded if they had missing factors or covariates.

**Table 1.** Factor characteristics in *ever depressed* and *currently depressed* groups from Q1 and Q2.

|  | Q1 |  | Q2 |  |
| --- | --- | --- | --- | --- |
|  | *Ever depressed* | *Currently depressed* | *Ever depressed* | *Currently depressed* |
| Sample Size | 14864 | 3096 | 11658 | 3240 |
| % Female | 69.7 | 62.2 | 71.4 | 64.9 |
| Mean Age at Questionnaire (SD) | 62.5 (7.5) | 60.6 (7.6) | 67.3 (7.3) | 66.9 (7.7) |
| Mean Body Mass Index (SD) | 26.9 (4.9) | 28.8 (6.3) | 26.9 (4.8) | 28.3 (5.6) |
| % Current Smoker | 9.2 | 14.1 | 9.1 | 13.4 |
| % Former Smoker | 36.3 | 34.8 | 35.4 | 35.6 |
| % Never Smoked | 54.3 | 51.1 | 55.4 | 50.9 |
| Mean TDI (SD) | -0.07 (0.96) | 0.19 (1.02) | -0.07 (0.96) | 0.16 (1.01) |
| % Asian | 0.7 | 1.5 | 0.5 | 1.2 |
| % Black | 0.6 | 1.0 | 0.7 | 0.9 |
| % Chinese | 0.2 | 0.4 | 0.2 | 0.2 |
| % Mixed | 0.5 | 0.8 | 0.7 | 0.8 |
| % Other | 0.4 | 0.9 | 0.5 | 0.7 |
| % White | 97.5 | 95.4 | 97.0 | 96.2 |
| % UK Place of Birth | 93.4 | 92.1 | 93.8 | 92.8 |
| % Asthma | 16.9 | 19.7 | 17.0 | 21.4 |
| % COPD | 3.8 | 7.5 | 2.8 | 6.0 |
| % Dementia | 0.4 | 1.5 | 0.1 | 0.4 |
| % End Stage Renal Disease | 0.2 | 0.3 | 0.1 | 0.2 |
| % Motor Neurone Disease | 0.1 | 0.3 | 0.003 | 0 |
| % Myocardial Infarction | 3.5 | 5.7 | 2.6 | 4.0 |
| % Parkinson's Disease | 0.5 | 1.4 | 0.2 | 0.6 |
| % Stroke | 2.4 | 3.4 | 1.4 | 3.2 |
| % Recurrent Depression | 56.6 | 86.3 | 58.9 | 74.6 |
| % Treatment Resistant Depression | 5.4 | 15.6 | 5.8 | 13.0 |

SD = standard deviation, TDI = Townsend deprivation index, COPD = chronic obstructive pulmonary disease.

#### Sociodemographic and lifestyle factors

Sex (Data-Field 31), age at questionnaire (Data-Fields 53, 21003, 20400, and 29197), body mass index (Data-Field 21001), smoking status (Data-Field 20116), Townsend Deprivation Index (Data-Field 22189), ethnicity (Data-Field 21000), and place of birth (Data-Field 1647) were fitted simultaneously as independent variables in the regression model. Body mass index (BMI) outliers (top/bottom 2.5% per sex) were set to missing and a reciprocal transformation (1/BMI) applied due to skewness. The transformed BMI values were multiplied by minus one to preserve the interpretability of effect direction. Smoking status was recorded as current smoker, former smoker, or never smoked. Townsend Deprivation Index (TDI) values were increased by 10, log10-transformed, and standardised. Self-reported ethnicity was recorded as either Asian, Black, Chinese, Mixed, Other, or White. Place of birth was a binary variable measuring whether the individual was born in the United Kingdom or not. Sex, ethnicity (White ethnicity as the reference), place of birth, and smoking status (current smoking as the reference) were categorical variables and age at questionnaire, BMI, and TDI were continuous variables.

#### Health variables

Eight algorithmically defined outcomes were used to define the health variables for analysis (https://biobank.ndph.ox.ac.uk/showcase/label.cgi?id=42). The presence of these conditions and the earliest date of diagnosis were derived from self-reported medical condition codes, electronic health records and Death Register records. The health variables were asthma (Data-Field 42014), chronic obstructive pulmonary disease (Data-Field 42016), dementia (Data-Field 42018), end stage renal disease (Data-Field 42046), motor neurone disease (Data-Field 42028), myocardial infarction (Data-Field 42000), Parkinson’s disease (Data-Field 42030), and stroke (Data-Field 42006). All health variables were fitted simultaneously as factors in a regression model with age at questionnaire and sex fitted as covariates.

#### Depression-related factors

A binary variable was derived from Data-Field 20442 (Q1) or 29033 (Q2) as to whether an individual had experienced a single episode of depression or had experienced recurrent episodes. Treatment resistant depression was also assessed as a binary variable following the definition in Fabbri, Hagenaars (40) and was defined as two or more switches between antidepressant drugs, with each drug prescribed for at least six consecutive weeks and the period between the prescription of two consecutive drugs was shorter than 14 weeks. Recurrence and treatment resistance were both fitted as factors and were examined in separate regression models with age at questionnaire and sex fitted as covariates.

#### Polygenic scores

Polygenic scores for other mental health conditions were analysed for an association with individuals’ Probability Scores for each cluster. Genome-wide summary statistics were obtained for Attention Deficit Hyperactivity Disorder (ADHD) (41), bipolar disorder (42), and schizophrenia (43) from studies with no UK Biobank participants. Polygenic scores were calculated using PRScs (44) via GenoPred (45) using a global shrinkage parameter estimated from the data. Polygenic scores were standardised according to the mean and standard deviation using ancestry matched reference data. The mental health polygenic scores for each condition were analysed in a separate model. Ancestry (European, Central/South Asian, African, East Asian, Middle Eastern, or Admixed American) was also included as a covariate in the regression model along with sex, age at questionnaire and the first 10 genetic principal components to account for population stratification (Data-Field 22009).

#### Multiple testing correction

Bonferroni corrections were applied according to the number of factor levels and the number of clusters per group, with α = 0.05. For example, for the Q1 *ever depressed* group (14 clusters), a *P*-value less than 0.05 / (25 × 14) = 1.43 × 10^-4^ was required for a factor to be associated with a cluster. For *currently depressed* in Q2, there were no depression cases diagnosed with motor neurone disease and so only 24 factors were examined for this group. The Bonferroni corrected significance thresholds are reported in Supplementary Table 2.

## Results

### Ever depressed

Using multivariate Bernoulli mixture modelling, 14 clusters were identified in Q1 and 13 clusters in Q2 (Figure 1a and 1b). The ordering of the Q2 clusters was updated based on the correlation with the Q1 Symptom Scores with a mean correlation between the two time points of 0.89, Table 2. Cluster sizes based on individuals’ highest Probability Scores are also provided in Table 2. Regression results for all clusters are provided in Supplementary Tables 3 – 110 with associated factors shown in Figure 1.

**Table 2.** Number of *ever depressed* individuals assigned to each cluster based on the maximum Probability Score calculated using Bernoulli-mixtures. The correlation between the symptom scores between the Mental Health Questionnaire (Q1) and the Mental Well-being Questionnaire (Q2) is provided. These correlations were obtained from a correlation matrix between clusters, where the maximum correlation was extracted iteratively without cluster replacement to identify the most correlated clusters. The Q2 clusters were reordered to reflect the maximum correlation with a Q1 cluster.

| Q1 *Ever depressed* | N (%) | Q2 *Ever depressed* | N (%) | Q1 – Q2 Symptom Score Correlation |
| --- | --- | --- | --- | --- |
| Atypical Symptoms | 1351 (9.1) | Atypical Symptoms | 1156 (9.9) | 0.99 |
| Hypersomnia, Weight Loss, and Mortality-Focus | 469 (3.2) | Hypersomnia, Weight Loss, and Mortality-Focus | 851 (7.3) | 0.91 |
| Hypersomnia and Weight Loss | 558 (3.8) | Weight Loss and Mortality-Focus | 49 (0.4) | 0.47 |
| Prolonged Sleep Latency and Weight Loss | 341 (2.3) | Prolonged Sleep Latency and Weight Loss | 642 (5.5) | 0.92 |
| Terminal Insomnia | 875 (5.9) | Terminal Insomnia | 783 (6.7) | 0.99 |
| Insomnia, Weight Gain and Mortality-Focus | 915 (6.2) | Insomnia, Weight Gain and Mortality-Focus | 957 (8.2) | 0.99 |
| Insomnia and Weight Gain | 1075 (7.2) | Insomnia and Weight Gain | 224 (1.9) | 0.96 |
| Insomnia without Weight Change | 420 (2.8) | Insomnia without Weight Change | 317 (2.7) | 0.99 |
| Insomnia and Low Self-worth | 1438 (9.7) | Melancholia with Low Self-worth and Mortality-Focus | 1945 (16.7) | 0.93 |
| Melancholia with Mortality-Focus | 470 (3.2) | Melancholia | 1854 (15.9) | 0.91 |
| Melancholia with Low Self-worth and Mortality-Focus | 4622 (31.1) | Melancholia with Low Self-worth | 1043 (8.9) | 0.98 |
| Melancholia with Low Self-worth | 303 (2.0) | Insomnia and Low Self-worth | 566 (4.9) | 0.80 |
| Melancholia with Preserved Self-worth and Mortality-Focus | 897 (6.0) | Insomnia and Mortality-Focus | 1271 (10.9) | 0.77 |
| Melancholia with Preserved Self-worth | 1130 (7.6) |  |  | NA |

**Figure 1.**
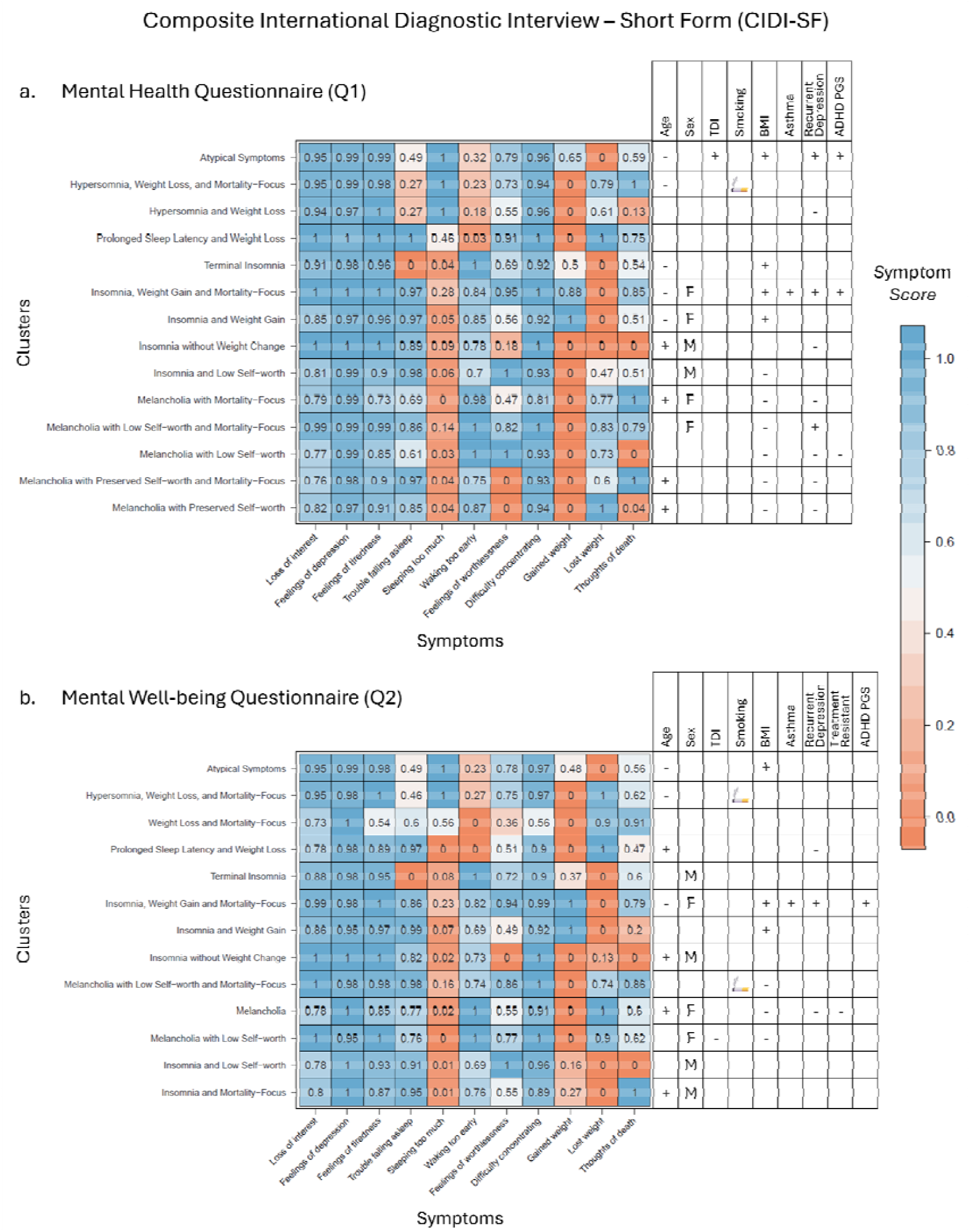
The clusters identified for *ever depressed* cases using the multivariate Bernoulli-mixtures approach and their associated factors. The Symptom Scores are shown for each symptom within each cluster at the Q1 (panel a) and Q2 (panel b) time periods. Higher Symptom Scores (shown in blue) indicate the presence of a symptom, whereas lower Scores (shown in orange) indicate symptoms not endorsed in that cluster. The factors associated with each cluster are shown alongside, with + indicating a positive association, i.e. older age or higher body mass index (BMI) being associated with higher Probability Scores for that cluster. A positive association with Townsend Deprivation Index (TDI) indicates an association with greater deprivation. A positive association for treatment resistance indicates a poorer response to antidepressants. PGS = Polygenic score, ADHD = Attention-deficit/hyperactivity disorder.

Across both time points, changes in sleep and weight, feelings of worthlessness, together with thoughts of death were the principal symptoms distinguishing clusters. Whereas, loss of interest, feelings of depression, feelings of tiredness, and difficulty concentrating showed minimal variation across clusters. A cluster with Atypical Symptoms (hypersomnia and gaining weight) was present in both Q1 and Q2, representing 9-10% of *ever depressed* individuals; these clusters were found in younger individuals with a higher BMI. Smaller clusters characterised by hypersomnia and losing weight were also identified in both waves. Sleep-disturbance clusters reflecting disruptions at either end of the sleep cycle were also found at both time points.

The Insomnia, Weight Gain and Mortality-Focus cluster, identified in both Q1 and Q2, showed a consistent pattern of association with the largest number of external factors. Individuals aligned with this cluster tended to be younger females with a higher BMI, more frequently diagnosed with asthma, greater genetic liability for ADHD, and a history of recurrent depressive episodes. Insomnia also featured across many other clusters, including several characterised by Melancholia (insomnia and losing weight). Five clusters included Melancholia in Q1, collectively comprising 49.9% of *ever depressed* individuals when making discrete assignments of individuals to clusters. The three Melancholia clusters in Q2 accounted for 41.5% of that sample.

Cluster assignments, based on individuals’ highest Probability Scores from the multivariate Bernoulli mixture model, were compared with those produced by agglomerative hierarchical clustering. Agreement between the two models was high. Rand Index values, which measure similarity between two clustering solutions (1 = perfect agreement; 0 = no agreement beyond chance), were 0.87 for Q1 and 0.89 for Q2. Visualisation of the assignments between the two machine learning approaches for *ever depressed* cases is provided in Supplementary Figure 2 (Q1) and Supplementary Figure 3 (Q2).

### Currently depressed

Among *currently depressed* cases, 11 clusters were identified in both Q1 and Q2 using multivariate Bernoulli-mixtures, Figure 2a and 2b. Factors associated with individuals’ Probability Scores for each cluster are also shown in Figure 2, with full regression outputs provided in Supplementary Tables 111 - 198. The ordering of the Q2 clusters was updated based on the correlation with Q1 Symptom Scores, with a mean correlation between the two time points of 0.81 (Table 3). The number of individuals assigned to each cluster based on their maximum Probability Score is also provided in Table 3.

**Figure 2.**
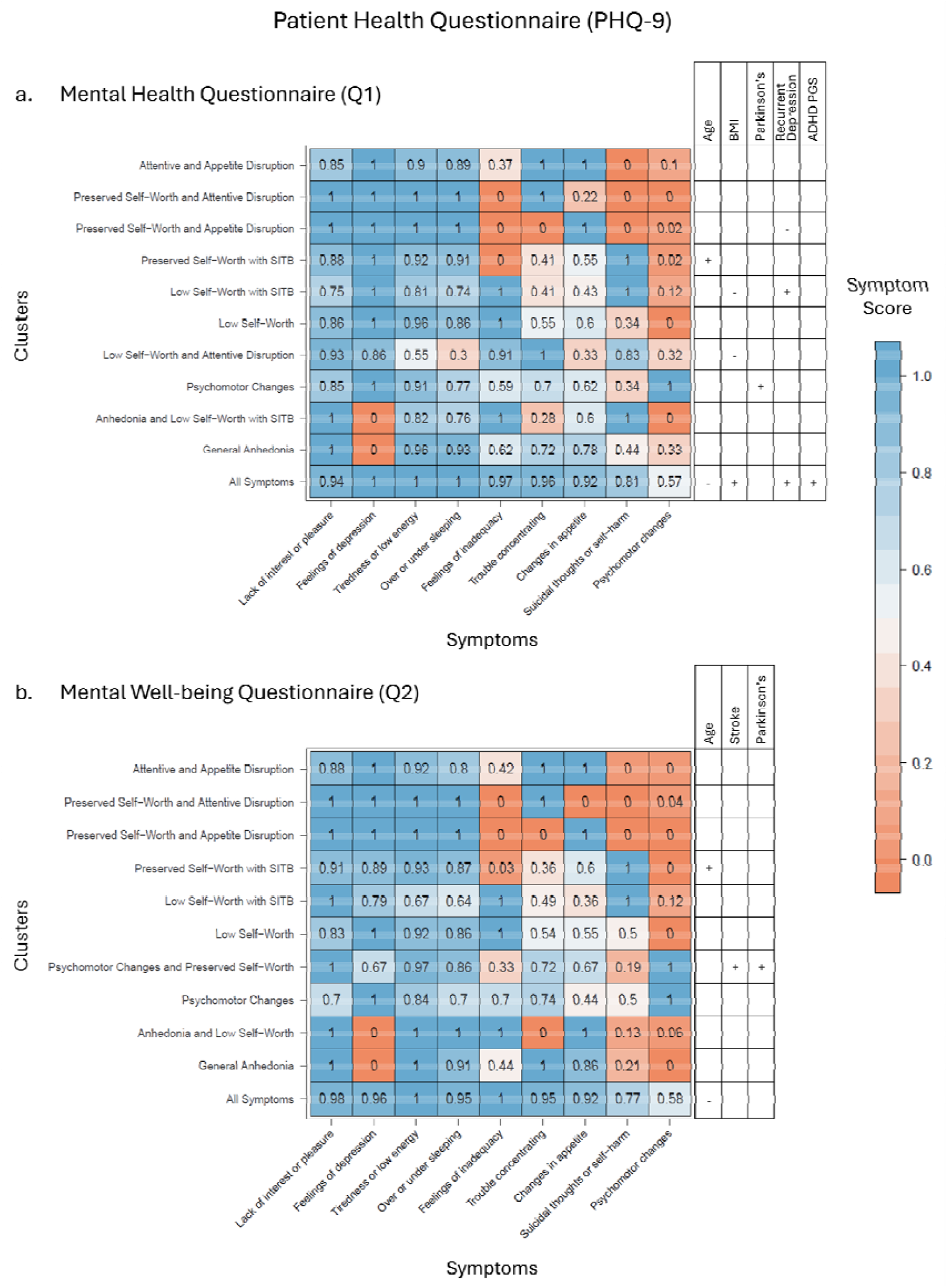
The clusters identified for *currently depressed* cases using the multivariate Bernoulli-mixtures approach and their associated factors. The Symptom Scores are shown for each symptom within each cluster at the Q1 (panel a) and Q2 (panel b) time periods. Higher Symptom Scores (shown in blue) indicate the presence of a symptom, whereas lower Scores (shown in orange) indicate symptoms not endorsed in that cluster. The factors associated with each cluster are shown alongside, with + indicating a positive association, i.e. older age or higher body mass index (BMI) being associated with higher Probability Scores for that cluster. PGS = Polygenic score, ADHD = Attention-deficit/hyperactivity disorder, SITB = Self-Injurious Thoughts and Behaviours.

**Table 3.** Number of *currently depressed* individuals assigned to each cluster based on the maximum Probability Score calculated using Bernoulli-mixtures. The correlation between the symptom scores between the Mental Health Questionnaire (Q1) and the Mental Well-being Questionnaire (Q2) is provided. These correlations were obtained from a correlation matrix between clusters, where the maximum correlation was extracted iteratively without cluster replacement to identify the most correlated clusters. The Q2 clusters were reordered to reflect the maximum correlation with a Q1 cluster.

| Q1 *Currently depressed* | N (%) | Q2 *Currently depressed* | N (%) | Q1 – Q2 Symptom Score Correlation |
| --- | --- | --- | --- | --- |
| Attentive and Appetite Disruption | 107 (3.5) | Attentive and Appetite Disruption | 175 (5.4) | 0.99 |
| Preserved Self-Worth and Attentive Disruption | 67 (2.2) | Preserved Self-Worth and Attentive Disruption | 100 (3.1) | 0.99 |
| Preserved Self-Worth and Appetite Disruption | 131 (4.2) | Preserved Self-Worth and Appetite Disruption | 159 (4.9) | 1.00 |
| Preserved Self-Worth with SITB | 206 (6.7) | Preserved Self-Worth with SITB | 271 (8.4) | 0.99 |
| Low Self-Worth with SITB | 471 (15.2) | Low Self-Worth with SITB | 301 (9.3) | 0.91 |
| Low Self-Worth | 635 (20.5) | Low Self-Worth | 1053 (32.5) | 0.99 |
| Low Self-Worth and Attentive Disruption | 121 (3.9) | Psychomotor Changes and Preserved Self-Worth | 195 (6.0) | -0.41 |
| Psychomotor Changes | 291 (9.4) | Psychomotor Changes | 151 (4.7) | 0.86 |
| Anhedonia and Low Self-Worth with SITB | 25 (0.8) | Anhedonia and Low Self-Worth | 28 (0.9) | 0.70 |
| General Anhedonia | 338 (10.9) | General Anhedonia | 148 (4.6) | 0.93 |
| All Symptoms | 704 (22.7) | All Symptoms | 659 (20.3) | 0.97 |

Clusters for *currently depressed* individuals were primarily distinguished by the presence or absence of feelings of inadequacy, trouble concentrating, changes in appetite, and suicidal thoughts or self-harm. In both Q1 and Q2, there were two clusters characterised by the cardinal symptom of loss of interest or pleasure (anhedonia) rather than feelings of depression. Additional clusters were also defined by the presence of psychomotor changes. The All Symptoms cluster in Q1 was associated with the most associated factors, including younger age, higher BMI, recurrent episodes of depression, and increased polygenic liability for ADHD. There were not as many associated factors with the *currently depressed* clusters compared to the *ever depressed* clusters, although there were considerable fewer *currently depressed* individuals thereby reducing statistical power.

Comparing cluster assignments from the Bernoulli mixture model with those from agglomerative hierarchical clustering yielded Rand Index values of 0.84 (Q1) and 0.83 (Q2). Sankey diagrams visualising the assignments between the two machine learning models are shown in Supplementary Figure 4 (Q1) and Supplementary Figure 5 (Q2).

### Symptom Scores and Prevalence

Symptoms Scores derived from the multivariate Bernoulli mixture model were highly correlated with observed symptom prevalence based on individuals assigned to their highest-probability cluster (r = 0.99). Symptom Scores therefore accurately reflected the presence or absence of symptoms within clusters.

### Regression Analyses

#### Age

In the *ever depressed* analysis, none of the eight clusters associated with older age across Q1 and Q2 had high Symptom Scores (≥ 0.60) for feelings of worthlessness. In contrast, seven out of the eight clusters associated with younger age showed high Symptom Scores for worthlessness. A similar pattern was observed for feelings of inadequacy in the *currently depressed* analysis.

Age-related patterns also emerged for weight and sleep symptoms. Younger-age clusters were more likely to endorse weight gain, with six out of eight clusters associated with younger age having moderate Symptom Scores (≥0.40) for gaining weight. Conversely, seven of eight older-age associated clusters had a Symptom Score of zero for gaining weight. Sleeping too much was also rare among older-age clusters, with Symptom Scores ≤0.20 in all eight clusters. These combined patterns relate atypical symptomatology (hypersomnia and weight gain) with younger age and melancholic features (insomnia and weight loss) with older age.

#### Sex and Body Mass Index

Across the *ever depressed* analyses, only one of the six male-associated clusters had a moderate Symptom Score (≥ 0.40) for either weight gain or weight loss. In contrast, all seven female-associated clusters had very high Symptom Scores (≥ 0.80) for weight change in either direction. BMI was also strongly aligned with symptom patterns. All seven clusters associated with higher BMI had a Symptom Score of zero for losing weight and at least moderate Symptom Scores (≥ 0.40) for gaining weight. In contrast, the nine clusters associated with lower BMI had a zero Symptom Score for gaining weight and at least a moderate Symptom Score (≥ 0.40) for losing weight. All nine of these low BMI-associated clusters also had very low Symptom Scores (≤ 0.20) for sleeping too much. These findings suggest sex-specific patterns in somatic depressive symptomatology, which are also associated with an individual’s BMI.

#### Recurrent Depression

In the *ever depressed* analysis, all four clusters associated with recurrent depression across Q1 and Q2 had at least moderate Symptom Scores (≥ 0.40) for thoughts of death. Three of these four clusters also had zero Symptom Scores for losing weight. Whereas all eight clusters associated with single episode depression had zero Symptom Scores for gaining weight.

#### Parkinson’s Disease

In the *currently depressed* analysis, Parkinson’s disease was associated with two clusters (Q1: *P* = 1.00 × 10^-6^ and Q2: *P* = 3.51 × 10^-11^), both of which had Symptom Scores of one for psychomotor changes. A diagnosis of Parkinson’s disease prior to the mental health questionnaire (*P* = 1.84 × 10^-4^) and after the questionnaire (*P* = 0.0024) were both associated with individuals’ Probability Scores for the Psychomotor Changes cluster in Q1. After the well-being questionnaire (Q2), there were no individuals diagnosed with Parkinson’s disease so timing of diagnosis could not be examined.

## Discussion

This study applied unsupervised machine learning to identify symptom-based clusters of depression in the large UK Biobank population cohort. By modelling symptom patterns without imposing predefined diagnostic categories, coherent and replicable depressive subtypes were characterised. The UK Biobank contains symptom information collected at two time points, the mental health questionnaire (Q1; 2017) and the mental well-being questionnaire (Q2; 2022). The clusters identified between these two questionnaires were highly similar in terms of the sets of symptoms that were endorsed (mean correlation ≥ 0.81). Comparisons of the results from the two different clustering algorithms were also highly concordant (Rand Index ≥ 0.83) further supporting the replicability of the results.

In the *ever depressed* analysis, somatic symptoms (particularly the direction of weight change and the presence of insomnia or hypersomnia) were important drivers for cluster differentiation. Clusters aligned closely with existing DSM-5 criteria for melancholic and atypical subtypes of depression, supporting the validity of the machine learning approach and providing empirical evidence that these subtypes emerge naturally from data-driven methods. In both Q1 and Q2, clusters displaying Atypical Symptoms (oversleeping and gaining weight) were found, although their prevalence (9-10%) was lower than that generally reported (15-29%) (46, 47). Clusters featuring the less commonly described combination of hypersomnia with weight loss also appeared consistently and showed associations with younger age and smoking. Although evidence for causal links between depression and smoking are inconsistent (48), this subtype may reflect a distinct mood–somatic profile that warrants further investigation.

Melancholic clusters (insomnia and losing weight) were the most prevalent, accounting for 41.5 - 49.9% of *ever depressed* individuals. These clusters were generally associated with older age, female sex, and lower BMI. Estimates of the prevalence of melancholic depression amongst those with disorder vary widely from 7-70% (49) with the type of cohort analysed (inpatient vs. outpatient) identified as a key factor in prevalence variability (50). However, the sample sizes (n = 14,864 and 11,658) and the replicable nature of the current analysis increase confidence in the prevalences reported here for a population-based cohort. Across both questionnaires, insomnia frequently appeared in combination with other symptoms. Notably, the Insomnia, Weight-Gain and Mortality-Focus clusters had the greatest number of associated factors in the regression analysis, including asthma. Substantial research has examined the role of inflammatory dysregulation and depression (51), with elevated levels of inflammatory markers, including C-reactive protein (52), interleukin-6, and tumor necrosis factor-α (53), observed during depressive episodes. Autoimmune diseases and infections have also been identified as risk factors for subsequent depression diagnoses (54, 55). Asthma is a common inflammatory disease affecting the lungs and is comorbid with depression (56–58). Evidence also indicates that females may experience worse exacerbations or poorer outcomes (59, 60) which aligns with the demographic profile of these clusters.

Across both questionnaires, Parkinson’s disease was associated with clusters with very high Symptom Scores for psychomotor changes. This symptom of depression was assessed by participants reporting slowed moving or speaking or that they were fidgety or restless, which are key features of Parkinson’s disease. The prevalence of depression among those with Parkinson’s is estimated to be 38%, more than twice that of the general population (61). This increased depression prevalence has been attributed to fears around the disease and the impact of living with the condition (62, 63). It has also been attributed to changes in neurotransmission, including the noradrenaline, monoaminergic, and dopamine systems, which are known to be affected in Parkinson’s and have roles in mood regulation (64, 65). In the current study, the diagnosis of Parkinson’s disease also occurred after completing the Mental Health Questionnaire, raising the possibility that prominent psychomotor symptoms during a depressive episode could precede or signal emerging neurological disease. Therefore, clinicians should consider whether an individual endorsing the psychomotor changes item on the Patient Health Questionnaire may warrant further neurological evaluation, particularly in older adults.

Despite compelling evidence of the replication of clusters across different time points and machine learning models, the sample was drawn from a single cohort, the UK Biobank. The breadth of phenotyping and size of the UK Biobank make it a valuable resource; however, it is unlikely to be representative of the global population and compared to the general UK population it has been shown to be healthier and wealthier (66). The youngest participant in the current study was 46 at the point the Mental Health Questionnaire was completed and therefore the results presented do not capture the symptoms experienced by those in early adulthood. Therefore, validation of the current results should be sought in study cohorts from different demographic and geographical backgrounds. The current study examined self-reported depressive symptoms and examined historical episodes of depression (for the *ever depressed* analyses) and is therefore likely to be subject to recall bias (67). Additionally, it has been shown that there is only a moderate degree of correlation between self-reported and clinically ascertained symptoms (68). This highlights the need for further exploration of symptom clusters using clinician-rated measures.

## Conclusion

This study demonstrates that unsupervised machine learning can uncover clinically meaningful and replicable symptom-based subtypes of depression. These findings highlight the substantial heterogeneity within current diagnostic criteria and support the need for more personalised diagnostic and treatment approaches. While the DSM-5 provides a validated framework for identifying depressive symptoms, it may group multiple distinct subtypes under a single diagnosis. The existing melancholic and atypical depression subtypes were validated, but other latent subtypes likely remain uncharacterised. As research continues to advance towards personalised mental health care, the insights from this study offer a valuable contribution to our understanding of depression’s complexity.

## Acknowledgements

This research was conducted using the UK Biobank resource, application number 82087. We are grateful to the UK Biobank and all its voluntary participants. The UK Biobank study was conducted under generic approval from the NHS National Research Ethics Service (approval letter dated June 17, 2011, Ref 11/NW/0382). All participants gave full informed written consent.

D.M.H, and F.D.R-d-P are supported by an MRC Career Development Award (Reference MR/Y011112/1). M.V-R is in receipt of a PhD studentship funded by the National Institute for Health and Care Research (NIHR) Maudsley Biomedical Research Centre (BRC). C.M.L acknowledges MRC grant MR/N015746/1. This investigation represents independent research part-funded by the National Institute for Health Research (NIHR) Maudsley Biomedical Research Centre at South London and Maudsley NHS Foundation Trust and King’s College London. The views expressed are those of the authors and not necessarily those of the NHS, the NIHR or the Department of Health and Social Care.

For the purpose of open access, the author has applied a CC BY public copyright licence to any Author Accepted Manuscript version arising from this submission.

## Data availability

Available from UK Biobank subject to standard procedures (www.ukbiobank.ac.uk). The R code used to conduct this research is available from: https://github.com/davemhoward/MLdepressionclustering

