## Supplementary Figures for "Reproducible symptom subtypes of depression identified using unsupervised machine learning"

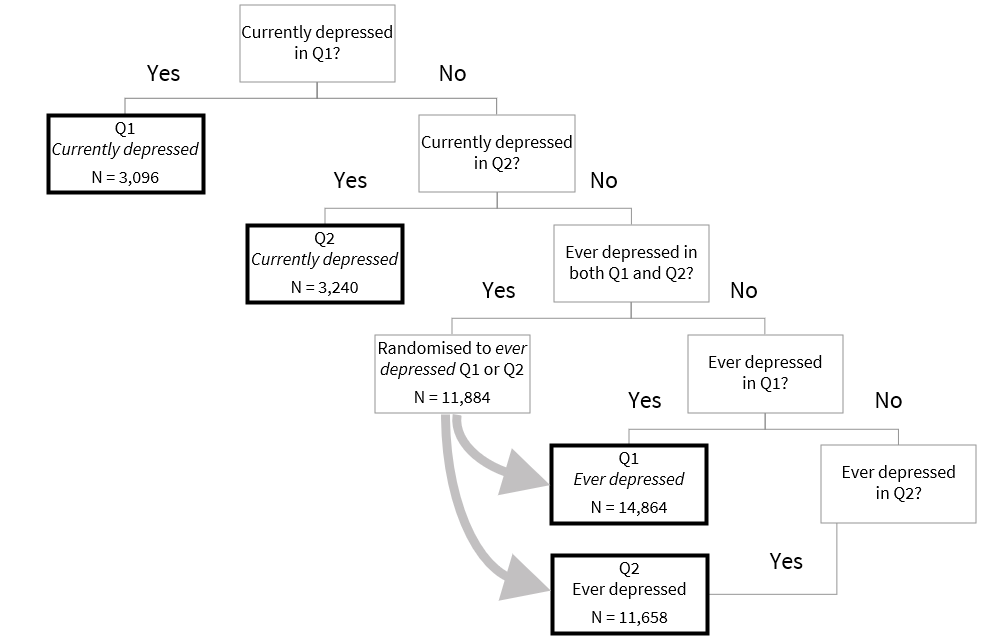

**Supplementary Figure 1**. Flowchart illustrating the classification process for allocating individuals with depression into the four analysis groups.

The four groups for analysis are highlighted by thicker bordered boxes. The number of individuals (N) with complete symptom information in each group is provided. Q1 = Mental Health Questionnaire. Q2 = Well-being Questionnaire.

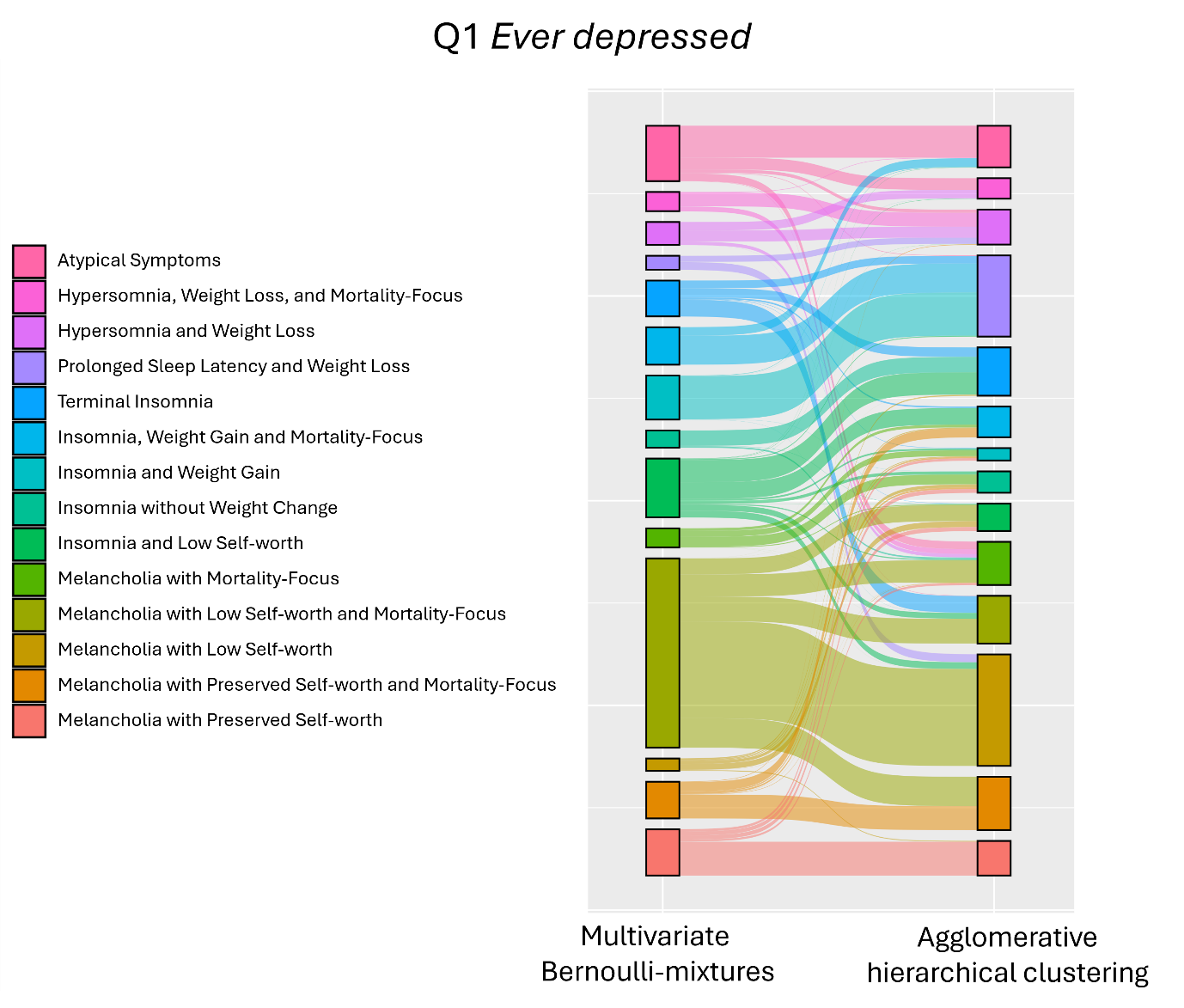

**Supplementary Figure 2**. Sankey plot illustrating the assignment of *ever depressed* individuals to clusters using multivariate Bernoulli-mixtures and agglomerative hierarchical clustering based on symptoms reported in the Mental Health Questionnaire (Q1).

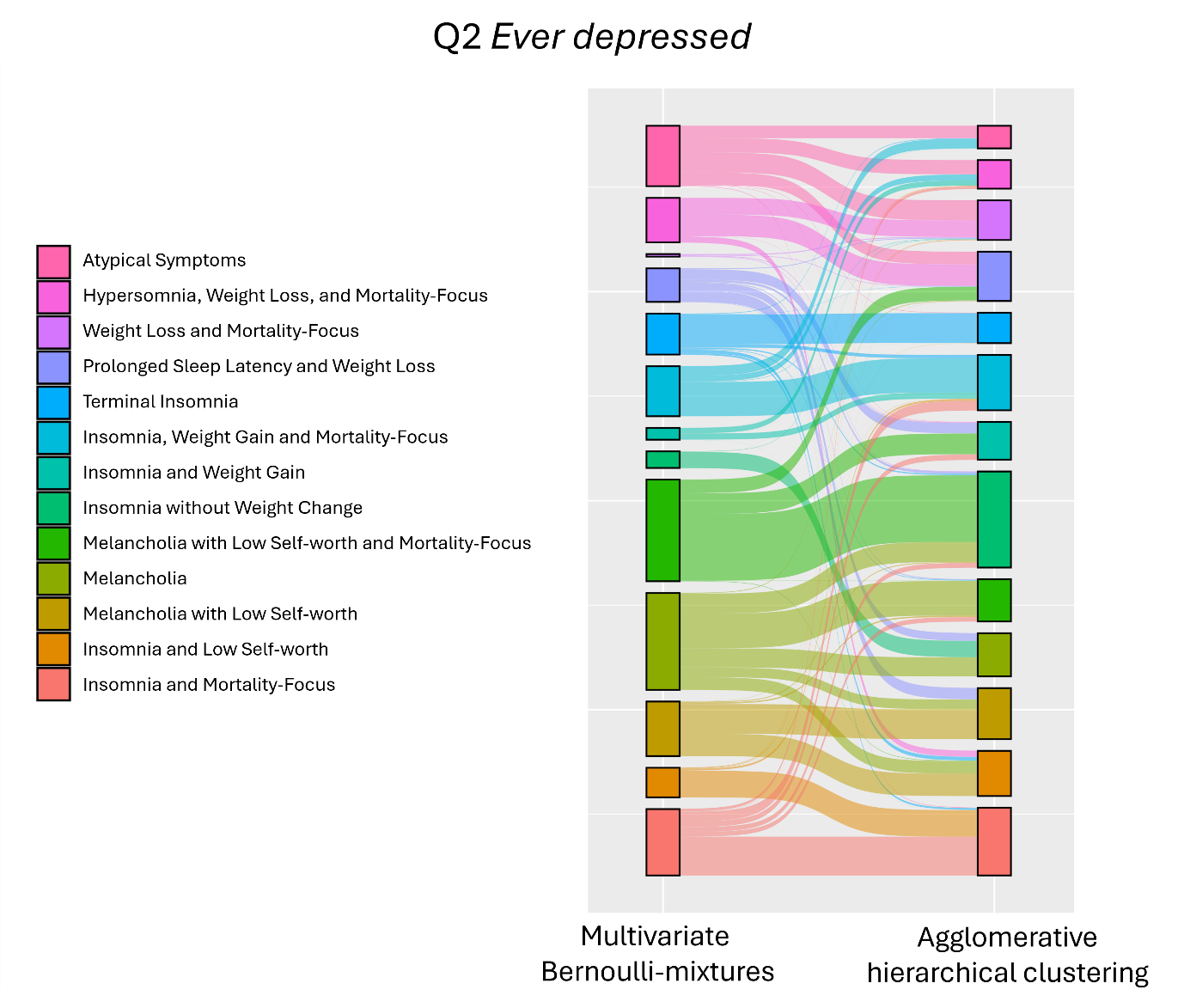

**Supplementary Figure 3**. Sankey plot illustrating the assignment *ever depressed* individuals to clusters using multivariate Bernoulli-mixtures and agglomerative hierarchical clustering based on symptoms reported in the Mental Well-being Questionnaire (Q2).

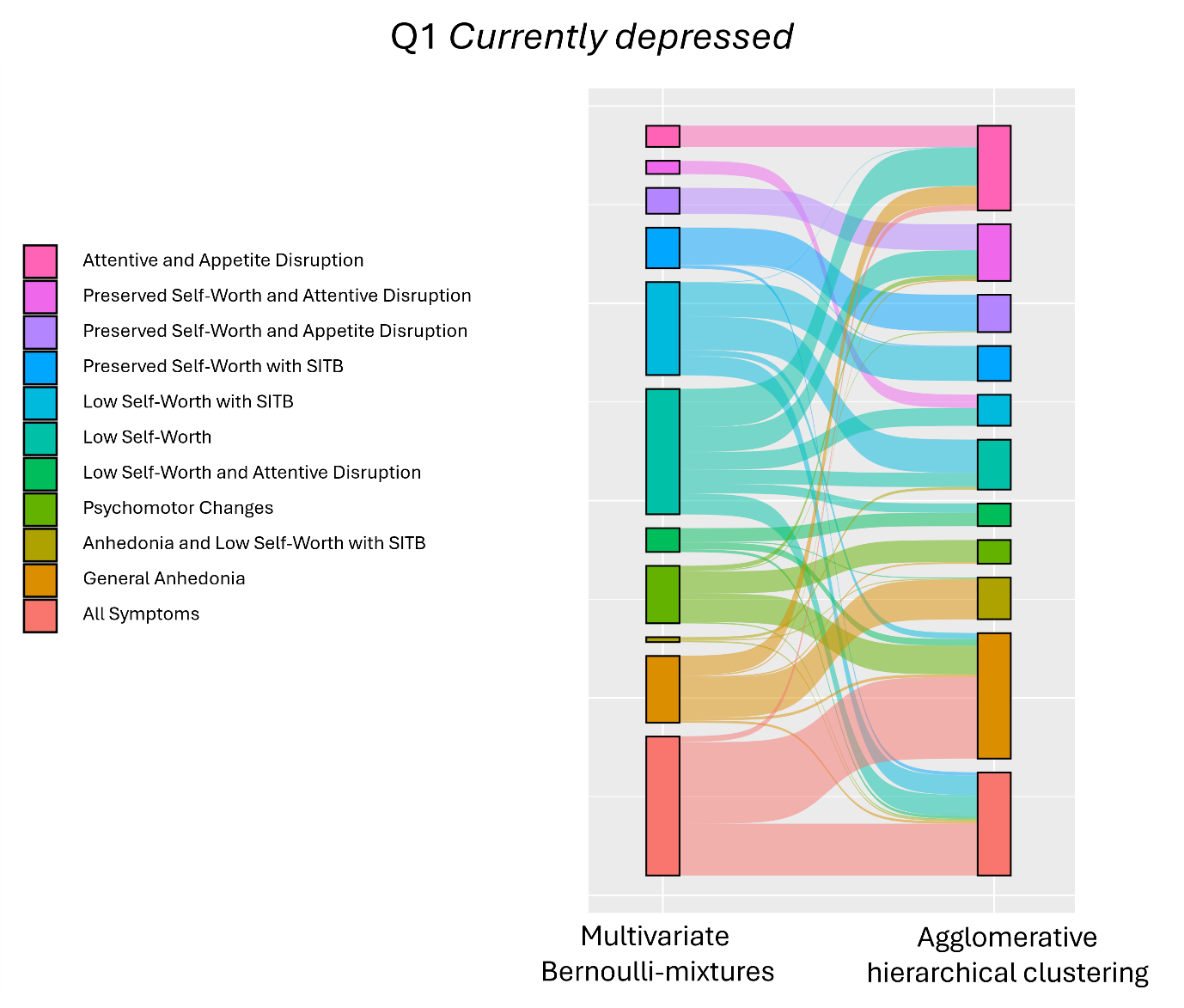

**Supplementary Figure 4**. Sankey plot illustrating the assignment of *currently depressed* individuals to clusters using multivariate Bernoulli-mixtures and agglomerative hierarchical clustering based on symptoms reported in the mental health questionnaire (Q1).

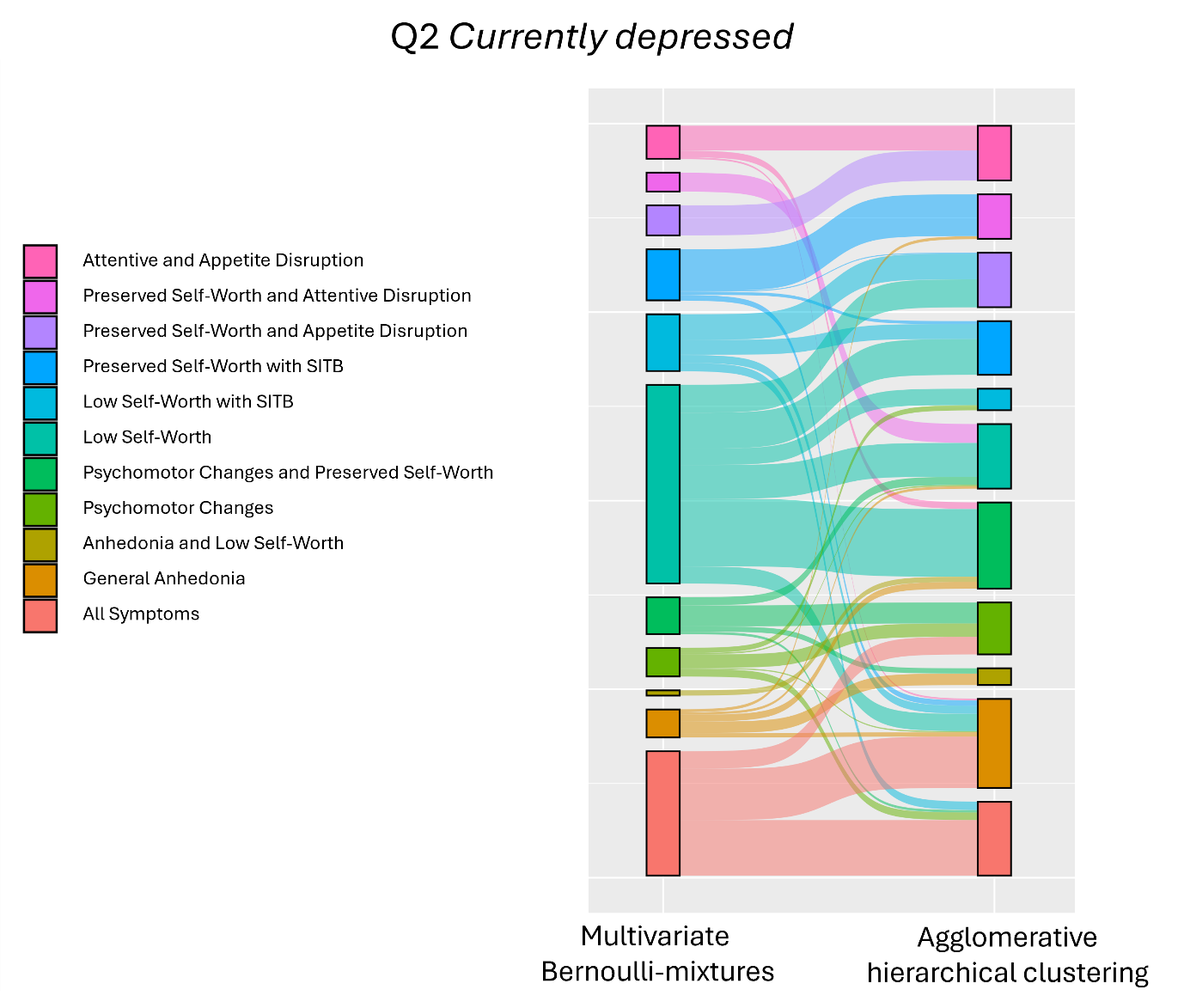

**Supplementary Figure 5**. Sankey plot illustrating the assignment of *currently depressed* individuals to clusters using multivariate Bernoulli-mixtures and agglomerative hierarchical clustering based on symptoms reported in the mental well-being questionnaire (Q2).
