## Supplementary Tables for "Reproducible symptom subtypes of depression identified using unsupervised machine learning"

Supplementary Table 1. Prevalence of symptoms among *ever depressed* and *currently depressed* individuals within the Mental Health Questionnaire (Q1) and the Mental Well-being Questionnaire (Q2).

|  | Symptom | Q1 | Q2 |
| --- | --- | --- | --- |
| *Ever depressed* | Loss of interest | 0.897 | 0.896 |
|  | Feelings of depression | 0.985 | 0.984 |
|  | Feelings of tiredness | 0.943 | 0.936 |
|  | Trouble falling asleep | 0.776 | 0.757 |
|  | Sleeping too much | 0.231 | 0.216 |
|  | Waking too early | 0.774 | 0.713 |
|  | Feelings of worthlessness | 0.653 | 0.694 |
|  | Difficulty concentrating | 0.951 | 0.948 |
|  | Gained weight | 0.213 | 0.204 |
|  | Lost weight | 0.512 | 0.529 |
|  | Thoughts of death | 0.609 | 0.627 |
| *Currently depressed* | Lack of interest or pleasure | 0.889 | 0.911 |
|  | Feelings of depression | 0.877 | 0.888 |
|  | Tiredness of low energy | 0.916 | 0.915 |
|  | Over or under sleeping | 0.854 | 0.852 |
|  | Feelings of inadequacy | 0.742 | 0.707 |
|  | Trouble concentrating | 0.667 | 0.650 |
|  | Change in appetite | 0.670 | 0.653 |
|  | Suicidal thoughts or self-harm | 0.568 | 0.530 |
|  | Psychomotor changes | 0.286 | 0.243 |

Supplementary Table 2. *P*-values threshold for significance in the regression analyses reported in Supplementary Tables 2-197 using an alpha of 0.05 and applying a Bonferroni correction

| Group | *P*-value threshold for significance |
| --- | --- |
| Q1 *Ever depressed* | 1.43 × 10^-4^ |
| Q2 *Ever depressed* | 1.54 × 10^-4^ |
| Q1 *Currently depressed* | 1.82 × 10^-4^ |
| Q2 *Currently depressed* | 1.89 × 10^-4^ |

### Q1 (Mental Health Questionnaire) - *Ever depressed*

Number of individuals analysed: 14,864

Number of clusters identified: 14

#### Atypical Symptoms

Supplementary Table 3. Multivariable linear regression of individuals’ probability scores for Atypical Symptoms on demographic variables.

| Demographic variables | Effect Size | Standard Error | *P*-value |
| --- | --- | --- | --- |
| Age | -0.00246 | 0.00024 | **<2.00 × 10^-16^** |
| Sex | 0.00113 | 0.00391 | 0.77 |
| Ethnicity - Asian | -0.02237 | 0.02232 | 0.32 |
| Ethnicity - Black | -0.00743 | 0.02324 | 0.75 |
| Ethnicity - Chinese | 0.00232 | 0.04134 | 0.96 |
| Ethnicity - Mixed | 0.00004 | 0.02478 | 1.00 |
| Ethnicity - Other | -0.02921 | 0.02873 | 0.31 |
| Place of Birth | 0.00363 | 0.00772 | 0.64 |
| Townsend Deprivation Index | 0.00749 | 0.00191 | **9.05 × 10^-5^** |
| Smoking - Former | -0.00857 | 0.00669 | 0.20 |
| Smoking - Never | -0.01442 | 0.00646 | 0.026 |
| Body Mass Index | 0.03024 | 0.00178 | **<2.00 × 10^-16^** |

Probability scores were calculated using Bernoulli-mixtures applied to ever depressed individuals at Q1. All variables were fitted simultaneously. Sex, ethnicity (European ethnicity as the reference), place of birth, and smoking (current smoking as the reference) were fitted as factors. Bold values indicate P-values that were significant after Bonferroni correction.

Supplementary Table 4. Multivariable linear regression of individuals’ probability scores for Atypical Symptoms on health variables.

| Health variables | Effect Size | Standard Error | *P*-value |
| --- | --- | --- | --- |
| Myocardial infarction | 0.02345 | 0.00991 | 0.018 |
| Stroke | 0.03410 | 0.01172 | 0.0036 |
| Asthma | 0.01624 | 0.00482 | 7.62 × 10^-4^ |
| COPD | 0.01530 | 0.00956 | 0.11 |
| Dementia | 0.01432 | 0.02802 | 0.61 |
| End stage renal disease | 0.02263 | 0.04541 | 0.62 |
| Motor neurone disease | -0.06498 | 0.06559 | 0.32 |
| Parkinson’s disease | 0.00567 | 0.02541 | 0.82 |

Probability scores were calculated using Bernoulli-mixtures applied to ever depressed individuals at Q1. All variables were fitted simultaneously with age at questionnaire and sex fitted as covariates. Bold values indicate P-values that were significant after Bonferroni correction. COPD = chronic obstructive pulmonary disease.

Supplementary Table 5. Linear regressions of individuals’ probability scores for Atypical Symptoms on depression recurrence and on treatment resistant depression.

| Depression | Effect Size | Standard Error | *P*-value |
| --- | --- | --- | --- |
| Recurrent | 0.03003 | 0.00365 | **<2.00 × 10^-16^** |
| Treatment resistance | 0.02354 | 0.02394 | 0.33 |

Recurrence and treatment resistance were both fitted as factors and were examined separately with age at questionnaire and sex fitted as covariates. Bold values indicate P-values that were significant after Bonferroni correction.

Supplementary Table 6. Linear regressions of individuals’ probability scores for Atypical Symptoms on polygenic scores for bipolar disorder, schizophrenia, and Attention-Deficit/Hyperactivity Disorder (ADHD).

| Mental health polygenic scores | Effect Size | Standard Error | *P*-value |
| --- | --- | --- | --- |
| Bipolar disorder | 0.00311 | 0.00186 | 0.10 |
| Schizophrenia | 0.00247 | 0.00184 | 0.18 |
| ADHD | 0.00787 | 0.00187 | **2.44 × 10^-5^** |

The polygenic scores for each trait were examined separately with age, sex, ancestry, and the first 10 genetic principal components fitted as covariates. Bold values indicate P-values that were significant after Bonferroni correction.

#### Hypersomnia, Weight Loss, and Mortality-Focus

Supplementary Table 7. Multivariable linear regression of individuals’ probability scores for Hypersomnia, Weight Loss, and Mortality-Focus on demographic variables.

| Demographic variables | Effect Size | Standard Error | *P*-value |
| --- | --- | --- | --- |
| Age | -0.00078 | 0.00017 | **3.87 × 10^-6^** |
| Sex | -0.00549 | 0.00272 | 0.044 |
| Ethnicity - Asian | -0.01190 | 0.01556 | 0.44 |
| Ethnicity - Black | 0.03761 | 0.01620 | 0.020 |
| Ethnicity - Chinese | -0.01097 | 0.02882 | 0.70 |
| Ethnicity - Mixed | 0.02821 | 0.01727 | 0.10 |
| Ethnicity - Other | 0.00214 | 0.02003 | 0.92 |
| Place of Birth | 0.00050 | 0.00538 | 0.93 |
| Townsend Deprivation Index | 0.00184 | 0.00133 | 0.17 |
| Smoking - Former | -0.01290 | 0.00466 | 0.006 |
| Smoking - Never | -0.01824 | 0.00450 | **5.15 × 10^-5^** |
| Body Mass Index | -0.00343 | 0.00124 | 0.006 |

Probability scores were calculated using Bernoulli-mixtures applied to ever depressed individuals at Q1. All variables were fitted simultaneously. Sex, ethnicity (European ethnicity as the reference), place of birth, and smoking (current smoking as the reference) were fitted as factors. Bold values indicate P-values that were significant after Bonferroni correction.

Supplementary Table 8. Multivariable linear regression of individuals’ probability scores for Hypersomnia, Weight Loss, and Mortality-Focus on health variables.

| Health variables | Effect Size | Standard Error | *P*-value |
| --- | --- | --- | --- |
| Myocardial infarction | 0.00505 | 0.00676 | 0.45 |
| Stroke | -0.00415 | 0.00799 | 0.60 |
| Asthma | 0.00426 | 0.00329 | 0.20 |
| COPD | 0.00945 | 0.00652 | 0.15 |
| Dementia | 0.05980 | 0.01911 | 0.0018 |
| End stage renal disease | 0.05170 | 0.03097 | 0.10 |
| Motor neurone disease | 0.05146 | 0.04474 | 0.25 |
| Parkinson’s disease | -0.00219 | 0.01733 | 0.90 |

Probability scores were calculated using Bernoulli-mixtures applied to ever depressed individuals at Q1. All variables were fitted simultaneously with age at questionnaire and sex fitted as covariates. Bold values indicate P-values that were significant after Bonferroni correction. COPD = chronic obstructive pulmonary disease.

Supplementary Table 9. Linear regressions of individuals’ probability scores for Hypersomnia, Weight Loss, and Mortality-Focus on depression recurrence and on treatment resistant depression.

| Depression | Effect Size | Standard Error | *P*-value |
| --- | --- | --- | --- |
| Recurrent | 0.00768 | 0.00251 | 0.0022 |
| Treatment resistance | 0.00576 | 0.01568 | 0.71 |

Recurrence and treatment resistance were both fitted as factors and were examined separately with age at questionnaire and sex fitted as covariates. Bold values indicate P-values that were significant after Bonferroni correction.

Supplementary Table 10. Linear regressions of individuals’ probability scores for Hypersomnia, Weight Loss, and Mortality-Focus on polygenic scores for bipolar disorder, schizophrenia, and Attention-Deficit/Hyperactivity Disorder (ADHD).

| Mental health polygenic scores | Effect Size | Standard Error | *P*-value |
| --- | --- | --- | --- |
| Bipolar disorder | -0.00094 | 0.00127 | 0.46 |
| Schizophrenia | 0.00323 | 0.00125 | 0.010 |
| ADHD | 0.00244 | 0.00127 | 0.06 |

The polygenic scores for each trait were examined separately with age, sex, ancestry, and the first 10 genetic principal components fitted as covariates. Bold values indicate P-values that were significant after Bonferroni correction.

#### Hypersomnia and Weight Loss

Supplementary Table 11. Multivariable linear regression of individuals’ probability scores for Hypersomnia and Weight Loss on demographic variables.

| Demographic variables | Effect Size | Standard Error | *P*-value |
| --- | --- | --- | --- |
| Age | -0.00009 | 0.00017 | 0.60 |
| Sex | 0.00337 | 0.00269 | 0.21 |
| Ethnicity - Asian | -0.00669 | 0.01536 | 0.66 |
| Ethnicity - Black | -0.01662 | 0.01599 | 0.30 |
| Ethnicity - Chinese | 0.01384 | 0.02844 | 0.63 |
| Ethnicity - Mixed | -0.01365 | 0.01705 | 0.42 |
| Ethnicity - Other | 0.00665 | 0.01977 | 0.74 |
| Place of Birth | -0.00514 | 0.00531 | 0.33 |
| Townsend Deprivation Index | 0.00070 | 0.00132 | 0.60 |
| Smoking - Former | -0.00847 | 0.00460 | 0.07 |
| Smoking - Never | -0.01348 | 0.00445 | 0.0024 |
| Body Mass Index | -0.00113 | 0.00123 | 0.36 |

Probability scores were calculated using Bernoulli-mixtures applied to ever depressed individuals at Q1. All variables were fitted simultaneously. Sex, ethnicity (European ethnicity as the reference), place of birth, and smoking (current smoking as the reference) were fitted as factors. Bold values indicate P-values that were significant after Bonferroni correction.

Supplementary Table 12. Multivariable linear regression of individuals’ probability scores for Hypersomnia and Weight Loss on health variables.

| Health variables | Effect Size | Standard Error | *P*-value |
| --- | --- | --- | --- |
| Myocardial infarction | 0.00401 | 0.00263 | 0.13 |
| Stroke | 0.00579 | 0.00665 | 0.38 |
| Asthma | -0.00569 | 0.00786 | 0.47 |
| COPD | -0.00317 | 0.00324 | 0.33 |
| Dementia | 0.00123 | 0.00641 | 0.85 |
| End stage renal disease | -0.00651 | 0.01880 | 0.73 |
| Motor neurone disease | -0.00351 | 0.03046 | 0.91 |
| Parkinson’s disease | -0.02963 | 0.04400 | 0.50 |

Probability scores were calculated using Bernoulli-mixtures applied to ever depressed individuals at Q1. All variables were fitted simultaneously with age at questionnaire and sex fitted as covariates. Bold values indicate P-values that were significant after Bonferroni correction. COPD = chronic obstructive pulmonary disease.

Supplementary Table 13. Linear regressions of individuals’ probability scores for Hypersomnia and Weight Loss on depression recurrence and on treatment resistant depression.

| Depression | Effect Size | Standard Error | *P*-value |
| --- | --- | --- | --- |
| Recurrent | -0.01458 | 0.00247 | **3.60 × 10^-9^** |
| Treatment resistance | -0.00707 | 0.01430 | 0.62 |

Recurrence and treatment resistance were both fitted as factors and were examined separately with age at questionnaire and sex fitted as covariates. Bold values indicate P-values that were significant after Bonferroni correction.

Supplementary Table 14. Linear regressions of individuals’ probability scores for Hypersomnia and Weight Loss on polygenic scores for bipolar disorder, schizophrenia, and Attention-Deficit/Hyperactivity Disorder (ADHD).

| Mental health polygenic scores | Effect Size | Standard Error | *P*-value |
| --- | --- | --- | --- |
| Bipolar disorder | 0.00088 | 0.00125 | 0.48 |
| Schizophrenia | -0.00054 | 0.00123 | 0.66 |
| ADHD | 0.00005 | 0.00125 | 0.97 |

The polygenic scores for each trait were examined separately with age, sex, ancestry, and the first 10 genetic principal components fitted as covariates. Bold values indicate P-values that were significant after Bonferroni correction.

#### Prolonged Sleep Latency and Weight Loss

Supplementary Table 15. Multivariable linear regression of individuals’ probability scores for Prolonged Sleep Latency and Weight Loss on demographic variables.

| Demographic variables | Effect Size | Standard Error | *P*-value |
| --- | --- | --- | --- |
| Age | -0.00025 | 0.00010 | 0.016 |
| Sex | -0.00495 | 0.00168 | 0.0033 |
| Ethnicity - Asian | 0.01390 | 0.00962 | 0.15 |
| Ethnicity - Black | 0.01125 | 0.01001 | 0.26 |
| Ethnicity - Chinese | 0.05967 | 0.01781 | 8.08 × 10^-4^ |
| Ethnicity - Mixed | 0.01102 | 0.01067 | 0.30 |
| Ethnicity - Other | 0.00716 | 0.01238 | 0.56 |
| Place of Birth | -0.00520 | 0.00332 | 0.12 |
| Townsend Deprivation Index | 0.00081 | 0.00082 | 0.33 |
| Smoking - Former | -0.00465 | 0.00288 | 0.11 |
| Smoking - Never | -0.00805 | 0.00278 | 0.0038 |
| Body Mass Index | -0.00245 | 0.00077 | 0.0014 |

Probability scores were calculated using Bernoulli-mixtures applied to ever depressed individuals at Q1. All variables were fitted simultaneously. Sex, ethnicity (European ethnicity as the reference), place of birth, and smoking (current smoking as the reference) were fitted as factors. Bold values indicate P-values that were significant after Bonferroni correction.

Supplementary Table 16. Multivariable linear regression of individuals’ probability scores for Prolonged Sleep Latency and Weight Loss on health variables.

| Health variables | Effect Size | Standard Error | *P*-value |
| --- | --- | --- | --- |
| Myocardial infarction | 0.00003 | 0.00417 | 0.99 |
| Stroke | -0.00231 | 0.00493 | 0.64 |
| Asthma | -0.00112 | 0.00203 | 0.58 |
| COPD | 0.00719 | 0.00402 | 0.07 |
| Dementia | 0.00984 | 0.01178 | 0.40 |
| End stage renal disease | 0.03944 | 0.01909 | 0.039 |
| Motor neurone disease | -0.01800 | 0.02757 | 0.51 |
| Parkinson’s disease | -0.00154 | 0.01068 | 0.89 |

Probability scores were calculated using Bernoulli-mixtures applied to ever depressed individuals at Q1. All variables were fitted simultaneously with age at questionnaire and sex fitted as covariates. Bold values indicate P-values that were significant after Bonferroni correction. COPD = chronic obstructive pulmonary disease.

Supplementary Table 17. Linear regressions of individuals’ probability scores for Prolonged Sleep Latency and Weight Loss on depression recurrence and on treatment resistant depression.

| Depression | Effect Size | Standard Error | *P*-value |
| --- | --- | --- | --- |
| Recurrent | 0.00065 | 0.00153 | 0.67 |
| Treatment resistance | 0.00420 | 0.01060 | 0.69 |

Recurrence and treatment resistance were both fitted as factors and were examined separately with age at questionnaire and sex fitted as covariates. Bold values indicate P-values that were significant after Bonferroni correction.

Supplementary Table 18. Linear regressions of individuals’ probability scores for Prolonged Sleep Latency and Weight Loss on polygenic scores for bipolar disorder, schizophrenia, and Attention-Deficit/Hyperactivity Disorder (ADHD).

| Mental health polygenic scores | Effect Size | Standard Error | *P*-value |
| --- | --- | --- | --- |
| Bipolar disorder | -0.00042 | 0.00078 | 0.60 |
| Schizophrenia | -0.00052 | 0.00077 | 0.50 |
| ADHD | -0.00061 | 0.00078 | 0.44 |

The polygenic scores for each trait were examined separately with age, sex, ancestry, and the first 10 genetic principal components fitted as covariates. Bold values indicate P-values that were significant after Bonferroni correction.

#### Terminal Insomnia

Supplementary Table 19. Multivariable linear regression of individuals’ probability scores for Terminal Insomnia on demographic variables.

| Demographic variables | Effect Size | Standard Error | *P*-value |
| --- | --- | --- | --- |
| Age | -0.00085 | 0.00019 | **1.29 × 10^-5^** |
| Sex | 0.00735 | 0.00312 | 0.019 |
| Ethnicity - Asian | -0.03451 | 0.01785 | 0.05 |
| Ethnicity - Black | -0.04287 | 0.01858 | 0.021 |
| Ethnicity - Chinese | -0.03374 | 0.03305 | 0.31 |
| Ethnicity - Mixed | -0.01158 | 0.01981 | 0.56 |
| Ethnicity - Other | -0.02772 | 0.02297 | 0.23 |
| Place of Birth | 0.00860 | 0.00617 | 0.16 |
| Townsend Deprivation Index | -0.00300 | 0.00153 | 0.050 |
| Smoking - Former | 0.01636 | 0.00535 | 0.0022 |
| Smoking - Never | 0.01574 | 0.00517 | 0.0023 |
| Body Mass Index | 0.00583 | 0.00142 | **4.20 × 10^-5^** |

Probability scores were calculated using Bernoulli-mixtures applied to ever depressed individuals at Q1. All variables were fitted simultaneously. Sex, ethnicity (European ethnicity as the reference), place of birth, and smoking (current smoking as the reference) were fitted as factors. Bold values indicate P-values that were significant after Bonferroni correction.

Supplementary Table 20. Multivariable linear regression of individuals’ probability scores for Terminal Insomnia on health variables.

| Health variables | Effect Size | Standard Error | *P*-value |
| --- | --- | --- | --- |
| Myocardial infarction | 0.00682 | 0.00305 | 0.40 |
| Stroke | 0.00648 | 0.00773 | 0.50 |
| Asthma | 0.00612 | 0.00914 | 0.25 |
| COPD | -0.00430 | 0.00376 | 0.69 |
| Dementia | 0.00293 | 0.00746 | 0.29 |
| End stage renal disease | -0.02291 | 0.02185 | 0.63 |
| Motor neurone disease | -0.01728 | 0.03541 | 0.35 |
| Parkinson’s disease | -0.04795 | 0.05115 | 0.0024 |

Probability scores were calculated using Bernoulli-mixtures applied to ever depressed individuals at Q1. All variables were fitted simultaneously with age at questionnaire and sex fitted as covariates. Bold values indicate P-values that were significant after Bonferroni correction. COPD = chronic obstructive pulmonary disease.

Supplementary Table 21. Linear regressions of individuals’ probability scores for Terminal Insomnia on depression recurrence and on treatment resistant depression.

| Depression | Effect Size | Standard Error | *P*-value |
| --- | --- | --- | --- |
| Recurrent | 0.00291 | 0.00286 | 0.31 |
| Treatment resistance | 0.00503 | 0.01617 | 0.76 |

Recurrence and treatment resistance were both fitted as factors and were examined separately with age at questionnaire and sex fitted as covariates. Bold values indicate P-values that were significant after Bonferroni correction.

Supplementary Table 22. Linear regressions of individuals’ probability scores for Terminal Insomnia on polygenic scores for bipolar disorder, schizophrenia, and Attention-Deficit/Hyperactivity Disorder (ADHD).

| Mental health polygenic scores | Effect Size | Standard Error | *P*-value |
| --- | --- | --- | --- |
| Bipolar disorder | 0.00120 | 0.00146 | 0.41 |
| Schizophrenia | 0.00027 | 0.00144 | 0.85 |
| ADHD | -0.00246 | 0.00146 | 0.09 |

The polygenic scores for each trait were examined separately with age, sex, ancestry, and the first 10 genetic principal components fitted as covariates. Bold values indicate P-values that were significant after Bonferroni correction.

#### Insomnia, Weight Gain and Mortality-Focus

Supplementary Table 23. Multivariable linear regression of individuals’ probability scores for Insomnia, Weight Gain and Mortality-Focus on demographic variables.

| Demographic variables | Effect Size | Standard Error | *P*-value |
| --- | --- | --- | --- |
| Age | -0.00176 | 0.00019 | **<2.00 × 10^-16^** |
| Sex | -0.01230 | 0.00298 | **3.73 × 10^-5^** |
| Ethnicity - Asian | 0.01942 | 0.01704 | 0.25 |
| Ethnicity - Black | -0.00719 | 0.01774 | 0.69 |
| Ethnicity - Chinese | -0.01978 | 0.03156 | 0.53 |
| Ethnicity - Mixed | 0.00104 | 0.01891 | 0.96 |
| Ethnicity - Other | 0.02863 | 0.02193 | 0.19 |
| Place of Birth | 0.00116 | 0.00589 | 0.84 |
| Townsend Deprivation Index | 0.00131 | 0.00146 | 0.37 |
| Smoking - Former | -0.00823 | 0.00510 | 0.11 |
| Smoking - Never | -0.00410 | 0.00493 | 0.41 |
| Body Mass Index | 0.03352 | 0.00136 | **<2.00 × 10^-16^** |

Probability scores were calculated using Bernoulli-mixtures applied to ever depressed individuals at Q1. All variables were fitted simultaneously. Sex, ethnicity (European ethnicity as the reference), place of birth, and smoking (current smoking as the reference) were fitted as factors. Bold values indicate P-values that were significant after Bonferroni correction.

Supplementary Table 24. Multivariable linear regression of individuals’ probability scores for Insomnia, Weight Gain and Mortality-Focus on health variables.

| Health variables | Effect Size | Standard Error | *P*-value |
| --- | --- | --- | --- |
| Myocardial infarction | 0.01051 | 0.00757 | 0.17 |
| Stroke | 0.01313 | 0.00895 | 0.14 |
| Asthma | 0.01824 | 0.00368 | **7.44 × 10^-7^** |
| COPD | 0.01251 | 0.00730 | 0.09 |
| Dementia | 0.00258 | 0.02141 | 0.90 |
| End stage renal disease | -0.05801 | 0.03469 | 0.09 |
| Motor neurone disease | -0.02819 | 0.05011 | 0.57 |
| Parkinson’s disease | -0.01604 | 0.01941 | 0.41 |

Probability scores were calculated using Bernoulli-mixtures applied to ever depressed individuals at Q1. All variables were fitted simultaneously with age at questionnaire and sex fitted as covariates. Bold values indicate P-values that were significant after Bonferroni correction. COPD = chronic obstructive pulmonary disease.

Supplementary Table 25. Linear regressions of individuals’ probability scores for Insomnia, Weight Gain and Mortality-Focus on depression recurrence and on treatment resistant depression.

| Depression | Effect Size | Standard Error | *P*-value |
| --- | --- | --- | --- |
| Recurrent | 0.03929 | 0.00277 | **<2.00 × 10^-16^** |
| Treatment resistance | 0.01676 | 0.01833 | 0.36 |

Recurrence and treatment resistance were both fitted as factors and were examined separately with age at questionnaire and sex fitted as covariates. Bold values indicate P-values that were significant after Bonferroni correction.

Supplementary Table 26. Linear regressions of individuals’ probability scores for Insomnia, Weight Gain and Mortality-Focus on polygenic scores for bipolar disorder, schizophrenia, and Attention-Deficit/Hyperactivity Disorder (ADHD).

| Mental health polygenic scores | Effect Size | Standard Error | *P*-value |
| --- | --- | --- | --- |
| Bipolar disorder | 0.00076 | 0.00142 | 0.59 |
| Schizophrenia | -0.00132 | 0.00140 | 0.35 |
| ADHD | 0.00599 | 0.00142 | **2.57 × 10^-5^** |

The polygenic scores for each trait were examined separately with age, sex, ancestry, and the first 10 genetic principal components fitted as covariates. Bold values indicate P-values that were significant after Bonferroni correction.

#### Insomnia and Weight Gain

Supplementary Table 27. Multivariable linear regression of individuals’ probability scores for Insomnia and Weight Gain on demographic variables.

| Demographic variables | Effect Size | Standard Error | *P*-value |
| --- | --- | --- | --- |
| Age | -0.00145 | 0.00027 | **6.04 × 10^-8^** |
| Sex | -0.02316 | 0.00428 | **6.44 × 10^-8^** |
| Ethnicity - Asian | 0.00376 | 0.02447 | 0.88 |
| Ethnicity - Black | -0.05013 | 0.02548 | 0.049 |
| Ethnicity - Chinese | 0.00683 | 0.04531 | 0.88 |
| Ethnicity - Mixed | 0.01330 | 0.02716 | 0.62 |
| Ethnicity - Other | -0.03769 | 0.03149 | 0.23 |
| Place of Birth | 0.01853 | 0.00846 | 0.029 |
| Townsend Deprivation Index | 0.00053 | 0.00210 | 0.80 |
| Smoking - Former | -0.00462 | 0.00733 | 0.53 |
| Smoking - Never | 0.00785 | 0.00708 | 0.27 |
| Body Mass Index | 0.05200 | 0.00195 | **<2.00 × 10^-16^** |

Probability scores were calculated using Bernoulli-mixtures applied to ever depressed individuals at Q1. All variables were fitted simultaneously. Sex, ethnicity (European ethnicity as the reference), place of birth, and smoking (current smoking as the reference) were fitted as factors. Bold values indicate P-values that were significant after Bonferroni correction.

Supplementary Table 28. Multivariable linear regression of individuals’ probability scores for Insomnia and Weight Gain on health variables.

| Health variables | Effect Size | Standard Error | *P*-value |
| --- | --- | --- | --- |
| Myocardial infarction | 0.00574 | 0.01088 | 0.60 |
| Stroke | 0.00020 | 0.01286 | 0.99 |
| Asthma | 0.01480 | 0.00530 | 0.0052 |
| COPD | 0.00703 | 0.01050 | 0.50 |
| Dementia | -0.00376 | 0.03077 | 0.90 |
| End stage renal disease | -0.04413 | 0.04986 | 0.38 |
| Motor neurone disease | -0.07738 | 0.07201 | 0.28 |
| Parkinson’s disease | -0.04712 | 0.02790 | 0.09 |

Probability scores were calculated using Bernoulli-mixtures applied to ever depressed individuals at Q1. All variables were fitted simultaneously with age at questionnaire and sex fitted as covariates. Bold values indicate P-values that were significant after Bonferroni correction. COPD = chronic obstructive pulmonary disease.

Supplementary Table 29. Linear regressions of individuals’ probability scores for Insomnia and Weight Gain on depression recurrence and on treatment resistant depression.

| Depression | Effect Size | Standard Error | *P*-value |
| --- | --- | --- | --- |
| Recurrent | 0.01359 | 0.00402 | 7.16 × 10^-4^ |
| Treatment resistance | 0.02887 | 0.02239 | 0.20 |

Recurrence and treatment resistance were both fitted as factors and were examined separately with age at questionnaire and sex fitted as covariates. Bold values indicate P-values that were significant after Bonferroni correction.

Supplementary Table 30. Linear regressions of individuals’ probability scores for Insomnia and Weight Gain on polygenic scores for bipolar disorder, schizophrenia, and Attention-Deficit/Hyperactivity Disorder (ADHD).

| Mental health polygenic scores | Effect Size | Standard Error | *P*-value |
| --- | --- | --- | --- |
| Bipolar disorder | 0.00002 | 0.00203 | 0.99 |
| Schizophrenia | -0.00178 | 0.00200 | 0.37 |
| ADHD | 0.00549 | 0.00203 | 0.0070 |

The polygenic scores for each trait were examined separately with age, sex, ancestry, and the first 10 genetic principal components fitted as covariates. Bold values indicate P-values that were significant after Bonferroni correction.

#### Insomnia without Weight Change

Supplementary Table 31. Multivariable linear regression of individuals’ probability scores for Insomnia without Weight Change on demographic variables.

| Demographic variables | Effect Size | Standard Error | *P*-value |
| --- | --- | --- | --- |
| Age | 0.00124 | 0.00019 | **2.53 × 10^-11^** |
| Sex | 0.03648 | 0.00299 | **<2.00 × 10^-16^** |
| Ethnicity - Asian | 0.02372 | 0.01709 | 0.17 |
| Ethnicity - Black | 0.00622 | 0.01780 | 0.73 |
| Ethnicity - Chinese | -0.02267 | 0.03166 | 0.47 |
| Ethnicity - Mixed | -0.01019 | 0.01897 | 0.59 |
| Ethnicity - Other | -0.02051 | 0.02200 | 0.35 |
| Place of Birth | -0.00798 | 0.00591 | 0.18 |
| Townsend Deprivation Index | -0.00447 | 0.00146 | 0.0023 |
| Smoking - Former | 0.00747 | 0.00512 | 0.14 |
| Smoking - Never | 0.01064 | 0.00495 | 0.03 |
| Body Mass Index | -0.00485 | 0.00136 | 3.72 × 10^-4^ |

Probability scores were calculated using Bernoulli-mixtures applied to ever depressed individuals at Q1. All variables were fitted simultaneously. Sex, ethnicity (European ethnicity as the reference), place of birth, and smoking (current smoking as the reference) were fitted as factors. Bold values indicate P-values that were significant after Bonferroni correction.

Supplementary Table 32. Multivariable linear regression of individuals’ probability scores for Insomnia without Weight Change on health variables.

| Health variables | Effect Size | Standard Error | *P*-value |
| --- | --- | --- | --- |
| Myocardial infarction | -0.00291 | 0.00728 | 0.69 |
| Stroke | -0.00631 | 0.00861 | 0.46 |
| Asthma | -0.00362 | 0.00355 | 0.31 |
| COPD | -0.00504 | 0.00703 | 0.47 |
| Dementia | -0.03888 | 0.02060 | 0.06 |
| End stage renal disease | 0.04409 | 0.03338 | 0.19 |
| Motor neurone disease | -0.04102 | 0.04821 | 0.39 |
| Parkinson’s disease | 0.01351 | 0.01868 | 0.47 |

Probability scores were calculated using Bernoulli-mixtures applied to ever depressed individuals at Q1. All variables were fitted simultaneously with age at questionnaire and sex fitted as covariates. Bold values indicate P-values that were significant after Bonferroni correction. COPD = chronic obstructive pulmonary disease.

Supplementary Table 33. Linear regressions of individuals’ probability scores for Insomnia without Weight Change on depression recurrence and on treatment resistant depression.

| Depression | Effect Size | Standard Error | *P*-value |
| --- | --- | --- | --- |
| Recurrent | -0.01415 | 0.00268 | **1.37 × 10^-7^** |
| Treatment resistance | -0.02244 | 0.01378 | 0.10 |

Recurrence and treatment resistance were both fitted as factors and were examined separately with age at questionnaire and sex fitted as covariates. Bold values indicate P-values that were significant after Bonferroni correction.

Supplementary Table 34. Linear regressions of individuals’ probability scores for Insomnia without Weight Change on polygenic scores for bipolar disorder, schizophrenia, and Attention-Deficit/Hyperactivity Disorder (ADHD).

| Mental health polygenic scores | Effect Size | Standard Error | *P*-value |
| --- | --- | --- | --- |
| Bipolar disorder | -0.00224 | 0.00137 | 0.10 |
| Schizophrenia | -0.00240 | 0.00135 | 0.08 |
| ADHD | -0.00173 | 0.00138 | 0.21 |

The polygenic scores for each trait were examined separately with age, sex, ancestry, and the first 10 genetic principal components fitted as covariates. Bold values indicate P-values that were significant after Bonferroni correction.

#### Insomnia and Low Self-worth

Supplementary Table 35. Multivariable linear regression of individuals’ probability scores for Insomnia and Low Self-worth on demographic variables.

| Demographic variables | Effect Size | Standard Error | *P*-value |
| --- | --- | --- | --- |
| Age | 0.00060 | 0.00025 | 0.018 |
| Sex | 0.05109 | 0.00406 | **<2.00 × 10^-16^** |
| Ethnicity - Asian | 0.00441 | 0.02319 | 0.85 |
| Ethnicity - Black | -0.01643 | 0.02414 | 0.50 |
| Ethnicity - Chinese | 0.13497 | 0.04294 | 0.0017 |
| Ethnicity - Mixed | -0.02220 | 0.02574 | 0.39 |
| Ethnicity - Other | 0.01396 | 0.02984 | 0.64 |
| Place of Birth | 0.01042 | 0.00802 | 0.19 |
| Townsend Deprivation Index | 0.00079 | 0.00199 | 0.69 |
| Smoking - Former | 0.00701 | 0.00695 | 0.31 |
| Smoking - Never | 0.00993 | 0.00671 | 0.14 |
| Body Mass Index | -0.02492 | 0.00185 | **<2.00 × 10^-16^** |

Probability scores were calculated using Bernoulli-mixtures applied to ever depressed individuals at Q1. All variables were fitted simultaneously. Sex, ethnicity (European ethnicity as the reference), place of birth, and smoking (current smoking as the reference) were fitted as factors. Bold values indicate P-values that were significant after Bonferroni correction.

Supplementary Table 36. Multivariable linear regression of individuals’ probability scores for Insomnia and Low Self-worth on health variables.

| Health variables | Effect Size | Standard Error | *P*-value |
| --- | --- | --- | --- |
| Myocardial infarction | 0.00219 | 0.01010 | 0.83 |
| Stroke | 0.00053 | 0.01194 | 0.96 |
| Asthma | -0.01098 | 0.00492 | 0.026 |
| COPD | -0.02182 | 0.00975 | 0.025 |
| Dementia | 0.00887 | 0.02857 | 0.76 |
| End stage renal disease | 0.06010 | 0.04629 | 0.19 |
| Motor neurone disease | -0.01016 | 0.06686 | 0.88 |
| Parkinson’s disease | -0.05320 | 0.02590 | 0.040 |

Probability scores were calculated using Bernoulli-mixtures applied to ever depressed individuals at Q1. All variables were fitted simultaneously with age at questionnaire and sex fitted as covariates. Bold values indicate P-values that were significant after Bonferroni correction. COPD = chronic obstructive pulmonary disease.

Supplementary Table 37. Linear regressions of individuals’ probability scores for Insomnia and Low Self-worth on depression recurrence and on treatment resistant depression.

| Depression | Effect Size | Standard Error | *P*-value |
| --- | --- | --- | --- |
| Recurrent | -0.00597 | 0.00373 | 0.11 |
| Treatment resistance | -0.00446 | 0.01951 | 0.82 |

Recurrence and treatment resistance were both fitted as factors and were examined separately with age at questionnaire and sex fitted as covariates. Bold values indicate P-values that were significant after Bonferroni correction.

Supplementary Table 38. Linear regressions of individuals’ probability scores for Insomnia and Low Self-worth on polygenic scores for bipolar disorder, schizophrenia, and Attention-Deficit/Hyperactivity Disorder (ADHD).

| Mental health polygenic scores | Effect Size | Standard Error | *P*-value |
| --- | --- | --- | --- |
| Bipolar disorder | -0.00169 | 0.00190 | 0.37 |
| Schizophrenia | 0.00126 | 0.00187 | 0.50 |
| ADHD | -0.00600 | 0.00190 | 0.0016 |

The polygenic scores for each trait were examined separately with age, sex, ancestry, and the first 10 genetic principal components fitted as covariates. Bold values indicate P-values that were significant after Bonferroni correction.

#### Melancholia with Mortality-Focus

Supplementary Table 39. Multivariable linear regression of individuals’ probability scores for Melancholia with Mortality-Focus on demographic variables.

| Demographic variables | Effect Size | Standard Error | *P*-value |
| --- | --- | --- | --- |
| Age | 0.00135 | 0.00015 | **<2.00 × 10^-16^** |
| Sex | -0.00919 | 0.00240 | **1.32 × 10^-4^** |
| Ethnicity - Asian | -0.00926 | 0.01373 | 0.50 |
| Ethnicity - Black | 0.03402 | 0.01430 | 0.017 |
| Ethnicity - Chinese | 0.04156 | 0.02543 | 0.10 |
| Ethnicity - Mixed | -0.00489 | 0.01524 | 0.75 |
| Ethnicity - Other | 0.04508 | 0.01767 | 0.011 |
| Place of Birth | 0.00623 | 0.00475 | 0.19 |
| Townsend Deprivation Index | -0.00039 | 0.00118 | 0.74 |
| Smoking - Former | 0.01019 | 0.00411 | 0.013 |
| Smoking - Never | 0.00570 | 0.00397 | 0.15 |
| Body Mass Index | -0.01218 | 0.00110 | **<2.00 × 10^-16^** |

Probability scores were calculated using Bernoulli-mixtures applied to ever depressed individuals at Q1. All variables were fitted simultaneously. Sex, ethnicity (European ethnicity as the reference), place of birth, and smoking (current smoking as the reference) were fitted as factors. Bold values indicate P-values that were significant after Bonferroni correction.

Supplementary Table 40. Multivariable linear regression of individuals’ probability scores for Melancholia with Mortality-Focus on health variables.

| Health variables | Effect Size | Standard Error | *P*-value |
| --- | --- | --- | --- |
| Myocardial infarction | -0.01691 | 0.00596 | 0.0046 |
| Stroke | -0.01864 | 0.00705 | 0.008 |
| Asthma | -0.00791 | 0.00290 | 0.006 |
| COPD | -0.00315 | 0.00575 | 0.58 |
| Dementia | -0.00661 | 0.01686 | 0.69 |
| End stage renal disease | 0.00252 | 0.02732 | 0.93 |
| Motor neurone disease | -0.00776 | 0.03946 | 0.84 |
| Parkinson’s disease | 0.01351 | 0.01529 | 0.38 |

Probability scores were calculated using Bernoulli-mixtures applied to ever depressed individuals at Q1. All variables were fitted simultaneously with age at questionnaire and sex fitted as covariates. Bold values indicate P-values that were significant after Bonferroni correction. COPD = chronic obstructive pulmonary disease.

Supplementary Table 41. Linear regressions of individuals’ probability scores for Melancholia with Mortality-Focus on depression recurrence and on treatment resistant depression.

| Depression | Effect Size | Standard Error | *P*-value |
| --- | --- | --- | --- |
| Recurrent | -0.00850 | 0.00219 | **1.06 × 10^-4^** |
| Treatment resistance | 0.00036 | 0.01102 | 0.97 |

Recurrence and treatment resistance were both fitted as factors and were examined separately with age at questionnaire and sex fitted as covariates. Bold values indicate P-values that were significant after Bonferroni correction.

Supplementary Table 42. Linear regressions of individuals’ probability scores for Melancholia with Mortality-Focus on polygenic scores for bipolar disorder, schizophrenia, and Attention-Deficit/Hyperactivity Disorder (ADHD).

| Mental health polygenic scores | Effect Size | Standard Error | *P*-value |
| --- | --- | --- | --- |
| Bipolar disorder | -0.00098 | 0.00112 | 0.38 |
| Schizophrenia | 0.00087 | 0.00110 | 0.43 |
| ADHD | -0.00235 | 0.00112 | 0.037 |

The polygenic scores for each trait were examined separately with age, sex, ancestry, and the first 10 genetic principal components fitted as covariates. Bold values indicate P-values that were significant after Bonferroni correction.

#### Melancholia with Low Self-worth and Mortality-Focus

Supplementary Table 43. Multivariable linear regression of individuals’ probability scores for Melancholia with Low Self-worth and Mortality-Focus on demographic variables.

| Demographic variables | Effect Size | Standard Error | *P*-value |
| --- | --- | --- | --- |
| Age | -0.00009 | 0.00034 | 0.79 |
| Sex | -0.03259 | 0.00549 | **3.02 × 10^-9^** |
| Ethnicity - Asian | 0.01353 | 0.03138 | 0.67 |
| Ethnicity - Black | -0.01019 | 0.03268 | 0.76 |
| Ethnicity - Chinese | -0.11100 | 0.05812 | 0.06 |
| Ethnicity - Mixed | -0.03450 | 0.03483 | 0.32 |
| Ethnicity - Other | -0.00993 | 0.04039 | 0.81 |
| Place of Birth | -0.02338 | 0.01085 | 0.031 |
| Townsend Deprivation Index | 0.00080 | 0.00269 | 0.77 |
| Smoking - Former | -0.00171 | 0.00940 | 0.86 |
| Smoking - Never | -0.01231 | 0.00908 | 0.18 |
| Body Mass Index | -0.04258 | 0.00250 | **<2.00 × 10^-16^** |

Probability scores were calculated using Bernoulli-mixtures applied to ever depressed individuals at Q1. All variables were fitted simultaneously. Sex, ethnicity (European ethnicity as the reference), place of birth, and smoking (current smoking as the reference) were fitted as factors. Bold values indicate P-values that were significant after Bonferroni correction.

Supplementary Table 44. Multivariable linear regression of individuals’ probability scores for Melancholia with Low Self-worth and Mortality-Focus on health variables.

| Health variables | Effect Size | Standard Error | *P*-value |
| --- | --- | --- | --- |
| Myocardial infarction | -0.00498 | 0.01372 | 0.72 |
| Stroke | 0.00047 | 0.01623 | 0.98 |
| Asthma | -0.00480 | 0.00668 | 0.47 |
| COPD | -0.00516 | 0.01324 | 0.70 |
| Dementia | -0.01457 | 0.03881 | 0.71 |
| End stage renal disease | -0.06082 | 0.06288 | 0.33 |
| Motor neurone disease | 0.09904 | 0.09083 | 0.28 |
| Parkinson’s disease | 0.00984 | 0.03519 | 0.78 |

Probability scores were calculated using Bernoulli-mixtures applied to ever depressed individuals at Q1. All variables were fitted simultaneously with age at questionnaire and sex fitted as covariates. Bold values indicate P-values that were significant after Bonferroni correction. COPD = chronic obstructive pulmonary disease.

Supplementary Table 45. Linear regressions of individuals’ probability scores for Melancholia with Low Self-worth and Mortality-Focus on depression recurrence and on treatment resistant depression.

| Depression | Effect Size | Standard Error | *P*-value |
| --- | --- | --- | --- |
| Recurrent | 0.04040 | 0.00505 | **1.27 × 10^-15^** |
| Treatment resistance | 0.00613 | 0.03182 | 0.85 |

Recurrence and treatment resistance were both fitted as factors and were examined separately with age at questionnaire and sex fitted as covariates. Bold values indicate P-values that were significant after Bonferroni correction.

Supplementary Table 46. Linear regressions of individuals’ probability scores for Melancholia with Low Self-worth and Mortality-Focus on polygenic scores for bipolar disorder, schizophrenia, and Attention-Deficit/Hyperactivity Disorder (ADHD).

| Mental health polygenic scores | Effect Size | Standard Error | *P*-value |
| --- | --- | --- | --- |
| Bipolar disorder | 0.00830 | 0.00257 | 0.0013 |
| Schizophrenia | 0.00605 | 0.00254 | 0.017 |
| ADHD | 0.00033 | 0.00258 | 0.90 |

The polygenic scores for each trait were examined separately with age, sex, ancestry, and the first 10 genetic principal components fitted as covariates. Bold values indicate P-values that were significant after Bonferroni correction.

#### Melancholia with Low Self-worth

Supplementary Table 47. Multivariable linear regression of individuals’ probability scores for Melancholia with Low Self-worth on demographic variables.

| Demographic variables | Effect Size | Standard Error | *P*-value |
| --- | --- | --- | --- |
| Age | -0.00020 | 0.00013 | 0.11 |
| Sex | 0.00299 | 0.00205 | 0.14 |
| Ethnicity - Asian | -0.01522 | 0.01173 | 0.19 |
| Ethnicity - Black | -0.00554 | 0.01221 | 0.65 |
| Ethnicity - Chinese | -0.01626 | 0.02171 | 0.45 |
| Ethnicity - Mixed | -0.01967 | 0.01301 | 0.13 |
| Ethnicity - Other | -0.00464 | 0.01509 | 0.76 |
| Place of Birth | -0.00270 | 0.00405 | 0.51 |
| Townsend Deprivation Index | -0.00186 | 0.00100 | 0.06 |
| Smoking - Former | 0.00492 | 0.00351 | 0.16 |
| Smoking - Never | 0.00714 | 0.00339 | 0.04 |
| Body Mass Index | -0.00773 | 0.00094 | **<2.00 × 10^-16^** |

Probability scores were calculated using Bernoulli-mixtures applied to ever depressed individuals at Q1. All variables were fitted simultaneously. Sex, ethnicity (European ethnicity as the reference), place of birth, and smoking (current smoking as the reference) were fitted as factors. Bold values indicate P-values that were significant after Bonferroni correction.

Supplementary Table 48. Multivariable linear regression of individuals’ probability scores for Melancholia with Low Self-worth on health variables.

| Health variables | Effect Size | Standard Error | *P*-value |
| --- | --- | --- | --- |
| Myocardial infarction | -0.00354 | 0.00504 | 0.48 |
| Stroke | 0.01165 | 0.00596 | 0.051 |
| Asthma | 0.00011 | 0.00246 | 0.96 |
| COPD | -0.01152 | 0.00487 | 0.018 |
| Dementia | -0.00700 | 0.01427 | 0.62 |
| End stage renal disease | -0.01400 | 0.02312 | 0.54 |
| Motor neurone disease | -0.03430 | 0.03339 | 0.30 |
| Parkinson’s disease | 0.00670 | 0.01294 | 0.60 |

Probability scores were calculated using Bernoulli-mixtures applied to ever depressed individuals at Q1. All variables were fitted simultaneously with age at questionnaire and sex fitted as covariates. Bold values indicate P-values that were significant after Bonferroni correction. COPD = chronic obstructive pulmonary disease.

Supplementary Table 49. Linear regressions of individuals’ probability scores for Melancholia with Low Self-worth on depression recurrence and on treatment resistant depression.

| Depression | Effect Size | Standard Error | *P*-value |
| --- | --- | --- | --- |
| Recurrent | -0.00825 | 0.00188 | **1.11 × 10^-5^** |
| Treatment resistance | -0.02288 | 0.01082 | 0.035 |

Recurrence and treatment resistance were both fitted as factors and were examined separately with age at questionnaire and sex fitted as covariates. Bold values indicate P-values that were significant after Bonferroni correction.

Supplementary Table 50. Linear regressions of individuals’ probability scores for Melancholia with Low Self-worth on polygenic scores for bipolar disorder, schizophrenia, and Attention-Deficit/Hyperactivity Disorder (ADHD).

| Mental health polygenic scores | Effect Size | Standard Error | *P*-value |
| --- | --- | --- | --- |
| Bipolar disorder | 0.00086 | 0.00095 | 0.36 |
| Schizophrenia | -0.00007 | 0.00094 | 0.94 |
| ADHD | -0.00504 | 0.00095 | **1.25 × 10^-7^** |

The polygenic scores for each trait were examined separately with age, sex, ancestry, and the first 10 genetic principal components fitted as covariates. Bold values indicate P-values that were significant after Bonferroni correction.

#### Melancholia with Preserved Self-worth and Mortality-Focus

Supplementary Table 51. Multivariable linear regression of individuals’ probability scores for Melancholia with Preserved Self-worth and Mortality-Focus on demographic variables.

| Demographic variables | Effect Size | Standard Error | *P*-value |
| --- | --- | --- | --- |
| Age | 0.00286 | 0.00023 | **<2.00 × 10^-16^** |
| Sex | -0.01139 | 0.00371 | 0.0022 |
| Ethnicity - Asian | 0.01074 | 0.02121 | 0.61 |
| Ethnicity - Black | 0.04225 | 0.02208 | 0.06 |
| Ethnicity - Chinese | -0.01288 | 0.03928 | 0.74 |
| Ethnicity - Mixed | 0.03330 | 0.02354 | 0.16 |
| Ethnicity - Other | 0.00267 | 0.02730 | 0.92 |
| Place of Birth | 0.00134 | 0.00733 | 0.85 |
| Townsend Deprivation Index | -0.00087 | 0.00182 | 0.63 |
| Smoking - Former | 0.00130 | 0.00635 | 0.84 |
| Smoking - Never | 0.00826 | 0.00614 | 0.18 |
| Body Mass Index | -0.00860 | 0.00169 | **3.75 × 10^-7^** |

Probability scores were calculated using Bernoulli-mixtures applied to ever depressed individuals at Q1. All variables were fitted simultaneously. Sex, ethnicity (European ethnicity as the reference), place of birth, and smoking (current smoking as the reference) were fitted as factors. Bold values indicate P-values that were significant after Bonferroni correction.

Supplementary Table 52. Multivariable linear regression of individuals’ probability scores for Melancholia with Preserved Self-worth and Mortality-Focus on health variables.

| Health variables | Effect Size | Standard Error | *P*-value |
| --- | --- | --- | --- |
| Myocardial infarction | -0.00953 | 0.00915 | 0.30 |
| Stroke | -0.01679 | 0.01082 | 0.12 |
| Asthma | -0.01048 | 0.00446 | 0.019 |
| COPD | -0.01436 | 0.00883 | 0.10 |
| Dementia | -0.01135 | 0.02589 | 0.66 |
| End stage renal disease | 0.00685 | 0.04194 | 0.87 |
| Motor neurone disease | 0.04564 | 0.06059 | 0.45 |
| Parkinson’s disease | 0.02874 | 0.02347 | 0.22 |

Probability scores were calculated using Bernoulli-mixtures applied to ever depressed individuals at Q1. All variables were fitted simultaneously with age at questionnaire and sex fitted as covariates. Bold values indicate P-values that were significant after Bonferroni correction. COPD = chronic obstructive pulmonary disease.

Supplementary Table 53. Linear regressions of individuals’ probability scores for Melancholia with Preserved Self-worth and Mortality-Focus on depression recurrence and on treatment resistant depression.

| Depression | Effect Size | Standard Error | *P*-value |
| --- | --- | --- | --- |
| Recurrent | -0.02949 | 0.00338 | **<2.00 × 10^-16^** |
| Treatment resistance | -0.02454 | 0.01257 | 0.051 |

Recurrence and treatment resistance were both fitted as factors and were examined separately with age at questionnaire and sex fitted as covariates. Bold values indicate P-values that were significant after Bonferroni correction.

Supplementary Table 54. Linear regressions of individuals’ probability scores for Melancholia with Preserved Self-worth and Mortality-Focus on polygenic scores for bipolar disorder, schizophrenia, and Attention-Deficit/Hyperactivity Disorder (ADHD).

| Mental health polygenic scores | Effect Size | Standard Error | *P*-value |
| --- | --- | --- | --- |
| Bipolar disorder | -0.00275 | 0.00172 | 0.11 |
| Schizophrenia | -0.00309 | 0.00169 | 0.07 |
| ADHD | -0.00111 | 0.00172 | 0.52 |

The polygenic scores for each trait were examined separately with age, sex, ancestry, and the first 10 genetic principal components fitted as covariates. Bold values indicate P-values that were significant after Bonferroni correction.

#### Melancholia with Preserved Self-worth

Supplementary Table 55. Multivariable linear regression of individuals’ probability scores for Melancholia with Preserved Self-worth on demographic variables.

| Demographic variables | Effect Size | Standard Error | *P*-value |
| --- | --- | --- | --- |
| Age | 0.00188 | 0.00028 | **8.50 × 10^-12^** |
| Sex | -0.00333 | 0.00442 | 0.45 |
| Ethnicity - Asian | 0.01049 | 0.02524 | 0.68 |
| Ethnicity - Black | 0.02504 | 0.02629 | 0.34 |
| Ethnicity - Chinese | -0.03184 | 0.04675 | 0.50 |
| Ethnicity - Mixed | 0.02977 | 0.02802 | 0.29 |
| Ethnicity - Other | 0.02340 | 0.03249 | 0.47 |
| Place of Birth | -0.00601 | 0.00873 | 0.49 |
| Townsend Deprivation Index | -0.00368 | 0.00216 | 0.09 |
| Smoking - Former | 0.00189 | 0.00756 | 0.80 |
| Smoking - Never | 0.00535 | 0.00731 | 0.46 |
| Body Mass Index | -0.01372 | 0.00201 | **9.83 × 10^-12^** |

Probability scores were calculated using Bernoulli-mixtures applied to ever depressed individuals at Q1. All variables were fitted simultaneously. Sex, ethnicity (European ethnicity as the reference), place of birth, and smoking (current smoking as the reference) were fitted as factors. Bold values indicate P-values that were significant after Bonferroni correction.

Supplementary Table 56. Multivariable linear regression of individuals’ probability scores for Melancholia with Preserved Self-worth on health variables.

| Health variables | Effect Size | Standard Error | *P*-value |
| --- | --- | --- | --- |
| Myocardial infarction | -0.02137 | 0.01086 | 0.049 |
| Stroke | -0.01232 | 0.01285 | 0.34 |
| Asthma | -0.00727 | 0.00529 | 0.17 |
| COPD | 0.00542 | 0.01048 | 0.61 |
| Dementia | 0.01618 | 0.03073 | 0.60 |
| End stage renal disease | -0.02959 | 0.04979 | 0.55 |
| Motor neurone disease | 0.16325 | 0.07192 | 0.023 |
| Parkinson’s disease | -0.02155 | 0.02787 | 0.44 |

Probability scores were calculated using Bernoulli-mixtures applied to ever depressed individuals at Q1. All variables were fitted simultaneously with age at questionnaire and sex fitted as covariates. Bold values indicate P-values that were significant after Bonferroni correction. COPD = chronic obstructive pulmonary disease.

Supplementary Table 57. Linear regressions of individuals’ probability scores for Melancholia with Preserved Self-worth on depression recurrence and on treatment resistant depression.

| Depression | Effect Size | Standard Error | *P*-value |
| --- | --- | --- | --- |
| Recurrent | -0.05361 | 0.00400 | **<2.00 × 10^-16^** |
| Treatment resistance | -0.00926 | 0.01885 | 0.62 |

Recurrence and treatment resistance were both fitted as factors and were examined separately with age at questionnaire and sex fitted as covariates. Bold values indicate P-values that were significant after Bonferroni correction.

Supplementary Table 58. Linear regressions of individuals’ probability scores for Melancholia with Preserved Self-worth on polygenic scores for bipolar disorder, schizophrenia, and Attention-Deficit/Hyperactivity Disorder (ADHD).

| Mental health polygenic scores | Effect Size | Standard Error | *P*-value |
| --- | --- | --- | --- |
| Bipolar disorder | -0.00611 | 0.00204 | 0.0028 |
| Schizophrenia | -0.00442 | 0.00201 | 0.028 |
| ADHD | -0.00287 | 0.00205 | 0.16 |

The polygenic scores for each trait were examined separately with age, sex, ancestry, and the first 10 genetic principal components fitted as covariates. Bold values indicate P-values that were significant after Bonferroni correction.

### Q2 (Mental Well-being Questionnaire) - *Ever depressed*

Number of individuals analysed: 11,658

Number of clusters identified: 13

#### Atypical Symptoms

Supplementary Table 59. Multivariable linear regression of individuals’ probability scores for Atypical Symptoms on demographic variables.

| Demographic variables | Effect Size | Standard Error | *P*-value |
| --- | --- | --- | --- |
| Age | -0.00236 | 0.00034 | **3.10 × 10^-12^** |
| Sex | 0.00592 | 0.00536 | 0.27 |
| Ethnicity - Asian | -0.02923 | 0.03472 | 0.40 |
| Ethnicity - Black | 0.00281 | 0.02982 | 0.92 |
| Ethnicity - Chinese | -0.01169 | 0.06440 | 0.86 |
| Ethnicity - Mixed | 0.00102 | 0.02852 | 0.97 |
| Ethnicity - Other | -0.00536 | 0.03410 | 0.88 |
| Place of Birth | 0.02335 | 0.01068 | 0.029 |
| Townsend Deprivation Index | 0.00718 | 0.00260 | 0.0058 |
| Smoking - Former | -0.00217 | 0.00915 | 0.81 |
| Smoking - Never | -0.01241 | 0.00883 | 0.16 |
| Body Mass Index | 0.02358 | 0.00242 | **<2.00 × 10^-16^** |

Probability scores were calculated using Bernoulli-mixtures applied to ever depressed individuals at Q2. All variables were fitted simultaneously. Sex, ethnicity (European ethnicity as the reference), place of birth, and smoking (current smoking as the reference) were fitted as factors. Bold values indicate P-values that were significant after Bonferroni correction.

Supplementary Table 60. Multivariable linear regression of individuals’ probability scores for Atypical Symptoms on health variables.

| Health variables | Effect Size | Standard Error | *P*-value |
| --- | --- | --- | --- |
| Myocardial infarction | -0.00017 | 0.01502 | 0.99 |
| Stroke | 0.00727 | 0.02033 | 0.72 |
| Asthma | 0.00182 | 0.00639 | 0.78 |
| COPD | 0.02990 | 0.01449 | 0.039 |
| Dementia | 0.02873 | 0.10482 | 0.78 |
| End stage renal disease | -0.08942 | 0.06233 | 0.15 |
| Motor neurone disease | 0.15598 | 0.14814 | 0.29 |
| Parkinson’s disease | 0.10870 | 0.05360 | 0.043 |

Probability scores were calculated using Bernoulli-mixtures applied to ever depressed individuals at Q2. All variables were fitted simultaneously with age at questionnaire and sex fitted as covariates. Bold values indicate P-values that were significant after Bonferroni correction. COPD = chronic obstructive pulmonary disease.

Supplementary Table 61. Linear regressions of individuals’ probability scores for Atypical Symptoms on depression recurrence and on treatment resistant depression.

| Depression | Effect Size | Standard Error | *P*-value |
| --- | --- | --- | --- |
| Recurrent | 0.01790 | 0.00712 | 0.012 |
| Treatment resistance | 0.04230 | 0.02849 | 0.14 |

Recurrence and treatment resistance were both fitted as factors and were examined separately with age at questionnaire and sex fitted as covariates. Bold values indicate P-values that were significant after Bonferroni correction.

Supplementary Table 62. Linear regressions of individuals’ probability scores for Atypical Symptoms on polygenic scores for bipolar disorder, schizophrenia, and Attention-Deficit/Hyperactivity Disorder (ADHD).

| Mental health polygenic scores | Effect Size | Standard Error | *P*-value |
| --- | --- | --- | --- |
| Bipolar disorder | -0.00240 | 0.00250 | 0.34 |
| Schizophrenia | -0.00150 | 0.00242 | 0.54 |
| ADHD | 0.00410 | 0.00247 | 0.10 |

The polygenic scores for each trait were examined separately with age, sex, ancestry, and the first 10 genetic principal components fitted as covariates. Bold values indicate P-values that were significant after Bonferroni correction.

#### Hypersomnia, Weight Loss, and Mortality-Focus

Supplementary Table 63. Multivariable linear regression of individuals’ probability scores for Hypersomnia, Weight Loss, and Mortality-Focus on demographic variables.

| Demographic variables | Effect Size | Standard Error | *P*-value |
| --- | --- | --- | --- |
| Age | -0.00156 | 0.00031 | **3.36 × 10^-7^** |
| Sex | -0.01340 | 0.00486 | 0.0059 |
| Ethnicity - Asian | -0.02821 | 0.03146 | 0.37 |
| Ethnicity - Black | 0.02478 | 0.02702 | 0.36 |
| Ethnicity - Chinese | 0.00846 | 0.05835 | 0.88 |
| Ethnicity - Mixed | -0.01788 | 0.02584 | 0.49 |
| Ethnicity - Other | 0.02098 | 0.03089 | 0.50 |
| Place of Birth | -0.00827 | 0.00967 | 0.39 |
| Townsend Deprivation Index | 0.00319 | 0.00236 | 0.18 |
| Smoking - Former | -0.01814 | 0.00829 | 0.029 |
| Smoking - Never | -0.03103 | 0.00800 | **1.05 × 10^-4^** |
| Body Mass Index | -0.00615 | 0.00220 | 0.0051 |

Probability scores were calculated using Bernoulli-mixtures applied to ever depressed individuals at Q2. All variables were fitted simultaneously. Sex, ethnicity (European ethnicity as the reference), place of birth, and smoking (current smoking as the reference) were fitted as factors. Bold values indicate P-values that were significant after Bonferroni correction.

Supplementary Table 64. Multivariable linear regression of individuals’ probability scores for Hypersomnia, Weight Loss, and Mortality-Focus on health variables.

| Health variables | Effect Size | Standard Error | *P*-value |
| --- | --- | --- | --- |
| Myocardial infarction | 0.01466 | 0.01351 | 0.28 |
| Stroke | -0.01588 | 0.01830 | 0.39 |
| Asthma | 0.01004 | 0.00575 | 0.08 |
| COPD | 0.00821 | 0.01303 | 0.53 |
| Dementia | -0.06958 | 0.09431 | 0.46 |
| End stage renal disease | 0.20018 | 0.05609 | 3.60 × 10^-4^ |
| Motor neurone disease | 0.25007 | 0.13329 | 0.06 |
| Parkinson’s disease | -0.01989 | 0.04822 | 0.68 |

Probability scores were calculated using Bernoulli-mixtures applied to ever depressed individuals at Q2. All variables were fitted simultaneously with age at questionnaire and sex fitted as covariates. Bold values indicate P-values that were significant after Bonferroni correction. COPD = chronic obstructive pulmonary disease.

Supplementary Table 65. Linear regressions of individuals’ probability scores for Hypersomnia, Weight Loss, and Mortality-Focus on depression recurrence and on treatment resistant depression.

| Depression | Effect Size | Standard Error | *P*-value |
| --- | --- | --- | --- |
| Recurrent | 0.01866 | 0.00646 | 0.0039 |
| Treatment resistance | 0.03644 | 0.02590 | 0.16 |

Recurrence and treatment resistance were both fitted as factors and were examined separately with age at questionnaire and sex fitted as covariates. Bold values indicate P-values that were significant after Bonferroni correction.

Supplementary Table 66. Linear regressions of individuals’ probability scores for Hypersomnia, Weight Loss, and Mortality-Focus on polygenic scores for bipolar disorder, schizophrenia, and Attention-Deficit/Hyperactivity Disorder (ADHD).

| Mental health polygenic scores | Effect Size | Standard Error | *P*-value |
| --- | --- | --- | --- |
| Bipolar disorder | 0.00623 | 0.00225 | 0.0057 |
| Schizophrenia | 0.00005 | 0.00218 | 0.98 |
| ADHD | 0.00639 | 0.00223 | 0.0041 |

The polygenic scores for each trait were examined separately with age, sex, ancestry, and the first 10 genetic principal components fitted as covariates. Bold values indicate P-values that were significant after Bonferroni correction.

#### Weight Loss and Mortality-Focus

Supplementary Table 67. Multivariable linear regression of individuals’ probability scores for Weight Loss and Mortality-Focus on demographic variables.

| Demographic variables | Effect Size | Standard Error | *P*-value |
| --- | --- | --- | --- |
| Age | 0.00008 | 0.00008 | 0.31 |
| Sex | 0.00112 | 0.00122 | 0.36 |
| Ethnicity - Asian | 0.01372 | 0.00788 | 0.08 |
| Ethnicity - Black | 0.00397 | 0.00677 | 0.56 |
| Ethnicity - Chinese | 0.05346 | 0.01461 | 2.55 × 10^-4^ |
| Ethnicity - Mixed | -0.00425 | 0.00647 | 0.51 |
| Ethnicity - Other | -0.00605 | 0.00774 | 0.43 |
| Place of Birth | 0.00169 | 0.00242 | 0.48 |
| Townsend Deprivation Index | 0.00044 | 0.00059 | 0.45 |
| Smoking - Former | -0.00212 | 0.00208 | 0.31 |
| Smoking - Never | -0.00039 | 0.00200 | 0.85 |
| Body Mass Index | -0.00137 | 0.00055 | 0.013 |

Probability scores were calculated using Bernoulli-mixtures applied to ever depressed individuals at Q2. All variables were fitted simultaneously. Sex, ethnicity (European ethnicity as the reference), place of birth, and smoking (current smoking as the reference) were fitted as factors. Bold values indicate P-values that were significant after Bonferroni correction.

Supplementary Table 68. Multivariable linear regression of individuals’ probability scores for Weight Loss and Mortality-Focus on health variables.

| Health variables | Effect Size | Standard Error | *P*-value |
| --- | --- | --- | --- |
| Myocardial infarction | -0.00442 | 0.00341 | 0.20 |
| Stroke | -0.00569 | 0.00462 | 0.22 |
| Asthma | 0.00114 | 0.00145 | 0.43 |
| COPD | -0.00021 | 0.00329 | 0.95 |
| Dementia | -0.00695 | 0.02379 | 0.77 |
| End stage renal disease | 0.01203 | 0.01415 | 0.40 |
| Motor neurone disease | -0.00751 | 0.03363 | 0.82 |
| Parkinson’s disease | -0.00692 | 0.01217 | 0.57 |

Probability scores were calculated using Bernoulli-mixtures applied to ever depressed individuals at Q2. All variables were fitted simultaneously with age at questionnaire and sex fitted as covariates. Bold values indicate P-values that were significant after Bonferroni correction. COPD = chronic obstructive pulmonary disease.

Supplementary Table 69. Linear regressions of individuals’ probability scores for Weight Loss and Mortality-Focus on depression recurrence and on treatment resistant depression.

| Depression | Effect Size | Standard Error | *P*-value |
| --- | --- | --- | --- |
| Recurrent | -0.00189 | 0.00185 | 0.31 |
| Treatment resistance | 0.00500 | 0.00367 | 0.17 |

Recurrence and treatment resistance were both fitted as factors and were examined separately with age at questionnaire and sex fitted as covariates. Bold values indicate P-values that were significant after Bonferroni correction.

Supplementary Table 70. Linear regressions of individuals’ probability scores for Weight Loss and Mortality-Focus on polygenic scores for bipolar disorder, schizophrenia, and Attention-Deficit/Hyperactivity Disorder (ADHD).

| Mental health polygenic scores | Effect Size | Standard Error | *P*-value |
| --- | --- | --- | --- |
| Bipolar disorder | 0.00011 | 0.00056 | 0.85 |
| Schizophrenia | 0.00063 | 0.00054 | 0.25 |
| ADHD | 0.00006 | 0.00056 | 0.91 |

The polygenic scores for each trait were examined separately with age, sex, ancestry, and the first 10 genetic principal components fitted as covariates. Bold values indicate P-values that were significant after Bonferroni correction.

#### Prolonged Sleep Latency and Weight Loss

Supplementary Table 71. Multivariable linear regression of individuals’ probability scores for Prolonged Sleep Latency and Weight Loss on demographic variables.

| Demographic variables | Effect Size | Standard Error | *P*-value |
| --- | --- | --- | --- |
| Age | 0.00189 | 0.00026 | **4.64 × 10^-13^** |
| Sex | -0.01479 | 0.00415 | 3.67 × 10^-4^ |
| Ethnicity - Asian | -0.01108 | 0.02686 | 0.68 |
| Ethnicity - Black | 0.00192 | 0.02307 | 0.93 |
| Ethnicity - Chinese | -0.06071 | 0.04983 | 0.22 |
| Ethnicity - Mixed | -0.00751 | 0.02207 | 0.73 |
| Ethnicity - Other | -0.03589 | 0.02638 | 0.17 |
| Place of Birth | 0.00637 | 0.00826 | 0.44 |
| Townsend Deprivation Index | 0.00113 | 0.00201 | 0.57 |
| Smoking - Former | -0.01398 | 0.00708 | 0.048 |
| Smoking - Never | -0.00584 | 0.00683 | 0.39 |
| Body Mass Index | -0.00674 | 0.00187 | 3.26 × 10^-4^ |

Probability scores were calculated using Bernoulli-mixtures applied to ever depressed individuals at Q2. All variables were fitted simultaneously. Sex, ethnicity (European ethnicity as the reference), place of birth, and smoking (current smoking as the reference) were fitted as factors. Bold values indicate P-values that were significant after Bonferroni correction.

Supplementary Table 72. Multivariable linear regression of individuals’ probability scores for Prolonged Sleep Latency and Weight Loss on health variables.

| Health variables | Effect Size | Standard Error | *P*-value |
| --- | --- | --- | --- |
| Myocardial infarction | -0.00809 | 0.01145 | 0.48 |
| Stroke | 0.00370 | 0.01550 | 0.81 |
| Asthma | -0.00610 | 0.00487 | 0.21 |
| COPD | -0.00003 | 0.01104 | 1.00 |
| Dementia | -0.04843 | 0.07989 | 0.54 |
| End stage renal disease | 0.07091 | 0.04751 | 0.14 |
| Motor neurone disease | -0.04850 | 0.11290 | 0.67 |
| Parkinson’s disease | -0.02850 | 0.04085 | 0.49 |

Probability scores were calculated using Bernoulli-mixtures applied to ever depressed individuals at Q2. All variables were fitted simultaneously with age at questionnaire and sex fitted as covariates. Bold values indicate P-values that were significant after Bonferroni correction. COPD = chronic obstructive pulmonary disease.

Supplementary Table 73. Linear regressions of individuals’ probability scores for Prolonged Sleep Latency and Weight Loss on depression recurrence and on treatment resistant depression.

| Depression | Effect Size | Standard Error | *P*-value |
| --- | --- | --- | --- |
| Recurrent | -0.02197 | 0.00540 | **4.84 × 10^-5^** |
| Treatment resistance | -0.00783 | 0.01590 | 0.62 |

Recurrence and treatment resistance were both fitted as factors and were examined separately with age at questionnaire and sex fitted as covariates. Bold values indicate P-values that were significant after Bonferroni correction.

Supplementary Table 74. Linear regressions of individuals’ probability scores for Prolonged Sleep Latency and Weight Loss on polygenic scores for bipolar disorder, schizophrenia, and Attention-Deficit/Hyperactivity Disorder (ADHD).

| Mental health polygenic scores | Effect Size | Standard Error | *P*-value |
| --- | --- | --- | --- |
| Bipolar disorder | -0.00129 | 0.00189 | 0.50 |
| Schizophrenia | 0.00026 | 0.00184 | 0.89 |
| ADHD | -0.00132 | 0.00187 | 0.48 |

The polygenic scores for each trait were examined separately with age, sex, ancestry, and the first 10 genetic principal components fitted as covariates. Bold values indicate P-values that were significant after Bonferroni correction.

#### Terminal Insomnia

Supplementary Table 75. Multivariable linear regression of individuals’ probability scores for Terminal Insomnia on demographic variables.

| Demographic variables | Effect Size | Standard Error | *P*-value |
| --- | --- | --- | --- |
| Age | -0.00031 | 0.00025 | 0.21 |
| Sex | 0.02524 | 0.00402 | **3.62 × 10^-10^** |
| Ethnicity - Asian | -0.01462 | 0.02603 | 0.57 |
| Ethnicity - Black | -0.03817 | 0.02236 | 0.09 |
| Ethnicity - Chinese | -0.05959 | 0.04828 | 0.22 |
| Ethnicity - Mixed | -0.01950 | 0.02138 | 0.36 |
| Ethnicity - Other | 0.00348 | 0.02556 | 0.89 |
| Place of Birth | 0.00867 | 0.00801 | 0.28 |
| Townsend Deprivation Index | -0.00183 | 0.00195 | 0.35 |
| Smoking - Former | 0.01657 | 0.00686 | 0.016 |
| Smoking - Never | 0.01900 | 0.00662 | 0.0041 |
| Body Mass Index | 0.00470 | 0.00182 | 0.010 |

Probability scores were calculated using Bernoulli-mixtures applied to ever depressed individuals at Q2. All variables were fitted simultaneously. Sex, ethnicity (European ethnicity as the reference), place of birth, and smoking (current smoking as the reference) were fitted as factors. Bold values indicate P-values that were significant after Bonferroni correction.

Supplementary Table 76. Multivariable linear regression of individuals’ probability scores for Terminal Insomnia on health variables.

| Health variables | Effect Size | Standard Error | *P*-value |
| --- | --- | --- | --- |
| Myocardial infarction | -0.01302 | 0.01118 | 0.24 |
| Stroke | 0.01961 | 0.01513 | 0.20 |
| Asthma | -0.00152 | 0.00475 | 0.75 |
| COPD | -0.01942 | 0.01078 | 0.07 |
| Dementia | 0.03337 | 0.07801 | 0.67 |
| End stage renal disease | -0.06379 | 0.04639 | 0.17 |
| Motor neurone disease | -0.05880 | 0.11026 | 0.59 |
| Parkinson’s disease | -0.01207 | 0.03989 | 0.76 |

Probability scores were calculated using Bernoulli-mixtures applied to ever depressed individuals at Q2. All variables were fitted simultaneously with age at questionnaire and sex fitted as covariates. Bold values indicate P-values that were significant after Bonferroni correction. COPD = chronic obstructive pulmonary disease.

Supplementary Table 77. Linear regressions of individuals’ probability scores for Terminal Insomnia on depression recurrence and on treatment resistant depression.

| Depression | Effect Size | Standard Error | *P*-value |
| --- | --- | --- | --- |
| Recurrent | 0.00956 | 0.00547 | 0.08 |
| Treatment resistance | -0.00975 | 0.01857 | 0.60 |

Recurrence and treatment resistance were both fitted as factors and were examined separately with age at questionnaire and sex fitted as covariates. Bold values indicate P-values that were significant after Bonferroni correction.

Supplementary Table 78. Linear regressions of individuals’ probability scores for Terminal Insomnia on polygenic scores for bipolar disorder, schizophrenia, and Attention-Deficit/Hyperactivity Disorder (ADHD).

| Mental health polygenic scores | Effect Size | Standard Error | *P*-value |
| --- | --- | --- | --- |
| Bipolar disorder | -0.00072 | 0.00186 | 0.70 |
| Schizophrenia | 0.00057 | 0.00180 | 0.75 |
| ADHD | -0.00330 | 0.00184 | 0.07 |

The polygenic scores for each trait were examined separately with age, sex, ancestry, and the first 10 genetic principal components fitted as covariates. Bold values indicate P-values that were significant after Bonferroni correction.

#### Insomnia, Weight Gain and Mortality-Focus

Supplementary Table 79. Multivariable linear regression of individuals’ probability scores for Insomnia, Weight Gain and Mortality-Focus on demographic variables.

| Demographic variables | Effect Size | Standard Error | *P*-value |
| --- | --- | --- | --- |
| Age | -0.00253 | 0.00026 | **<2.00 × 10^-16^** |
| Sex | -0.01846 | 0.00415 | **8.81 × 10^-6^** |
| Ethnicity - Asian | 0.01785 | 0.02687 | 0.51 |
| Ethnicity - Black | -0.00366 | 0.02308 | 0.87 |
| Ethnicity - Chinese | -0.01414 | 0.04984 | 0.78 |
| Ethnicity - Mixed | 0.04069 | 0.02207 | 0.07 |
| Ethnicity - Other | 0.03448 | 0.02639 | 0.19 |
| Place of Birth | 0.01230 | 0.00826 | 0.14 |
| Townsend Deprivation Index | 0.00033 | 0.00201 | 0.87 |
| Smoking - Former | -0.00109 | 0.00708 | 0.88 |
| Smoking - Never | 0.00033 | 0.00683 | 0.96 |
| Body Mass Index | 0.04221 | 0.00188 | **<2.00 × 10^-16^** |

Probability scores were calculated using Bernoulli-mixtures applied to ever depressed individuals at Q2. All variables were fitted simultaneously. Sex, ethnicity (European ethnicity as the reference), place of birth, and smoking (current smoking as the reference) were fitted as factors. Bold values indicate P-values that were significant after Bonferroni correction.

Supplementary Table 80. Multivariable linear regression of individuals’ probability scores for Insomnia, Weight Gain and Mortality-Focus on health variables.

| Health variables | Effect Size | Standard Error | *P*-value |
| --- | --- | --- | --- |
| Myocardial infarction | 0.03888 | 0.01191 | 0.0011 |
| Stroke | 0.04322 | 0.01612 | 0.0074 |
| Asthma | 0.03017 | 0.00506 | **2.64 × 10^-9^** |
| COPD | 0.01025 | 0.01149 | 0.37 |
| Dementia | 0.03264 | 0.08310 | 0.69 |
| End stage renal disease | -0.04930 | 0.04942 | 0.32 |
| Motor neurone disease | 0.00044 | 0.11746 | 1.00 |
| Parkinson’s disease | -0.00991 | 0.04249 | 0.82 |

Probability scores were calculated using Bernoulli-mixtures applied to ever depressed individuals at Q2. All variables were fitted simultaneously with age at questionnaire and sex fitted as covariates. Bold values indicate P-values that were significant after Bonferroni correction. COPD = chronic obstructive pulmonary disease.

Supplementary Table 81. Linear regressions of individuals’ probability scores for Insomnia, Weight Gain and Mortality-Focus on depression recurrence and on treatment resistant depression.

| Depression | Effect Size | Standard Error | *P*-value |
| --- | --- | --- | --- |
| Recurrent | 0.04226 | 0.00528 | **1.49 × 10^-15^** |
| Treatment resistance | 0.06992 | 0.02214 | 0.0016 |

Recurrence and treatment resistance were both fitted as factors and were examined separately with age at questionnaire and sex fitted as covariates. Bold values indicate P-values that were significant after Bonferroni correction.

Supplementary Table 82. Linear regressions of individuals’ probability scores for Insomnia, Weight Gain and Mortality-Focus on polygenic scores for bipolar disorder, schizophrenia, and Attention-Deficit/Hyperactivity Disorder (ADHD).

| Mental health polygenic scores | Effect Size | Standard Error | *P*-value |
| --- | --- | --- | --- |
| Bipolar disorder | 0.00073 | 0.00197 | 0.71 |
| Schizophrenia | -0.00107 | 0.00191 | 0.58 |
| ADHD | 0.00821 | 0.00195 | **2.52 × 10^-5^** |

The polygenic scores for each trait were examined separately with age, sex, ancestry, and the first 10 genetic principal components fitted as covariates. Bold values indicate P-values that were significant after Bonferroni correction.

#### Insomnia and Weight Gain

Supplementary Table 83. Multivariable linear regression of individuals’ probability scores for Insomnia and Weight Gain on demographic variables.

| Demographic variables | Effect Size | Standard Error | *P*-value |
| --- | --- | --- | --- |
| Age | -0.00051 | 0.00016 | 0.0016 |
| Sex | -0.00508 | 0.00255 | 0.046 |
| Ethnicity - Asian | 0.00728 | 0.01651 | 0.66 |
| Ethnicity - Black | 0.00081 | 0.01418 | 0.95 |
| Ethnicity - Chinese | -0.00249 | 0.03063 | 0.94 |
| Ethnicity - Mixed | 0.01344 | 0.01356 | 0.32 |
| Ethnicity - Other | -0.02629 | 0.01622 | 0.11 |
| Place of Birth | 0.00662 | 0.00508 | 0.19 |
| Townsend Deprivation Index | 0.00059 | 0.00124 | 0.63 |
| Smoking - Former | 0.00734 | 0.00435 | 0.09 |
| Smoking - Never | 0.01133 | 0.00420 | 0.0070 |
| Body Mass Index | 0.01580 | 0.00115 | **<2.00 × 10^-16^** |

Probability scores were calculated using Bernoulli-mixtures applied to ever depressed individuals at Q2. All variables were fitted simultaneously. Sex, ethnicity (European ethnicity as the reference), place of birth, and smoking (current smoking as the reference) were fitted as factors. Bold values indicate P-values that were significant after Bonferroni correction.

Supplementary Table 84. Multivariable linear regression of individuals’ probability scores for Insomnia and Weight Gain on health variables.

| Health variables | Effect Size | Standard Error | *P*-value |
| --- | --- | --- | --- |
| Myocardial infarction | 0.01619 | 0.00971 | 0.10 |
| Stroke | 0.00633 | 0.00305 | 0.038 |
| Asthma | -0.01600 | 0.00691 | 0.021 |
| COPD | -0.02454 | 0.05003 | 0.62 |
| Dementia | 0.06440 | 0.02976 | 0.030 |
| End stage renal disease | -0.02789 | 0.07072 | 0.69 |
| Motor neurone disease | 0.00288 | 0.02558 | 0.91 |
| Parkinson’s disease | 0.01619 | 0.00971 | 0.10 |

Probability scores were calculated using Bernoulli-mixtures applied to ever depressed individuals at Q2. All variables were fitted simultaneously with age at questionnaire and sex fitted as covariates. Bold values indicate P-values that were significant after Bonferroni correction. COPD = chronic obstructive pulmonary disease.

Supplementary Table 85. Linear regressions of individuals’ probability scores for Insomnia and Weight Gain on depression recurrence and on treatment resistant depression.

| Depression | Effect Size | Standard Error | *P*-value |
| --- | --- | --- | --- |
| Recurrent | 0.00003 | 0.00327 | 0.99 |
| Treatment resistance | 0.01834 | 0.00886 | 0.039 |

Recurrence and treatment resistance were both fitted as factors and were examined separately with age at questionnaire and sex fitted as covariates. Bold values indicate P-values that were significant after Bonferroni correction.

Supplementary Table 86. Linear regressions of individuals’ probability scores for Insomnia and Weight Gain on polygenic scores for bipolar disorder, schizophrenia, and Attention-Deficit/Hyperactivity Disorder (ADHD).

| Mental health polygenic scores | Effect Size | Standard Error | *P*-value |
| --- | --- | --- | --- |
| Bipolar disorder | -0.00063 | 0.00119 | 0.59 |
| Schizophrenia | -0.00193 | 0.00115 | 0.09 |
| ADHD | 0.00098 | 0.00117 | 0.41 |

The polygenic scores for each trait were examined separately with age, sex, ancestry, and the first 10 genetic principal components fitted as covariates. Bold values indicate P-values that were significant after Bonferroni correction.

#### Insomnia without Weight Change

Supplementary Table 87. Multivariable linear regression of individuals’ probability scores for Insomnia without Weight Change on demographic variables.

| Demographic variables | Effect Size | Standard Error | *P*-value |
| --- | --- | --- | --- |
| Age | 0.00111 | 0.00018 | **1.40 × 10^-9^** |
| Sex | 0.01746 | 0.00291 | **1.92 × 10^-9^** |
| Ethnicity - Asian | 0.00443 | 0.01880 | 0.81 |
| Ethnicity - Black | 0.00565 | 0.01615 | 0.73 |
| Ethnicity - Chinese | 0.06962 | 0.03488 | 0.046 |
| Ethnicity - Mixed | -0.01511 | 0.01544 | 0.33 |
| Ethnicity - Other | 0.00694 | 0.01847 | 0.71 |
| Place of Birth | -0.00991 | 0.00578 | 0.09 |
| Townsend Deprivation Index | 0.00002 | 0.00141 | 0.99 |
| Smoking - Former | 0.00777 | 0.00495 | 0.12 |
| Smoking - Never | 0.01130 | 0.00478 | 0.018 |
| Body Mass Index | -0.00400 | 0.00131 | 0.0023 |

Probability scores were calculated using Bernoulli-mixtures applied to ever depressed individuals at Q2. All variables were fitted simultaneously. Sex, ethnicity (European ethnicity as the reference), place of birth, and smoking (current smoking as the reference) were fitted as factors. Bold values indicate P-values that were significant after Bonferroni correction.

Supplementary Table 88. Multivariable linear regression of individuals’ probability scores for Insomnia without Weight Change on health variables.

| Health variables | Effect Size | Standard Error | *P*-value |
| --- | --- | --- | --- |
| Myocardial infarction | -0.00265 | 0.00809 | 0.74 |
| Stroke | -0.00128 | 0.01096 | 0.91 |
| Asthma | -0.00329 | 0.00344 | 0.34 |
| COPD | -0.00754 | 0.00781 | 0.33 |
| Dementia | -0.02796 | 0.05649 | 0.62 |
| End stage renal disease | 0.02187 | 0.03360 | 0.52 |
| Motor neurone disease | -0.02435 | 0.07984 | 0.76 |
| Parkinson’s disease | 0.01103 | 0.02889 | 0.70 |

Probability scores were calculated using Bernoulli-mixtures applied to ever depressed individuals at Q2. All variables were fitted simultaneously with age at questionnaire and sex fitted as covariates. Bold values indicate P-values that were significant after Bonferroni correction. COPD = chronic obstructive pulmonary disease.

Supplementary Table 89. Linear regressions of individuals’ probability scores for Insomnia without Weight Change on depression recurrence and on treatment resistant depression.

| Depression | Effect Size | Standard Error | *P*-value |
| --- | --- | --- | --- |
| Recurrent | -0.00568 | 0.00403 | 0.16 |
| Treatment resistance | -0.01835 | 0.01151 | 0.11 |

Recurrence and treatment resistance were both fitted as factors and were examined separately with age at questionnaire and sex fitted as covariates. Bold values indicate P-values that were significant after Bonferroni correction.

Supplementary Table 90. Linear regressions of individuals’ probability scores for Insomnia without Weight Change on polygenic scores for bipolar disorder, schizophrenia, and Attention-Deficit/Hyperactivity Disorder (ADHD).

| Mental health polygenic scores | Effect Size | Standard Error | *P*-value |
| --- | --- | --- | --- |
| Bipolar disorder | -0.00351 | 0.00135 | 0.0093 |
| Schizophrenia | -0.00210 | 0.00131 | 0.11 |
| ADHD | -0.00242 | 0.00134 | 0.07 |

The polygenic scores for each trait were examined separately with age, sex, ancestry, and the first 10 genetic principal components fitted as covariates. Bold values indicate P-values that were significant after Bonferroni correction.

#### Melancholia with Low Self-worth and Mortality-Focus

Supplementary Table 91. Multivariable linear regression of individuals’ probability scores for Melancholia with Low Self-worth and Mortality-Focus on demographic variables.

| Demographic variables | Effect Size | Standard Error | *P*-value |
| --- | --- | --- | --- |
| Age | 0.00005 | 0.00025 | 0.86 |
| Sex | -0.00335 | 0.00400 | 0.40 |
| Ethnicity - Asian | -0.02065 | 0.02586 | 0.42 |
| Ethnicity - Black | -0.02473 | 0.02221 | 0.27 |
| Ethnicity - Chinese | -0.02175 | 0.04797 | 0.65 |
| Ethnicity - Mixed | 0.00300 | 0.02124 | 0.89 |
| Ethnicity - Other | -0.00431 | 0.02540 | 0.87 |
| Place of Birth | -0.01467 | 0.00795 | 0.07 |
| Townsend Deprivation Index | 0.00528 | 0.00194 | 0.0064 |
| Smoking - Former | -0.02167 | 0.00681 | 0.0015 |
| Smoking - Never | -0.02777 | 0.00658 | **2.44 × 10^-5^** |
| Body Mass Index | -0.01339 | 0.00181 | **1.26 × 10^-13^** |

Probability scores were calculated using Bernoulli-mixtures applied to ever depressed individuals at Q2. All variables were fitted simultaneously. Sex, ethnicity (European ethnicity as the reference), place of birth, and smoking (current smoking as the reference) were fitted as factors. Bold values indicate P-values that were significant after Bonferroni correction.

Supplementary Table 92. Multivariable linear regression of individuals’ probability scores for Melancholia with Low Self-worth and Mortality-Focus on health variables.

| Health variables | Effect Size | Standard Error | *P*-value |
| --- | --- | --- | --- |
| Myocardial infarction | 0.00022 | 0.01114 | 0.98 |
| Stroke | -0.01120 | 0.01508 | 0.46 |
| Asthma | -0.00348 | 0.00474 | 0.46 |
| COPD | 0.02223 | 0.01074 | 0.039 |
| Dementia | 0.09259 | 0.07774 | 0.23 |
| End stage renal disease | 0.04056 | 0.04623 | 0.38 |
| Motor neurone disease | 0.08190 | 0.10988 | 0.46 |
| Parkinson’s disease | 0.02946 | 0.03975 | 0.46 |

Probability scores were calculated using Bernoulli-mixtures applied to ever depressed individuals at Q2. All variables were fitted simultaneously with age at questionnaire and sex fitted as covariates. Bold values indicate P-values that were significant after Bonferroni correction. COPD = chronic obstructive pulmonary disease.

Supplementary Table 93. Linear regressions of individuals’ probability scores for Melancholia with Low Self-worth and Mortality-Focus on depression recurrence and on treatment resistant depression.

| Depression | Effect Size | Standard Error | *P*-value |
| --- | --- | --- | --- |
| Recurrent | 0.01397 | 0.00517 | 0.0069 |
| Treatment resistance | -0.00357 | 0.02020 | 0.86 |

Recurrence and treatment resistance were both fitted as factors and were examined separately with age at questionnaire and sex fitted as covariates. Bold values indicate P-values that were significant after Bonferroni correction.

Supplementary Table 94. Linear regressions of individuals’ probability scores for Melancholia with Low Self-worth and Mortality-Focus on polygenic scores for bipolar disorder, schizophrenia, and Attention-Deficit/Hyperactivity Disorder (ADHD).

| Mental health polygenic scores | Effect Size | Standard Error | *P*-value |
| --- | --- | --- | --- |
| Bipolar disorder | 0.00377 | 0.00184 | 0.041 |
| Schizophrenia | 0.00402 | 0.00179 | 0.024 |
| ADHD | 0.00239 | 0.00182 | 0.19 |

The polygenic scores for each trait were examined separately with age, sex, ancestry, and the first 10 genetic principal components fitted as covariates. Bold values indicate P-values that were significant after Bonferroni correction.

#### Melancholia

Supplementary Table 95. Multivariable linear regression of individuals’ probability scores for Melancholia on demographic variables.

| Demographic variables | Effect Size | Standard Error | *P*-value |
| --- | --- | --- | --- |
| Age | 0.00256 | 0.00039 | **7.32 × 10^-11^** |
| Sex | -0.04360 | 0.00625 | **3.18 × 10^-12^** |
| Ethnicity - Asian | 0.05783 | 0.04045 | 0.15 |
| Ethnicity - Black | 0.02938 | 0.03474 | 0.40 |
| Ethnicity - Chinese | -0.03137 | 0.07503 | 0.68 |
| Ethnicity - Mixed | 0.02506 | 0.03323 | 0.45 |
| Ethnicity - Other | 0.00578 | 0.03972 | 0.88 |
| Place of Birth | -0.01612 | 0.01244 | 0.20 |
| Townsend Deprivation Index | -0.00594 | 0.00303 | 0.050 |
| Smoking - Former | 0.01180 | 0.01066 | 0.27 |
| Smoking - Never | 0.01110 | 0.01029 | 0.28 |
| Body Mass Index | -0.03741 | 0.00282 | **<2.00 × 10^-16^** |

Probability scores were calculated using Bernoulli-mixtures applied to ever depressed individuals at Q2. All variables were fitted simultaneously. Sex, ethnicity (European ethnicity as the reference), place of birth, and smoking (current smoking as the reference) were fitted as factors. Bold values indicate P-values that were significant after Bonferroni correction.

Supplementary Table 96. Multivariable linear regression of individuals’ probability scores for Melancholia on health variables.

| Health variables | Effect Size | Standard Error | *P*-value |
| --- | --- | --- | --- |
| Myocardial infarction | -0.00976 | 0.01747 | 0.58 |
| Stroke | -0.04425 | 0.02366 | 0.06 |
| Asthma | -0.02417 | 0.00743 | 0.0012 |
| COPD | 0.00443 | 0.01685 | 0.79 |
| Dementia | -0.02699 | 0.12195 | 0.82 |
| End stage renal disease | -0.08434 | 0.07252 | 0.24 |
| Motor neurone disease | -0.06348 | 0.17236 | 0.71 |
| Parkinson’s disease | 0.00165 | 0.06236 | 0.98 |

Probability scores were calculated using Bernoulli-mixtures applied to ever depressed individuals at Q2. All variables were fitted simultaneously with age at questionnaire and sex fitted as covariates. Bold values indicate P-values that were significant after Bonferroni correction. COPD = chronic obstructive pulmonary disease.

Supplementary Table 97. Linear regressions of individuals’ probability scores for Melancholia on depression recurrence and on treatment resistant depression.

| Depression | Effect Size | Standard Error | *P*-value |
| --- | --- | --- | --- |
| Recurrent | -0.05268 | 0.00822 | **1.59 × 10^-10^** |
| Treatment resistance | -0.10023 | 0.02486 | **5.75 × 10^-5^** |

Recurrence and treatment resistance were both fitted as factors and were examined separately with age at questionnaire and sex fitted as covariates. Bold values indicate P-values that were significant after Bonferroni correction.

Supplementary Table 98. Linear regressions of individuals’ probability scores for Melancholia on polygenic scores for bipolar disorder, schizophrenia, and Attention-Deficit/Hyperactivity Disorder (ADHD).

| Mental health polygenic scores | Effect Size | Standard Error | *P*-value |
| --- | --- | --- | --- |
| Bipolar disorder | -0.00074 | 0.00290 | 0.80 |
| Schizophrenia | 0.00147 | 0.00281 | 0.60 |
| ADHD | -0.00809 | 0.00287 | 0.0048 |

The polygenic scores for each trait were examined separately with age, sex, ancestry, and the first 10 genetic principal components fitted as covariates. Bold values indicate P-values that were significant after Bonferroni correction.

#### Melancholia with Low Self-worth

Supplementary Table 99. Multivariable linear regression of individuals’ probability scores for Melancholia with Low Self-worth on demographic variables.

| Demographic variables | Effect Size | Standard Error | *P*-value |
| --- | --- | --- | --- |
| Age | 0.00012 | 0.00023 | 0.61 |
| Sex | -0.01709 | 0.00358 | **1.84 × 10^-6^** |
| Ethnicity - Asian | -0.00787 | 0.02318 | 0.73 |
| Ethnicity - Black | -0.01024 | 0.01991 | 0.61 |
| Ethnicity - Chinese | -0.01840 | 0.04299 | 0.67 |
| Ethnicity - Mixed | -0.00883 | 0.01904 | 0.64 |
| Ethnicity - Other | 0.01431 | 0.02276 | 0.53 |
| Place of Birth | -0.01187 | 0.00713 | 0.10 |
| Townsend Deprivation Index | -0.00725 | 0.00174 | **2.97 × 10^-5^** |
| Smoking - Former | -0.00128 | 0.00611 | 0.83 |
| Smoking - Never | -0.00026 | 0.00589 | 0.97 |
| Body Mass Index | -0.02262 | 0.00162 | **<2.00 × 10^-16^** |

Probability scores were calculated using Bernoulli-mixtures applied to ever depressed individuals at Q2. All variables were fitted simultaneously. Sex, ethnicity (European ethnicity as the reference), place of birth, and smoking (current smoking as the reference) were fitted as factors. Bold values indicate P-values that were significant after Bonferroni correction.

Supplementary Table 100. Multivariable linear regression of individuals’ probability scores for Melancholia with Low Self-worth on health variables.

| Health variables | Effect Size | Standard Error | *P*-value |
| --- | --- | --- | --- |
| Myocardial infarction | -0.00654 | 0.00999 | 0.51 |
| Stroke | -0.00049 | 0.01353 | 0.97 |
| Asthma | -0.01001 | 0.00425 | 0.019 |
| COPD | 0.00903 | 0.00964 | 0.35 |
| Dementia | 0.08764 | 0.06973 | 0.21 |
| End stage renal disease | -0.06029 | 0.04147 | 0.15 |
| Motor neurone disease | -0.11250 | 0.09856 | 0.25 |
| Parkinson’s disease | -0.00202 | 0.03566 | 0.95 |

Probability scores were calculated using Bernoulli-mixtures applied to ever depressed individuals at Q2. All variables were fitted simultaneously with age at questionnaire and sex fitted as covariates. Bold values indicate P-values that were significant after Bonferroni correction. COPD = chronic obstructive pulmonary disease.

Supplementary Table 101. Linear regressions of individuals’ probability scores for Melancholia with Low Self-worth on depression recurrence and on treatment resistant depression.

| Depression | Effect Size | Standard Error | *P*-value |
| --- | --- | --- | --- |
| Recurrent | -0.00214 | 0.00471 | 0.65 |
| Treatment resistance | -0.04675 | 0.01667 | 0.0051 |

Recurrence and treatment resistance were both fitted as factors and were examined separately with age at questionnaire and sex fitted as covariates. Bold values indicate P-values that were significant after Bonferroni correction.

Supplementary Table 102. Linear regressions of individuals’ probability scores for Melancholia with Low Self-worth on polygenic scores for bipolar disorder, schizophrenia, and Attention-Deficit/Hyperactivity Disorder (ADHD).

| Mental health polygenic scores | Effect Size | Standard Error | *P*-value |
| --- | --- | --- | --- |
| Bipolar disorder | 0.00202 | 0.00166 | 0.22 |
| Schizophrenia | 0.00234 | 0.00161 | 0.15 |
| ADHD | -0.00180 | 0.00164 | 0.27 |

The polygenic scores for each trait were examined separately with age, sex, ancestry, and the first 10 genetic principal components fitted as covariates. Bold values indicate P-values that were significant after Bonferroni correction.

#### Insomnia and Low Self-worth

Supplementary Table 103. Multivariable linear regression of individuals’ probability scores for Insomnia and Low Self-worth on demographic variables.

| Demographic variables | Effect Size | Standard Error | *P*-value |
| --- | --- | --- | --- |
| Age | 0.00020 | 0.00025 | 0.42 |
| Sex | 0.03143 | 0.00396 | **2.23 × 10^-15^** |
| Ethnicity - Asian | -0.01210 | 0.02563 | 0.64 |
| Ethnicity - Black | 0.01459 | 0.02201 | 0.51 |
| Ethnicity - Chinese | 0.04989 | 0.04754 | 0.29 |
| Ethnicity - Mixed | 0.02007 | 0.02105 | 0.34 |
| Ethnicity - Other | 0.03442 | 0.02517 | 0.17 |
| Place of Birth | -0.00492 | 0.00788 | 0.53 |
| Townsend Deprivation Index | -0.00377 | 0.00192 | 0.050 |
| Smoking - Former | 0.00315 | 0.00675 | 0.64 |
| Smoking - Never | 0.00903 | 0.00652 | 0.17 |
| Body Mass Index | -0.00307 | 0.00179 | 0.09 |

Probability scores were calculated using Bernoulli-mixtures applied to ever depressed individuals at Q2. All variables were fitted simultaneously. Sex, ethnicity (European ethnicity as the reference), place of birth, and smoking (current smoking as the reference) were fitted as factors. Bold values indicate P-values that were significant after Bonferroni correction.

Supplementary Table 104. Multivariable linear regression of individuals’ probability scores for Insomnia and Low Self-worth on health variables.

| Health variables | Effect Size | Standard Error | *P*-value |
| --- | --- | --- | --- |
| Myocardial infarction | 0.00191 | 0.01096 | 0.86 |
| Stroke | 0.00425 | 0.01484 | 0.77 |
| Asthma | 0.00108 | 0.00466 | 0.82 |
| COPD | -0.02624 | 0.01057 | 0.013 |
| Dementia | -0.05390 | 0.07649 | 0.48 |
| End stage renal disease | -0.01305 | 0.04549 | 0.77 |
| Motor neurone disease | -0.05224 | 0.10810 | 0.63 |
| Parkinson’s disease | -0.03638 | 0.03911 | 0.35 |

Probability scores were calculated using Bernoulli-mixtures applied to ever depressed individuals at Q2. All variables were fitted simultaneously with age at questionnaire and sex fitted as covariates. Bold values indicate P-values that were significant after Bonferroni correction. COPD = chronic obstructive pulmonary disease.

Supplementary Table 105. Linear regressions of individuals’ probability scores for Insomnia and Low Self-worth on depression recurrence and on treatment resistant depression.

| Depression | Effect Size | Standard Error | *P*-value |
| --- | --- | --- | --- |
| Recurrent | -0.01012 | 0.00528 | 0.06 |
| Treatment resistance | 0.00467 | 0.01646 | 0.78 |

Recurrence and treatment resistance were both fitted as factors and were examined separately with age at questionnaire and sex fitted as covariates. Bold values indicate P-values that were significant after Bonferroni correction.

Supplementary Table 106. Linear regressions of individuals’ probability scores for Insomnia and Low Self-worth on polygenic scores for bipolar disorder, schizophrenia, and Attention-Deficit/Hyperactivity Disorder (ADHD).

| Mental health polygenic scores | Effect Size | Standard Error | *P*-value |
| --- | --- | --- | --- |
| Bipolar disorder | -0.00077 | 0.00180 | 0.67 |
| Schizophrenia | -0.00051 | 0.00175 | 0.77 |
| ADHD | -0.00551 | 0.00179 | 0.0020 |

The polygenic scores for each trait were examined separately with age, sex, ancestry, and the first 10 genetic principal components fitted as covariates. Bold values indicate P-values that were significant after Bonferroni correction.

#### Insomnia and Mortality-Focus

Supplementary Table 107. Multivariable linear regression of individuals’ probability scores for Insomnia and Mortality-Focus on demographic variables.

| Demographic variables | Effect Size | Standard Error | *P*-value |
| --- | --- | --- | --- |
| Age | 0.00126 | 0.00031 | **4.85 × 10^-5^** |
| Sex | 0.03461 | 0.00494 | **2.68 × 10^-12^** |
| Ethnicity - Asian | 0.02265 | 0.03199 | 0.48 |
| Ethnicity - Black | -0.00711 | 0.02748 | 0.80 |
| Ethnicity - Chinese | 0.03871 | 0.05934 | 0.51 |
| Ethnicity - Mixed | -0.03021 | 0.02628 | 0.25 |
| Ethnicity - Other | -0.04250 | 0.03142 | 0.18 |
| Place of Birth | 0.00676 | 0.00984 | 0.49 |
| Townsend Deprivation Index | 0.00062 | 0.00240 | 0.80 |
| Smoking - Former | 0.01382 | 0.00843 | 0.10 |
| Smoking - Never | 0.01562 | 0.00814 | 0.055 |
| Body Mass Index | 0.00844 | 0.00223 | 1.59 × 10^-4^ |

Probability scores were calculated using Bernoulli-mixtures applied to ever depressed individuals at Q2. All variables were fitted simultaneously. Sex, ethnicity (European ethnicity as the reference), place of birth, and smoking (current smoking as the reference) were fitted as factors. Bold values indicate P-values that were significant after Bonferroni correction.

Supplementary Table 108. Multivariable linear regression of individuals’ probability scores for Insomnia and Mortality-Focus on health variables.

| Health variables | Effect Size | Standard Error | *P*-value |
| --- | --- | --- | --- |
| Myocardial infarction | -0.01205 | 0.01377 | 0.38 |
| Stroke | -0.01543 | 0.01864 | 0.41 |
| Asthma | -0.00199 | 0.00586 | 0.73 |
| COPD | -0.01461 | 0.01328 | 0.27 |
| Dementia | -0.01661 | 0.09609 | 0.86 |
| End stage renal disease | -0.04975 | 0.05715 | 0.38 |
| Motor neurone disease | -0.09317 | 0.13582 | 0.49 |
| Parkinson’s disease | -0.03803 | 0.04914 | 0.44 |

Probability scores were calculated using Bernoulli-mixtures applied to ever depressed individuals at Q2. All variables were fitted simultaneously with age at questionnaire and sex fitted as covariates. Bold values indicate P-values that were significant after Bonferroni correction. COPD = chronic obstructive pulmonary disease.

Supplementary Table 109. Linear regressions of individuals’ probability scores for Insomnia and Mortality-Focus on depression recurrence and on treatment resistant depression.

| Depression | Effect Size | Standard Error | *P*-value |
| --- | --- | --- | --- |
| Recurrent | -0.00790 | 0.00661 | 0.23 |
| Treatment resistance | 0.00981 | 0.02149 | 0.65 |

Recurrence and treatment resistance were both fitted as factors and were examined separately with age at questionnaire and sex fitted as covariates. Bold values indicate P-values that were significant after Bonferroni correction.

Supplementary Table 110. Linear regressions of individuals’ probability scores for Insomnia and Mortality-Focus on polygenic scores for bipolar disorder, schizophrenia, and Attention-Deficit/Hyperactivity Disorder (ADHD).

| Mental health polygenic scores | Effect Size | Standard Error | *P*-value |
| --- | --- | --- | --- |
| Bipolar disorder | -0.00280 | 0.00230 | 0.22 |
| Schizophrenia | -0.00225 | 0.00223 | 0.31 |
| ADHD | 0.00033 | 0.00227 | 0.89 |

The polygenic scores for each trait were examined separately with age, sex, ancestry, and the first 10 genetic principal components fitted as covariates. Bold values indicate P-values that were significant after Bonferroni correction.

### Q1 (Mental Health Questionnaire) - *Currently depressed*

Number of individuals analysed: 3,096

Number of clusters identified: 11

#### Attentive and Appetite Disruption

Supplementary Table 111. Multivariable linear regression of individuals’ probability scores for Attentive and Appetite Disruption on demographic variables.

| Demographic variables | Effect Size | Standard Error | *P*-value |
| --- | --- | --- | --- |
| Age | -0.00040 | 0.00040 | 0.32 |
| Sex | -0.02093 | 0.00630 | 9.05 × 10^-4^ |
| Ethnicity - Asian | 0.03728 | 0.02628 | 0.16 |
| Ethnicity - Black | 0.03971 | 0.03202 | 0.22 |
| Ethnicity - Chinese | -0.02857 | 0.04911 | 0.56 |
| Ethnicity - Mixed | -0.00548 | 0.03186 | 0.86 |
| Ethnicity - Other | -0.00718 | 0.03311 | 0.83 |
| Place of Birth | -0.00949 | 0.01296 | 0.46 |
| Townsend Deprivation Index | -0.00780 | 0.00307 | 0.011 |
| Smoking - Former | -0.01208 | 0.00967 | 0.21 |
| Smoking - Never | -0.00510 | 0.00924 | 0.58 |
| Body Mass Index | 0.00759 | 0.00284 | 0.0076 |

Probability scores were calculated using Bernoulli-mixtures applied to currently depressed individuals at Q1. All variables were fitted simultaneously. Sex, ethnicity (European ethnicity as the reference), place of birth, and smoking (current smoking as the reference) were fitted as factors. Bold values indicate P-values that were significant after Bonferroni correction.

Supplementary Table 112. Multivariable linear regression of individuals’ probability scores for Attentive and Appetite Disruption on health variables.

| Health variables | Effect Size | Standard Error | *P*-value |
| --- | --- | --- | --- |
| Myocardial infarction | -0.01802 | 0.00597 | 0.0026 |
| Stroke | -0.01534 | 0.01272 | 0.23 |
| Asthma | -0.00532 | 0.01584 | 0.74 |
| COPD | 0.00004 | 0.00734 | 1.00 |
| Dementia | 0.00590 | 0.01130 | 0.60 |
| End stage renal disease | -0.03616 | 0.02415 | 0.13 |
| Motor neurone disease | -0.01526 | 0.05011 | 0.76 |
| Parkinson’s disease | 0.05187 | 0.05593 | 0.35 |

Probability scores were calculated using Bernoulli-mixtures applied to currently depressed individuals at Q1. All variables were fitted simultaneously with age at questionnaire and sex fitted as covariates. Bold values indicate P-values that were significant after Bonferroni correction. COPD = chronic obstructive pulmonary disease.

Supplementary Table 113. Linear regressions of individuals’ probability scores for Attentive and Appetite Disruption on depression recurrence and on treatment resistant depression.

| Depression | Effect Size | Standard Error | *P*-value |
| --- | --- | --- | --- |
| Recurrent | -0.02907 | 0.00839 | 5.41 × 10^-4^ |
| Treatment resistance | 0.01719 | 0.01562 | 0.27 |

Recurrence and treatment resistance were both fitted as factors and were examined separately with age at questionnaire and sex fitted as covariates. Bold values indicate P-values that were significant after Bonferroni correction.

Supplementary Table 114. Linear regressions of individuals’ probability scores for Attentive and Appetite Disruption on polygenic scores for bipolar disorder, schizophrenia, and Attention-Deficit/Hyperactivity Disorder (ADHD).

| Mental health polygenic scores | Effect Size | Standard Error | *P*-value |
| --- | --- | --- | --- |
| Bipolar disorder | 0.00186 | 0.00292 | 0.52 |
| Schizophrenia | 0.00391 | 0.00293 | 0.18 |
| ADHD | -0.00121 | 0.00296 | 0.68 |

The polygenic scores for each trait were examined separately with age, sex, ancestry, and the first 10 genetic principal components fitted as covariates. Bold values indicate P-values that were significant after Bonferroni correction.

#### Preserved Self-Worth and Attentive Disruption

Supplementary Table 115. Multivariable linear regression of individuals’ probability scores for Preserved Self-Worth and Attentive Disruption on demographic variables.

| Demographic variables | Effect Size | Standard Error | *P*-value |
| --- | --- | --- | --- |
| Age | 0.00013 | 0.00039 | 0.79 |
| Sex | 0.01389 | 0.00615 | 0.73 |
| Ethnicity - Asian | -0.04379 | 0.02563 | 0.024 |
| Ethnicity - Black | -0.04232 | 0.03124 | 0.09 |
| Ethnicity - Chinese | 0.03371 | 0.04790 | 0.18 |
| Ethnicity - Mixed | -0.02222 | 0.03108 | 0.48 |
| Ethnicity - Other | 0.04585 | 0.03230 | 0.47 |
| Place of Birth | 0.02274 | 0.01264 | 0.16 |
| Townsend Deprivation Index | 0.00049 | 0.00300 | 0.07 |
| Smoking - Former | 0.00274 | 0.00944 | 0.87 |
| Smoking - Never | 0.01543 | 0.00901 | 0.77 |
| Body Mass Index | -0.00336 | 0.00277 | 0.09 |

Probability scores were calculated using Bernoulli-mixtures applied to currently depressed individuals at Q1. All variables were fitted simultaneously. Sex, ethnicity (European ethnicity as the reference), place of birth, and smoking (current smoking as the reference) were fitted as factors. Bold values indicate P-values that were significant after Bonferroni correction.

Supplementary Table 116. Multivariable linear regression of individuals’ probability scores for Preserved Self-Worth and Attentive Disruption on health variables.

| Health variables | Effect Size | Standard Error | *P*-value |
| --- | --- | --- | --- |
| Myocardial infarction | 0.01064 | 0.00564 | 0.06 |
| Stroke | -0.00351 | 0.01202 | 0.77 |
| Asthma | -0.01358 | 0.01497 | 0.36 |
| COPD | -0.00722 | 0.00694 | 0.30 |
| Dementia | 0.00445 | 0.01068 | 0.68 |
| End stage renal disease | -0.02794 | 0.02282 | 0.22 |
| Motor neurone disease | -0.03058 | 0.04736 | 0.52 |
| Parkinson’s disease | 0.00014 | 0.05286 | 1.00 |

Probability scores were calculated using Bernoulli-mixtures applied to currently depressed individuals at Q1. All variables were fitted simultaneously with age at questionnaire and sex fitted as covariates. Bold values indicate P-values that were significant after Bonferroni correction. COPD = chronic obstructive pulmonary disease.

Supplementary Table 117. Linear regressions of individuals’ probability scores for Preserved Self-Worth and Attentive Disruption on depression recurrence and on treatment resistant depression.

| Depression | Effect Size | Standard Error | *P*-value |
| --- | --- | --- | --- |
| Recurrent | -0.02154 | 0.00798 | 0.0070 |
| Treatment resistance | -0.01380 | 0.01502 | 0.36 |

Recurrence and treatment resistance were both fitted as factors and were examined separately with age at questionnaire and sex fitted as covariates. Bold values indicate P-values that were significant after Bonferroni correction.

Supplementary Table 118. Linear regressions of individuals’ probability scores for Preserved Self-Worth and Attentive Disruption on polygenic scores for bipolar disorder, schizophrenia, and Attention-Deficit/Hyperactivity Disorder (ADHD).

| Mental health polygenic scores | Effect Size | Standard Error | *P*-value |
| --- | --- | --- | --- |
| Bipolar disorder | 0.00116 | 0.00278 | 0.68 |
| Schizophrenia | 0.00214 | 0.00280 | 0.44 |
| ADHD | 0.00127 | 0.00282 | 0.65 |

The polygenic scores for each trait were examined separately with age, sex, ancestry, and the first 10 genetic principal components fitted as covariates. Bold values indicate P-values that were significant after Bonferroni correction.

#### Preserved Self-Worth and Appetite Disruption

Supplementary Table 119. Multivariable linear regression of individuals’ probability scores for Preserved Self-Worth and Appetite Disruption on demographic variables.

| Demographic variables | Effect Size | Standard Error | *P*-value |
| --- | --- | --- | --- |
| Age | 0.00052 | 0.00051 | 0.31 |
| Sex | -0.02019 | 0.00801 | 0.012 |
| Ethnicity - Asian | -0.00210 | 0.03341 | 0.95 |
| Ethnicity - Black | -0.01712 | 0.04072 | 0.67 |
| Ethnicity - Chinese | -0.02172 | 0.06244 | 0.73 |
| Ethnicity - Mixed | -0.00181 | 0.04051 | 0.96 |
| Ethnicity - Other | 0.00570 | 0.04210 | 0.89 |
| Place of Birth | -0.01910 | 0.01648 | 0.25 |
| Townsend Deprivation Index | 0.00229 | 0.00391 | 0.56 |
| Smoking - Former | 0.00105 | 0.01230 | 0.93 |
| Smoking - Never | -0.00668 | 0.01175 | 0.57 |
| Body Mass Index | 0.00201 | 0.00361 | 0.58 |

Probability scores were calculated using Bernoulli-mixtures applied to currently depressed individuals at Q1. All variables were fitted simultaneously. Sex, ethnicity (European ethnicity as the reference), place of birth, and smoking (current smoking as the reference) were fitted as factors. Bold values indicate P-values that were significant after Bonferroni correction.

Supplementary Table 120. Multivariable linear regression of individuals’ probability scores for Preserved Self-Worth and Appetite Disruption on health variables.

| Health variables | Effect Size | Standard Error | *P*-value |
| --- | --- | --- | --- |
| Myocardial infarction | -0.00389 | 0.01624 | 0.81 |
| Stroke | 0.01608 | 0.02023 | 0.43 |
| Asthma | -0.00510 | 0.00938 | 0.59 |
| COPD | -0.00102 | 0.01444 | 0.94 |
| Dementia | 0.00481 | 0.03084 | 0.88 |
| End stage renal disease | 0.06312 | 0.06400 | 0.32 |
| Motor neurone disease | -0.03703 | 0.07144 | 0.60 |
| Parkinson’s disease | -0.02374 | 0.03178 | 0.46 |

Probability scores were calculated using Bernoulli-mixtures applied to currently depressed individuals at Q1. All variables were fitted simultaneously with age at questionnaire and sex fitted as covariates. Bold values indicate P-values that were significant after Bonferroni correction. COPD = chronic obstructive pulmonary disease.

Supplementary Table 121. Linear regressions of individuals’ probability scores for Preserved Self-Worth and Appetite Disruption on depression recurrence and on treatment resistant depression.

| Depression | Effect Size | Standard Error | *P*-value |
| --- | --- | --- | --- |
| Recurrent | -0.04231 | 0.01083 | **9.64 × 10^-5^** |
| Treatment resistance | -0.01106 | 0.02199 | 0.62 |

Recurrence and treatment resistance were both fitted as factors and were examined separately with age at questionnaire and sex fitted as covariates. Bold values indicate P-values that were significant after Bonferroni correction.

Supplementary Table 122. Linear regressions of individuals’ probability scores for Preserved Self-Worth and Appetite Disruption on polygenic scores for bipolar disorder, schizophrenia, and Attention-Deficit/Hyperactivity Disorder (ADHD).

| Mental health polygenic scores | Effect Size | Standard Error | *P*-value |
| --- | --- | --- | --- |
| Bipolar disorder | -0.00663 | 0.00378 | 0.080 |
| Schizophrenia | -0.00496 | 0.00381 | 0.19 |
| ADHD | 0.00565 | 0.00384 | 0.14 |

The polygenic scores for each trait were examined separately with age, sex, ancestry, and the first 10 genetic principal components fitted as covariates. Bold values indicate P-values that were significant after Bonferroni correction.

#### Preserved Self-Worth with SITB

Supplementary Table 123. Multivariable linear regression of individuals’ probability scores for Preserved Self-Worth with SITB on demographic variables.

| Demographic variables | Effect Size | Standard Error | *P*-value |
| --- | --- | --- | --- |
| Age | 0.00272 | 0.00061 | **8.08 × 10^-6^** |
| Sex | 0.01541 | 0.00950 | 0.11 |
| Ethnicity - Asian | -0.00997 | 0.03963 | 0.80 |
| Ethnicity - Black | -0.01671 | 0.04830 | 0.73 |
| Ethnicity - Chinese | 0.01248 | 0.07406 | 0.87 |
| Ethnicity - Mixed | -0.01535 | 0.04805 | 0.75 |
| Ethnicity - Other | 0.01468 | 0.04993 | 0.77 |
| Place of Birth | -0.01015 | 0.01955 | 0.60 |
| Townsend Deprivation Index | -0.00272 | 0.00463 | 0.56 |
| Smoking - Former | -0.00162 | 0.01459 | 0.91 |
| Smoking - Never | 0.00622 | 0.01393 | 0.66 |
| Body Mass Index | 0.00005 | 0.00429 | 0.99 |

Probability scores were calculated using Bernoulli-mixtures applied to currently depressed individuals at Q1. All variables were fitted simultaneously. Sex, ethnicity (European ethnicity as the reference), place of birth, and smoking (current smoking as the reference) were fitted as factors. Bold values indicate P-values that were significant after Bonferroni correction.

Supplementary Table 124. Multivariable linear regression of individuals’ probability scores for Preserved Self-Worth with SITB on health variables.

| Health variables | Effect Size | Standard Error | *P*-value |
| --- | --- | --- | --- |
| Myocardial infarction | -0.00340 | 0.01898 | 0.86 |
| Stroke | -0.00492 | 0.02365 | 0.84 |
| Asthma | 0.00784 | 0.01096 | 0.47 |
| COPD | 0.01157 | 0.01687 | 0.49 |
| Dementia | -0.02163 | 0.03605 | 0.55 |
| End stage renal disease | -0.06411 | 0.07481 | 0.39 |
| Motor neurone disease | -0.06380 | 0.08350 | 0.44 |
| Parkinson’s disease | -0.03835 | 0.03715 | 0.30 |

Probability scores were calculated using Bernoulli-mixtures applied to currently depressed individuals at Q1. All variables were fitted simultaneously with age at questionnaire and sex fitted as covariates. Bold values indicate P-values that were significant after Bonferroni correction. COPD = chronic obstructive pulmonary disease.

Supplementary Table 125. Linear regressions of individuals’ probability scores for Preserved Self-Worth with SITB on depression recurrence and on treatment resistant depression.

| Depression | Effect Size | Standard Error | *P*-value |
| --- | --- | --- | --- |
| Recurrent | 0.01005 | 0.01297 | 0.44 |
| Treatment resistance | 0.02192 | 0.02535 | 0.39 |

Recurrence and treatment resistance were both fitted as factors and were examined separately with age at questionnaire and sex fitted as covariates. Bold values indicate P-values that were significant after Bonferroni correction.

Supplementary Table 126. Linear regressions of individuals’ probability scores for Preserved Self-Worth with SITB on polygenic scores for bipolar disorder, schizophrenia, and Attention-Deficit/Hyperactivity Disorder (ADHD).

| Mental health polygenic scores | Effect Size | Standard Error | *P*-value |
| --- | --- | --- | --- |
| Bipolar disorder | -0.00733 | 0.00438 | 0.09 |
| Schizophrenia | 0.00809 | 0.00440 | 0.07 |
| ADHD | -0.00203 | 0.00445 | 0.65 |

The polygenic scores for each trait were examined separately with age, sex, ancestry, and the first 10 genetic principal components fitted as covariates. Bold values indicate P-values that were significant after Bonferroni correction.

#### Low Self-Worth with SITB

Supplementary Table 127. Multivariable linear regression of individuals’ probability scores for Low Self-Worth with SITB on demographic variables.

| Demographic variables | Effect Size | Standard Error | *P*-value |
| --- | --- | --- | --- |
| Age | -0.00025 | 0.00062 | 0.68 |
| Sex | 0.00898 | 0.00962 | 0.35 |
| Ethnicity - Asian | -0.03724 | 0.04013 | 0.35 |
| Ethnicity - Black | -0.06247 | 0.04891 | 0.20 |
| Ethnicity - Chinese | 0.15859 | 0.07500 | 0.035 |
| Ethnicity - Mixed | 0.05119 | 0.04866 | 0.29 |
| Ethnicity - Other | 0.08279 | 0.05057 | 0.10 |
| Place of Birth | 0.01179 | 0.01980 | 0.55 |
| Townsend Deprivation Index | 0.00377 | 0.00469 | 0.42 |
| Smoking - Former | 0.04325 | 0.01478 | 0.00345 |
| Smoking - Never | 0.04282 | 0.01411 | 0.00243 |
| Body Mass Index | -0.02394 | 0.00434 | **3.85 × 10^-8^** |

Probability scores were calculated using Bernoulli-mixtures applied to currently depressed individuals at Q1. All variables were fitted simultaneously. Sex, ethnicity (European ethnicity as the reference), place of birth, and smoking (current smoking as the reference) were fitted as factors. Bold values indicate P-values that were significant after Bonferroni correction.

Supplementary Table 128. Multivariable linear regression of individuals’ probability scores for Low Self-Worth with SITB on health variables.

| Health variables | Effect Size | Standard Error | *P*-value |
| --- | --- | --- | --- |
| Myocardial infarction | -0.01382 | 0.01934 | 0.48 |
| Stroke | -0.00457 | 0.02410 | 0.85 |
| Asthma | -0.00743 | 0.01117 | 0.51 |
| COPD | -0.03289 | 0.01719 | 0.06 |
| Dementia | 0.06777 | 0.03674 | 0.07 |
| End stage renal disease | 0.09986 | 0.07624 | 0.19 |
| Motor neurone disease | 0.10570 | 0.08510 | 0.21 |
| Parkinson’s disease | -0.08140 | 0.03786 | 0.032 |

Probability scores were calculated using Bernoulli-mixtures applied to currently depressed individuals at Q1. All variables were fitted simultaneously with age at questionnaire and sex fitted as covariates. Bold values indicate P-values that were significant after Bonferroni correction. COPD = chronic obstructive pulmonary disease.

Supplementary Table 129. Linear regressions of individuals’ probability scores for Low Self-Worth with SITB on depression recurrence and on treatment resistant depression.

| Depression | Effect Size | Standard Error | *P*-value |
| --- | --- | --- | --- |
| Recurrent | 0.05075 | 0.01315 | **1.16 × 10^-4^** |
| Treatment resistance | 0.00413 | 0.02644 | 0.88 |

Recurrence and treatment resistance were both fitted as factors and were examined separately with age at questionnaire and sex fitted as covariates. Bold values indicate P-values that were significant after Bonferroni correction.

Supplementary Table 130. Linear regressions of individuals’ probability scores for Low Self-Worth with SITB on polygenic scores for bipolar disorder, schizophrenia, and Attention-Deficit/Hyperactivity Disorder (ADHD).

| Mental health polygenic scores | Effect Size | Standard Error | *P*-value |
| --- | --- | --- | --- |
| Bipolar disorder | 0.00129 | 0.00446 | 0.77 |
| Schizophrenia | -0.00317 | 0.00449 | 0.48 |
| ADHD | -0.01062 | 0.00453 | 0.019 |

The polygenic scores for each trait were examined separately with age, sex, ancestry, and the first 10 genetic principal components fitted as covariates. Bold values indicate P-values that were significant after Bonferroni correction.

#### Low Self-Worth

Supplementary Table 131. Multivariable linear regression of individuals’ probability scores for Low Self-Worth on demographic variables.

| Demographic variables | Effect Size | Standard Error | *P*-value |
| --- | --- | --- | --- |
| Age | -0.00097 | 0.00080 | 0.23 |
| Sex | -0.01084 | 0.01251 | 0.39 |
| Ethnicity - Asian | -0.02548 | 0.05216 | 0.63 |
| Ethnicity - Black | -0.01604 | 0.06356 | 0.80 |
| Ethnicity - Chinese | 0.16210 | 0.09747 | 0.10 |
| Ethnicity - Mixed | -0.11070 | 0.06324 | 0.08 |
| Ethnicity - Other | -0.08673 | 0.06572 | 0.19 |
| Place of Birth | -0.00250 | 0.02573 | 0.92 |
| Townsend Deprivation Index | -0.00002 | 0.00610 | 1.00 |
| Smoking - Former | -0.00758 | 0.01920 | 0.69 |
| Smoking - Never | 0.00037 | 0.01834 | 0.98 |
| Body Mass Index | -0.01188 | 0.00564 | 0.035 |

Probability scores were calculated using Bernoulli-mixtures applied to currently depressed individuals at Q1. All variables were fitted simultaneously. Sex, ethnicity (European ethnicity as the reference), place of birth, and smoking (current smoking as the reference) were fitted as factors. Bold values indicate P-values that were significant after Bonferroni correction.

Supplementary Table 132. Multivariable linear regression of individuals’ probability scores for Low Self-Worth on health variables.

| Health variables | Effect Size | Standard Error | *P*-value |
| --- | --- | --- | --- |
| Myocardial infarction | -0.00098 | 0.02490 | 0.97 |
| Stroke | -0.03005 | 0.03101 | 0.33 |
| Asthma | -0.01621 | 0.01437 | 0.26 |
| COPD | -0.03264 | 0.02213 | 0.14 |
| Dementia | -0.03525 | 0.04728 | 0.46 |
| End stage renal disease | -0.06289 | 0.09811 | 0.52 |
| Motor neurone disease | -0.11885 | 0.10951 | 0.28 |
| Parkinson’s disease | -0.15257 | 0.04872 | 0.0018 |

Probability scores were calculated using Bernoulli-mixtures applied to currently depressed individuals at Q1. All variables were fitted simultaneously with age at questionnaire and sex fitted as covariates. Bold values indicate P-values that were significant after Bonferroni correction. COPD = chronic obstructive pulmonary disease.

Supplementary Table 133. Linear regressions of individuals’ probability scores for Low Self-Worth on depression recurrence and on treatment resistant depression.

| Depression | Effect Size | Standard Error | *P*-value |
| --- | --- | --- | --- |
| Recurrent | 0.01146 | 0.01681 | 0.50 |
| Treatment resistance | -0.05217 | 0.03270 | 0.11 |

Recurrence and treatment resistance were both fitted as factors and were examined separately with age at questionnaire and sex fitted as covariates. Bold values indicate P-values that were significant after Bonferroni correction.

Supplementary Table 134. Linear regressions of individuals’ probability scores for Low Self-Worth on polygenic scores for bipolar disorder, schizophrenia, and Attention-Deficit/Hyperactivity Disorder (ADHD).

| Mental health polygenic scores | Effect Size | Standard Error | *P*-value |
| --- | --- | --- | --- |
| Bipolar disorder | -0.00474 | 0.00577 | 0.41 |
| Schizophrenia | -0.00093 | 0.00581 | 0.87 |
| ADHD | -0.02135 | 0.00585 | 2.67 × 10^-4^ |

The polygenic scores for each trait were examined separately with age, sex, ancestry, and the first 10 genetic principal components fitted as covariates. Bold values indicate P-values that were significant after Bonferroni correction.

#### Low Self-Worth and Attentive Disruption

Supplementary Table 135. Multivariable linear regression of individuals’ probability scores for Low Self-Worth and Attentive Disruption on demographic variables.

| Demographic variables | Effect Size | Standard Error | *P*-value |
| --- | --- | --- | --- |
| Age | 0.00077 | 0.00036 | 0.031 |
| Sex | 0.00805 | 0.00558 | 0.15 |
| Ethnicity - Asian | -0.00855 | 0.02326 | 0.71 |
| Ethnicity - Black | -0.02030 | 0.02835 | 0.47 |
| Ethnicity - Chinese | 0.00799 | 0.04348 | 0.85 |
| Ethnicity - Mixed | -0.00894 | 0.02821 | 0.75 |
| Ethnicity - Other | 0.01219 | 0.02931 | 0.68 |
| Place of Birth | 0.01407 | 0.01148 | 0.22 |
| Townsend Deprivation Index | 0.00163 | 0.00272 | 0.55 |
| Smoking - Former | -0.00382 | 0.00857 | 0.66 |
| Smoking - Never | -0.00402 | 0.00818 | 0.62 |
| Body Mass Index | -0.00993 | 0.00252 | **8.11 × 10^-5^** |

Probability scores were calculated using Bernoulli-mixtures applied to currently depressed individuals at Q1. All variables were fitted simultaneously. Sex, ethnicity (European ethnicity as the reference), place of birth, and smoking (current smoking as the reference) were fitted as factors. Bold values indicate P-values that were significant after Bonferroni correction.

Supplementary Table 136. Multivariable linear regression of individuals’ probability scores for Low Self-Worth and Attentive Disruption on health variables.

| Health variables | Effect Size | Standard Error | *P*-value |
| --- | --- | --- | --- |
| Myocardial infarction | -0.00454 | 0.01124 | 0.69 |
| Stroke | -0.00698 | 0.01400 | 0.62 |
| Asthma | -0.00546 | 0.00649 | 0.40 |
| COPD | -0.01135 | 0.00999 | 0.26 |
| Dementia | 0.00690 | 0.02135 | 0.75 |
| End stage renal disease | -0.03689 | 0.04430 | 0.41 |
| Motor neurone disease | -0.02860 | 0.04945 | 0.56 |
| Parkinson’s disease | 0.01604 | 0.02200 | 0.47 |

Probability scores were calculated using Bernoulli-mixtures applied to currently depressed individuals at Q1. All variables were fitted simultaneously with age at questionnaire and sex fitted as covariates. Bold values indicate P-values that were significant after Bonferroni correction. COPD = chronic obstructive pulmonary disease.

Supplementary Table 137. Linear regressions of individuals’ probability scores for Low Self-Worth and Attentive Disruption on depression recurrence and on treatment resistant depression.

| Depression | Effect Size | Standard Error | *P*-value |
| --- | --- | --- | --- |
| Recurrent | 0.00199 | 0.00761 | 0.79 |
| Treatment resistance | 0.02459 | 0.01499 | 0.10 |

Recurrence and treatment resistance were both fitted as factors and were examined separately with age at questionnaire and sex fitted as covariates. Bold values indicate P-values that were significant after Bonferroni correction.

Supplementary Table 138. Linear regressions of individuals’ probability scores for Low Self-Worth and Attentive Disruption on polygenic scores for bipolar disorder, schizophrenia, and Attention-Deficit/Hyperactivity Disorder (ADHD).

| Mental health polygenic scores | Effect Size | Standard Error | *P*-value |
| --- | --- | --- | --- |
| Bipolar disorder | 0.00186 | 0.00260 | 0.47 |
| Schizophrenia | -0.00228 | 0.00262 | 0.38 |
| ADHD | -0.00380 | 0.00264 | 0.15 |

The polygenic scores for each trait were examined separately with age, sex, ancestry, and the first 10 genetic principal components fitted as covariates. Bold values indicate P-values that were significant after Bonferroni correction.

#### Psychomotor Changes

Supplementary Table 139. Multivariable linear regression of individuals’ probability scores for Psychomotor Changes on demographic variables.

| Demographic variables | Effect Size | Standard Error | *P*-value |
| --- | --- | --- | --- |
| Age | 0.00219 | 0.00062 | 4.01 × 10^-4^ |
| Sex | 0.00365 | 0.00964 | 0.71 |
| Ethnicity - Asian | 0.08042 | 0.04021 | 0.046 |
| Ethnicity - Black | 0.07490 | 0.04900 | 0.13 |
| Ethnicity - Chinese | -0.12811 | 0.07514 | 0.09 |
| Ethnicity - Mixed | 0.05071 | 0.04875 | 0.30 |
| Ethnicity - Other | -0.07065 | 0.05067 | 0.16 |
| Place of Birth | 0.03250 | 0.01983 | 0.10 |
| Townsend Deprivation Index | -0.01047 | 0.00470 | 0.026 |
| Smoking - Former | 0.01294 | 0.01480 | 0.38 |
| Smoking - Never | 0.02259 | 0.01414 | 0.11 |
| Body Mass Index | -0.00024 | 0.00435 | 0.96 |

Probability scores were calculated using Bernoulli-mixtures applied to currently depressed individuals at Q1. All variables were fitted simultaneously. Sex, ethnicity (European ethnicity as the reference), place of birth, and smoking (current smoking as the reference) were fitted as factors. Bold values indicate P-values that were significant after Bonferroni correction.

Supplementary Table 140. Multivariable linear regression of individuals’ probability scores for Psychomotor Changes on health variables.

| Health variables | Effect Size | Standard Error | *P*-value |
| --- | --- | --- | --- |
| Myocardial infarction | -0.00565 | 0.01931 | 0.64 |
| Stroke | 0.02341 | 0.02406 | 0.77 |
| Asthma | 0.00365 | 0.01115 | 0.33 |
| COPD | 0.00446 | 0.01717 | 0.74 |
| Dementia | 0.05701 | 0.03668 | 0.79 |
| End stage renal disease | 0.02776 | 0.07611 | 0.12 |
| Motor neurone disease | 0.13568 | 0.08496 | 0.72 |
| Parkinson’s disease | 0.18523 | 0.03780 | **1.00 × 10^-6^** |

Probability scores were calculated using Bernoulli-mixtures applied to currently depressed individuals at Q1. All variables were fitted simultaneously with age at questionnaire and sex fitted as covariates. Bold values indicate P-values that were significant after Bonferroni correction. COPD = chronic obstructive pulmonary disease.

Supplementary Table 141. Linear regressions of individuals’ probability scores for Psychomotor Changes on depression recurrence and on treatment resistant depression.

| Depression | Effect Size | Standard Error | *P*-value |
| --- | --- | --- | --- |
| Recurrent | -0.02115 | 0.01295 | 0.10 |
| Treatment resistance | -0.03655 | 0.02032 | 0.07 |

Recurrence and treatment resistance were both fitted as factors and were examined separately with age at questionnaire and sex fitted as covariates. Bold values indicate P-values that were significant after Bonferroni correction.

Supplementary Table 142. Linear regressions of individuals’ probability scores for Psychomotor Changes on polygenic scores for bipolar disorder, schizophrenia, and Attention-Deficit/Hyperactivity Disorder (ADHD).

| Mental health polygenic scores | Effect Size | Standard Error | *P*-value |
| --- | --- | --- | --- |
| Bipolar disorder | 0.00608 | 0.00451 | 0.18 |
| Schizophrenia | -0.00161 | 0.00454 | 0.72 |
| ADHD | 0.00570 | 0.00458 | 0.21 |

The polygenic scores for each trait were examined separately with age, sex, ancestry, and the first 10 genetic principal components fitted as covariates. Bold values indicate P-values that were significant after Bonferroni correction.

#### Anhedonia and Low Self-Worth with SITB

Supplementary Table 143. Multivariable linear regression of individuals’ probability scores for Anhedonia and Low Self-Worth with SITB on demographic variables.

| Demographic variables | Effect Size | Standard Error | *P*-value |
| --- | --- | --- | --- |
| Age | 0.00016 | 0.00019 | 0.39 |
| Sex | 0.00295 | 0.00290 | 0.31 |
| Ethnicity - Asian | 0.02222 | 0.01209 | 0.07 |
| Ethnicity - Black | -0.00976 | 0.01473 | 0.51 |
| Ethnicity - Chinese | -0.00914 | 0.02259 | 0.69 |
| Ethnicity - Mixed | 0.02305 | 0.01465 | 0.12 |
| Ethnicity - Other | -0.01087 | 0.01523 | 0.48 |
| Place of Birth | -0.00011 | 0.00596 | 0.99 |
| Townsend Deprivation Index | 0.00091 | 0.00141 | 0.52 |
| Smoking - Former | -0.01077 | 0.00445 | 0.0156 |
| Smoking - Never | -0.01140 | 0.00425 | 0.0074 |
| Body Mass Index | -0.00005 | 0.00131 | 0.97 |

Probability scores were calculated using Bernoulli-mixtures applied to currently depressed individuals at Q1. All variables were fitted simultaneously. Sex, ethnicity (European ethnicity as the reference), place of birth, and smoking (current smoking as the reference) were fitted as factors. Bold values indicate P-values that were significant after Bonferroni correction.

Supplementary Table 144. Multivariable linear regression of individuals’ probability scores for Anhedonia and Low Self-Worth with SITB on health variables.

| Health variables | Effect Size | Standard Error | *P*-value |
| --- | --- | --- | --- |
| Myocardial infarction | -0.00071 | 0.00580 | 0.90 |
| Stroke | -0.01208 | 0.00722 | 0.09 |
| Asthma | -0.00160 | 0.00335 | 0.63 |
| COPD | 0.00334 | 0.00515 | 0.52 |
| Dementia | -0.01144 | 0.01101 | 0.30 |
| End stage renal disease | -0.00093 | 0.02285 | 0.97 |
| Motor neurone disease | 0.01273 | 0.02551 | 0.62 |
| Parkinson’s disease | -0.00273 | 0.01135 | 0.81 |

Probability scores were calculated using Bernoulli-mixtures applied to currently depressed individuals at Q1. All variables were fitted simultaneously with age at questionnaire and sex fitted as covariates. Bold values indicate P-values that were significant after Bonferroni correction. COPD = chronic obstructive pulmonary disease.

Supplementary Table 145. Linear regressions of individuals’ probability scores for Anhedonia and Low Self-Worth with SITB on depression recurrence and on treatment resistant depression.

| Depression | Effect Size | Standard Error | *P*-value |
| --- | --- | --- | --- |
| Recurrent | 0.00215 | 0.00383 | 0.58 |
| Treatment resistance | -0.00920 | 0.00853 | 0.28 |

Recurrence and treatment resistance were both fitted as factors and were examined separately with age at questionnaire and sex fitted as covariates. Bold values indicate P-values that were significant after Bonferroni correction.

Supplementary Table 146. Linear regressions of individuals’ probability scores for Anhedonia and Low Self-Worth with SITB on polygenic scores for bipolar disorder, schizophrenia, and Attention-Deficit/Hyperactivity Disorder (ADHD).

| Mental health polygenic scores | Effect Size | Standard Error | *P*-value |
| --- | --- | --- | --- |
| Bipolar disorder | -0.00071 | 0.00135 | 0.60 |
| Schizophrenia | -0.00165 | 0.00136 | 0.22 |
| ADHD | 0.00105 | 0.00137 | 0.44 |

The polygenic scores for each trait were examined separately with age, sex, ancestry, and the first 10 genetic principal components fitted as covariates. Bold values indicate P-values that were significant after Bonferroni correction.

#### General Anhedonia

Supplementary Table 147. Multivariable linear regression of individuals’ probability scores for General Anhedonia on demographic variables.

| Demographic variables | Effect Size | Standard Error | *P*-value |
| --- | --- | --- | --- |
| Age | -0.00129 | 0.00074 | 0.08 |
| Sex | 0.03221 | 0.01158 | 0.0054 |
| Ethnicity - Asian | -0.00180 | 0.04828 | 0.97 |
| Ethnicity - Black | 0.04098 | 0.05884 | 0.49 |
| Ethnicity - Chinese | -0.09749 | 0.09023 | 0.28 |
| Ethnicity - Mixed | 0.01978 | 0.05854 | 0.74 |
| Ethnicity - Other | 0.05970 | 0.06084 | 0.33 |
| Place of Birth | -0.00088 | 0.02382 | 0.97 |
| Townsend Deprivation Index | 0.00267 | 0.00565 | 0.64 |
| Smoking - Former | -0.01938 | 0.01778 | 0.28 |
| Smoking - Never | -0.02813 | 0.01698 | 0.10 |
| Body Mass Index | 0.00841 | 0.00522 | 0.11 |

Probability scores were calculated using Bernoulli-mixtures applied to currently depressed individuals at Q1. All variables were fitted simultaneously. Sex, ethnicity (European ethnicity as the reference), place of birth, and smoking (current smoking as the reference) were fitted as factors. Bold values indicate P-values that were significant after Bonferroni correction.

Supplementary Table 148. Multivariable linear regression of individuals’ probability scores for General Anhedonia on health variables.

| Health variables | Effect Size | Standard Error | *P*-value |
| --- | --- | --- | --- |
| Myocardial infarction | -0.01761 | 0.02383 | 0.46 |
| Stroke | -0.02157 | 0.02969 | 0.47 |
| Asthma | 0.00855 | 0.01376 | 0.53 |
| COPD | -0.01129 | 0.02118 | 0.59 |
| Dementia | -0.03740 | 0.04526 | 0.41 |
| End stage renal disease | 0.07357 | 0.09393 | 0.43 |
| Motor neurone disease | -0.02880 | 0.10484 | 0.78 |
| Parkinson’s disease | 0.07886 | 0.04664 | 0.09 |

Probability scores were calculated using Bernoulli-mixtures applied to currently depressed individuals at Q1. All variables were fitted simultaneously with age at questionnaire and sex fitted as covariates. Bold values indicate P-values that were significant after Bonferroni correction. COPD = chronic obstructive pulmonary disease.

Supplementary Table 149. Linear regressions of individuals’ probability scores for General Anhedonia on depression recurrence and on treatment resistant depression.

| Depression | Effect Size | Standard Error | *P*-value |
| --- | --- | --- | --- |
| Recurrent | -0.04677 | 0.01595 | 0.0034 |
| Treatment resistance | -0.04212 | 0.02887 | 0.15 |

Recurrence and treatment resistance were both fitted as factors and were examined separately with age at questionnaire and sex fitted as covariates. Bold values indicate P-values that were significant after Bonferroni correction.

Supplementary Table 150. Linear regressions of individuals’ probability scores for General Anhedonia on polygenic scores for bipolar disorder, schizophrenia, and Attention-Deficit/Hyperactivity Disorder (ADHD).

| Mental health polygenic scores | Effect Size | Standard Error | *P*-value |
| --- | --- | --- | --- |
| Bipolar disorder | 0.00722 | 0.00546 | 0.19 |
| Schizophrenia | -0.00240 | 0.00549 | 0.66 |
| ADHD | 0.00063 | 0.00554 | 0.91 |

The polygenic scores for each trait were examined separately with age, sex, ancestry, and the first 10 genetic principal components fitted as covariates. Bold values indicate P-values that were significant after Bonferroni correction.

#### All Symptoms

Supplementary Table 151. Multivariable linear regression of individuals’ probability scores for All Symptoms on demographic variables.

| Demographic variables | Effect Size | Standard Error | *P*-value |
| --- | --- | --- | --- |
| Age | -0.00358 | 0.00074 | **1.40 × 10^-6^** |
| Sex | -0.03317 | 0.01157 | 0.0042 |
| Ethnicity - Asian | -0.01099 | 0.04825 | 0.82 |
| Ethnicity - Black | 0.02912 | 0.05880 | 0.62 |
| Ethnicity - Chinese | -0.08984 | 0.09017 | 0.32 |
| Ethnicity - Mixed | 0.01982 | 0.05850 | 0.73 |
| Ethnicity - Other | -0.04548 | 0.06080 | 0.45 |
| Place of Birth | -0.03888 | 0.02380 | 0.10 |
| Townsend Deprivation Index | 0.00925 | 0.00564 | 0.10 |
| Smoking - Former | -0.00473 | 0.01776 | 0.79 |
| Smoking - Never | -0.03211 | 0.01697 | 0.059 |
| Body Mass Index | 0.03134 | 0.00522 | **2.17 × 10^-9^** |

Probability scores were calculated using Bernoulli-mixtures applied to currently depressed individuals at Q1. All variables were fitted simultaneously. Sex, ethnicity (European ethnicity as the reference), place of birth, and smoking (current smoking as the reference) were fitted as factors. Bold values indicate P-values that were significant after Bonferroni correction.

Supplementary Table 152. Multivariable linear regression of individuals’ probability scores for All Symptoms on health variables.

| Health variables | Effect Size | Standard Error | *P*-value |
| --- | --- | --- | --- |
| Myocardial infarction | 0.06945 | 0.02386 | 0.0036 |
| Stroke | 0.05958 | 0.02973 | 0.045 |
| Asthma | 0.02296 | 0.01378 | 0.10 |
| COPD | 0.05947 | 0.02121 | 0.005 |
| Dementia | 0.03334 | 0.04532 | 0.46 |
| End stage renal disease | -0.05365 | 0.09405 | 0.57 |
| Motor neurone disease | -0.02904 | 0.10498 | 0.78 |
| Parkinson’s disease | 0.03009 | 0.04670 | 0.52 |

Probability scores were calculated using Bernoulli-mixtures applied to currently depressed individuals at Q1. All variables were fitted simultaneously with age at questionnaire and sex fitted as covariates. Bold values indicate P-values that were significant after Bonferroni correction. COPD = chronic obstructive pulmonary disease.

Supplementary Table 153. Linear regressions of individuals’ probability scores for All Symptoms on depression recurrence and on treatment resistant depression.

| Depression | Effect Size | Standard Error | *P*-value |
| --- | --- | --- | --- |
| Recurrent | 0.08442 | 0.01644 | **2.99 × 10^-7^** |
| Treatment resistance | 0.05486 | 0.03622 | 0.13 |

Recurrence and treatment resistance were both fitted as factors and were examined separately with age at questionnaire and sex fitted as covariates. Bold values indicate P-values that were significant after Bonferroni correction.

Supplementary Table 154. Linear regressions of individuals’ probability scores for All Symptoms on polygenic scores for bipolar disorder, schizophrenia, and Attention-Deficit/Hyperactivity Disorder (ADHD).

| Mental health polygenic scores | Effect Size | Standard Error | *P*-value |
| --- | --- | --- | --- |
| Bipolar disorder | -0.00006 | 0.00555 | 0.99 |
| Schizophrenia | 0.00286 | 0.00559 | 0.61 |
| ADHD | 0.02471 | 0.00562 | **1.14 × 10^-5^** |

The polygenic scores for each trait were examined separately with age, sex, ancestry, and the first 10 genetic principal components fitted as covariates. Bold values indicate P-values that were significant after Bonferroni correction.

### Q2 (Mental Well-being Questionnaire) - *Currently depressed*

Number of individuals analysed: 3,240

Number of clusters identified: 11

#### Attentive and Appetite Disruption

Supplementary Table 155. Multivariable linear regression of individuals’ probability scores for Attentive and Appetite Disruption on demographic variables.

| Demographic variables | Effect Size | Standard Error | *P*-value |
| --- | --- | --- | --- |
| Age | -0.00040 | 0.00054 | 0.46 |
| Sex | -0.01569 | 0.00850 | 0.06 |
| Ethnicity - Asian | -0.04065 | 0.03858 | 0.29 |
| Ethnicity - Black | 0.02790 | 0.04523 | 0.54 |
| Ethnicity - Chinese | 0.00046 | 0.09960 | 1.00 |
| Ethnicity - Mixed | -0.06386 | 0.04353 | 0.14 |
| Ethnicity - Other | -0.03578 | 0.05503 | 0.52 |
| Place of Birth | -0.00341 | 0.01757 | 0.85 |
| Townsend Deprivation Index | 0.00334 | 0.00415 | 0.42 |
| Smoking - Former | 0.01411 | 0.01312 | 0.28 |
| Smoking - Never | 0.00733 | 0.01256 | 0.56 |
| Body Mass Index | 0.00475 | 0.00386 | 0.22 |

Probability scores were calculated using Bernoulli-mixtures applied to currently depressed individuals at Q2. All variables were fitted simultaneously. Sex, ethnicity (European ethnicity as the reference), place of birth, and smoking (current smoking as the reference) were fitted as factors. Bold values indicate P-values that were significant after Bonferroni correction.

Supplementary Table 156. Multivariable linear regression of individuals’ probability scores for Attentive and Appetite Disruption on health variables.

| Health variables | Effect Size | Standard Error | *P*-value |
| --- | --- | --- | --- |
| Myocardial infarction | -0.00945 | 0.02012 | 0.64 |
| Stroke | -0.04692 | 0.02235 | 0.036 |
| Asthma | -0.01000 | 0.00967 | 0.30 |
| COPD | -0.00325 | 0.01685 | 0.85 |
| Dementia | -0.04456 | 0.06366 | 0.48 |
| End stage renal disease | 0.03044 | 0.08363 | 0.72 |
| Parkinson’s disease | -0.05827 | 0.05281 | 0.27 |

Probability scores were calculated using Bernoulli-mixtures applied to currently depressed individuals at Q2. All variables were fitted simultaneously with age at questionnaire and sex fitted as covariates. Bold values indicate P-values that were significant after Bonferroni correction. COPD = chronic obstructive pulmonary disease.

Supplementary Table 157. Linear regressions of individuals’ probability scores for Attentive and Appetite Disruption on depression recurrence and on treatment resistant depression.

| Depression | Effect Size | Standard Error | *P*-value |
| --- | --- | --- | --- |
| Recurrent | -0.04812 | 0.01459 | 0.0010 |
| Treatment resistance | -0.00622 | 0.02426 | 0.80 |

Recurrence and treatment resistance were both fitted as factors and were examined separately with age at questionnaire and sex fitted as covariates. Bold values indicate P-values that were significant after Bonferroni correction.

Supplementary Table 158. Linear regressions of individuals’ probability scores for Attentive and Appetite Disruption on polygenic scores for bipolar disorder, schizophrenia, and Attention-Deficit/Hyperactivity Disorder (ADHD).

| Mental health polygenic scores | Effect Size | Standard Error | *P*-value |
| --- | --- | --- | --- |
| Bipolar disorder | -0.00471 | 0.00422 | 0.26 |
| Schizophrenia | -0.00699 | 0.00409 | 0.09 |
| ADHD | 0.00144 | 0.00412 | 0.73 |

The polygenic scores for each trait were examined separately with age, sex, ancestry, and the first 10 genetic principal components fitted as covariates. Bold values indicate P-values that were significant after Bonferroni correction.

#### Preserved Self-Worth and Attentive Disruption

Supplementary Table 159. Multivariable linear regression of individuals’ probability scores for Preserved Self-Worth and Attentive Disruption on demographic variables.

| Demographic variables | Effect Size | Standard Error | *P*-value |
| --- | --- | --- | --- |
| Age | 0.00064 | 0.00043 | 0.13 |
| Sex | 0.01654 | 0.00676 | 0.014 |
| Ethnicity - Asian | -0.01293 | 0.03068 | 0.67 |
| Ethnicity - Black | 0.00672 | 0.03597 | 0.85 |
| Ethnicity - Chinese | -0.05043 | 0.07920 | 0.52 |
| Ethnicity - Mixed | 0.01028 | 0.03461 | 0.77 |
| Ethnicity - Other | -0.04455 | 0.04377 | 0.31 |
| Place of Birth | 0.02484 | 0.01398 | 0.08 |
| Townsend Deprivation Index | -0.00236 | 0.00330 | 0.47 |
| Smoking - Former | 0.02297 | 0.01044 | 0.028 |
| Smoking - Never | 0.01654 | 0.00999 | 0.10 |
| Body Mass Index | -0.00287 | 0.00307 | 0.35 |

Probability scores were calculated using Bernoulli-mixtures applied to currently depressed individuals at Q2. All variables were fitted simultaneously. Sex, ethnicity (European ethnicity as the reference), place of birth, and smoking (current smoking as the reference) were fitted as factors. Bold values indicate P-values that were significant after Bonferroni correction.

Supplementary Table 160. Multivariable linear regression of individuals’ probability scores for Preserved Self-Worth and Attentive Disruption on health variables.

| Health variables | Effect Size | Standard Error | *P*-value |
| --- | --- | --- | --- |
| Myocardial infarction | -0.01754 | 0.01751 | 0.32 |
| Stroke | 0.01366 | 0.00757 | 0.07 |
| Asthma | -0.00780 | 0.01321 | 0.56 |
| COPD | -0.02431 | 0.04988 | 0.63 |
| Dementia | -0.03558 | 0.06553 | 0.59 |
| End stage renal disease | -0.02219 | 0.04138 | 0.59 |
| Motor neurone disease | -0.01754 | 0.01751 | 0.32 |
| Parkinson’s disease | 0.01366 | 0.00757 | 0.07 |

Probability scores were calculated using Bernoulli-mixtures applied to currently depressed individuals at Q2. All variables were fitted simultaneously with age at questionnaire and sex fitted as covariates. Bold values indicate P-values that were significant after Bonferroni correction. COPD = chronic obstructive pulmonary disease.

Supplementary Table 161. Linear regressions of individuals’ probability scores for Preserved Self-Worth and Attentive Disruption on depression recurrence and on treatment resistant depression.

| Depression | Effect Size | Standard Error | *P*-value |
| --- | --- | --- | --- |
| Recurrent | -0.01129 | 0.01130 | 0.32 |
| Treatment resistance | 0.00130 | 0.01859 | 0.94 |

Recurrence and treatment resistance were both fitted as factors and were examined separately with age at questionnaire and sex fitted as covariates. Bold values indicate P-values that were significant after Bonferroni correction.

Supplementary Table 162. Linear regressions of individuals’ probability scores for Preserved Self-Worth and Attentive Disruption on polygenic scores for bipolar disorder, schizophrenia, and Attention-Deficit/Hyperactivity Disorder (ADHD).

| Mental health polygenic scores | Effect Size | Standard Error | *P*-value |
| --- | --- | --- | --- |
| Bipolar disorder | 0.00344 | 0.00329 | 0.30 |
| Schizophrenia | -0.00156 | 0.00319 | 0.63 |
| ADHD | 0.00308 | 0.00321 | 0.34 |

The polygenic scores for each trait were examined separately with age, sex, ancestry, and the first 10 genetic principal components fitted as covariates. Bold values indicate P-values that were significant after Bonferroni correction.

#### Preserved Self-Worth and Appetite Disruption

Supplementary Table 163. Multivariable linear regression of individuals’ probability scores for Preserved Self-Worth and Appetite Disruption on demographic variables.

| Demographic variables | Effect Size | Standard Error | *P*-value |
| --- | --- | --- | --- |
| Age | 0.00043 | 0.00053 | 0.42 |
| Sex | -0.02728 | 0.00834 | 0.0011 |
| Ethnicity - Asian | -0.01559 | 0.03787 | 0.68 |
| Ethnicity - Black | -0.01236 | 0.04439 | 0.78 |
| Ethnicity - Chinese | -0.03633 | 0.09775 | 0.71 |
| Ethnicity - Mixed | -0.00966 | 0.04272 | 0.82 |
| Ethnicity - Other | 0.01379 | 0.05401 | 0.80 |
| Place of Birth | -0.00452 | 0.01725 | 0.79 |
| Townsend Deprivation Index | -0.00001 | 0.00408 | 1.00 |
| Smoking - Former | 0.00658 | 0.01288 | 0.61 |
| Smoking - Never | -0.00092 | 0.01233 | 0.94 |
| Body Mass Index | 0.00201 | 0.00379 | 0.60 |

Probability scores were calculated using Bernoulli-mixtures applied to currently depressed individuals at Q2. All variables were fitted simultaneously. Sex, ethnicity (European ethnicity as the reference), place of birth, and smoking (current smoking as the reference) were fitted as factors. Bold values indicate P-values that were significant after Bonferroni correction.

Supplementary Table 164. Multivariable linear regression of individuals’ probability scores for Preserved Self-Worth and Appetite Disruption on health variables.

| Health variables | Effect Size | Standard Error | *P*-value |
| --- | --- | --- | --- |
| Myocardial infarction | -0.00103 | 0.01966 | 0.96 |
| Stroke | -0.01992 | 0.02184 | 0.36 |
| Asthma | -0.01105 | 0.00945 | 0.24 |
| COPD | -0.00658 | 0.01647 | 0.69 |
| Dementia | 0.05704 | 0.06220 | 0.36 |
| End stage renal disease | -0.04961 | 0.08172 | 0.54 |
| Parkinson’s disease | -0.05585 | 0.05160 | 0.28 |

Probability scores were calculated using Bernoulli-mixtures applied to currently depressed individuals at Q2. All variables were fitted simultaneously with age at questionnaire and sex fitted as covariates. Bold values indicate P-values that were significant after Bonferroni correction. COPD = chronic obstructive pulmonary disease.

Supplementary Table 165. Linear regressions of individuals’ probability scores for Preserved Self-Worth and Appetite Disruption on depression recurrence and on treatment resistant depression.

| Depression | Effect Size | Standard Error | *P*-value |
| --- | --- | --- | --- |
| Recurrent | -0.01973 | 0.01350 | 0.14 |
| Treatment resistance | -0.07388 | 0.02914 | 0.012 |

Recurrence and treatment resistance were both fitted as factors and were examined separately with age at questionnaire and sex fitted as covariates. Bold values indicate P-values that were significant after Bonferroni correction.

Supplementary Table 166. Linear regressions of individuals’ probability scores for Preserved Self-Worth and Appetite Disruption on polygenic scores for bipolar disorder, schizophrenia, and Attention-Deficit/Hyperactivity Disorder (ADHD).

| Mental health polygenic scores | Effect Size | Standard Error | *P*-value |
| --- | --- | --- | --- |
| Bipolar disorder | -0.00638 | 0.00409 | 0.12 |
| Schizophrenia | -0.00678 | 0.00397 | 0.09 |
| ADHD | -0.00077 | 0.00399 | 0.85 |

The polygenic scores for each trait were examined separately with age, sex, ancestry, and the first 10 genetic principal components fitted as covariates. Bold values indicate P-values that were significant after Bonferroni correction.

#### Preserved Self-Worth with SITB

Supplementary Table 167. Multivariable linear regression of individuals’ probability scores for Preserved Self-Worth with SITB on demographic variables.

| Demographic variables | Effect Size | Standard Error | *P*-value |
| --- | --- | --- | --- |
| Age | 0.00342 | 0.00065 | **1.47 × 10^-7^** |
| Sex | 0.01284 | 0.01032 | 0.21 |
| Ethnicity - Asian | 0.00382 | 0.04686 | 0.94 |
| Ethnicity - Black | -0.01551 | 0.05493 | 0.78 |
| Ethnicity - Chinese | -0.07154 | 0.12096 | 0.55 |
| Ethnicity - Mixed | 0.10275 | 0.05286 | 0.052 |
| Ethnicity - Other | -0.06388 | 0.06684 | 0.34 |
| Place of Birth | -0.00254 | 0.02134 | 0.91 |
| Townsend Deprivation Index | -0.01105 | 0.00505 | 0.029 |
| Smoking - Former | -0.03032 | 0.01594 | 0.06 |
| Smoking - Never | -0.01756 | 0.01526 | 0.25 |
| Body Mass Index | 0.00037 | 0.00469 | 0.94 |

Probability scores were calculated using Bernoulli-mixtures applied to currently depressed individuals at Q2. All variables were fitted simultaneously. Sex, ethnicity (European ethnicity as the reference), place of birth, and smoking (current smoking as the reference) were fitted as factors. Bold values indicate P-values that were significant after Bonferroni correction.

Supplementary Table 168. Multivariable linear regression of individuals’ probability scores for Preserved Self-Worth with SITB on health variables.

| Health variables | Effect Size | Standard Error | *P*-value |
| --- | --- | --- | --- |
| Myocardial infarction | 0.00585 | 0.02464 | 0.81 |
| Stroke | -0.03649 | 0.02737 | 0.18 |
| Asthma | 0.00507 | 0.01184 | 0.67 |
| COPD | -0.03023 | 0.02064 | 0.14 |
| Dementia | -0.06533 | 0.07796 | 0.40 |
| End stage renal disease | -0.09528 | 0.10243 | 0.35 |
| Parkinson’s disease | -0.10293 | 0.06468 | 0.11 |

Probability scores were calculated using Bernoulli-mixtures applied to currently depressed individuals at Q2. All variables were fitted simultaneously with age at questionnaire and sex fitted as covariates. Bold values indicate P-values that were significant after Bonferroni correction. COPD = chronic obstructive pulmonary disease.

Supplementary Table 169. Linear regressions of individuals’ probability scores for Preserved Self-Worth with SITB on depression recurrence and on treatment resistant depression.

| Depression | Effect Size | Standard Error | *P*-value |
| --- | --- | --- | --- |
| Recurrent | 0.03051 | 0.01827 | 0.10 |
| Treatment resistance | -0.07990 | 0.03374 | 0.018 |

Recurrence and treatment resistance were both fitted as factors and were examined separately with age at questionnaire and sex fitted as covariates. Bold values indicate P-values that were significant after Bonferroni correction.

Supplementary Table 170. Linear regressions of individuals’ probability scores for Preserved Self-Worth with SITB on polygenic scores for bipolar disorder, schizophrenia, and Attention-Deficit/Hyperactivity Disorder (ADHD).

| Mental health polygenic scores | Effect Size | Standard Error | *P*-value |
| --- | --- | --- | --- |
| Bipolar disorder | 0.00522 | 0.00507 | 0.30 |
| Schizophrenia | 0.00222 | 0.00491 | 0.65 |
| ADHD | 0.00006 | 0.00494 | 0.99 |

The polygenic scores for each trait were examined separately with age, sex, ancestry, and the first 10 genetic principal components fitted as covariates. Bold values indicate P-values that were significant after Bonferroni correction.

#### Low Self-Worth with SITB

Supplementary Table 171. Multivariable linear regression of individuals’ probability scores for Low Self-Worth with SITB on demographic variables.

| Demographic variables | Effect Size | Standard Error | *P*-value |
| --- | --- | --- | --- |
| Age | 0.00063 | 0.00053 | 0.24 |
| Sex | 0.02225 | 0.00843 | 0.0084 |
| Ethnicity - Asian | -0.03090 | 0.03829 | 0.42 |
| Ethnicity - Black | -0.07592 | 0.04488 | 0.09 |
| Ethnicity - Chinese | 0.16274 | 0.09884 | 0.10 |
| Ethnicity - Mixed | -0.05644 | 0.04319 | 0.19 |
| Ethnicity - Other | 0.03122 | 0.05461 | 0.57 |
| Place of Birth | 0.01885 | 0.01744 | 0.28 |
| Townsend Deprivation Index | 0.00156 | 0.00412 | 0.71 |
| Smoking - Former | 0.00013 | 0.01302 | 0.99 |
| Smoking - Never | -0.00223 | 0.01247 | 0.86 |
| Body Mass Index | -0.01023 | 0.00383 | 0.0076 |

Probability scores were calculated using Bernoulli-mixtures applied to currently depressed individuals at Q2. All variables were fitted simultaneously. Sex, ethnicity (European ethnicity as the reference), place of birth, and smoking (current smoking as the reference) were fitted as factors. Bold values indicate P-values that were significant after Bonferroni correction.

Supplementary Table 172. Multivariable linear regression of individuals’ probability scores for Low Self-Worth with SITB on health variables.

| Health variables | Effect Size | Standard Error | *P*-value |
| --- | --- | --- | --- |
| Myocardial infarction | -0.02477 | 0.02008 | 0.22 |
| Stroke | -0.02613 | 0.02231 | 0.24 |
| Asthma | -0.00936 | 0.00965 | 0.33 |
| COPD | -0.03528 | 0.01682 | 0.036 |
| Dementia | 0.06263 | 0.06354 | 0.32 |
| End stage renal disease | 0.01861 | 0.08347 | 0.82 |
| Parkinson’s disease | -0.02627 | 0.05271 | 0.62 |

Probability scores were calculated using Bernoulli-mixtures applied to currently depressed individuals at Q2. All variables were fitted simultaneously with age at questionnaire and sex fitted as covariates. Bold values indicate P-values that were significant after Bonferroni correction. COPD = chronic obstructive pulmonary disease.

Supplementary Table 173. Linear regressions of individuals’ probability scores for Low Self-Worth with SITB on depression recurrence and on treatment resistant depression.

| Depression | Effect Size | Standard Error | *P*-value |
| --- | --- | --- | --- |
| Recurrent | 0.03293 | 0.01376 | 0.017 |
| Treatment resistance | -0.01238 | 0.02635 | 0.64 |

Recurrence and treatment resistance were both fitted as factors and were examined separately with age at questionnaire and sex fitted as covariates. Bold values indicate P-values that were significant after Bonferroni correction.

Supplementary Table 174. Linear regressions of individuals’ probability scores for Low Self-Worth with SITB on polygenic scores for bipolar disorder, schizophrenia, and Attention-Deficit/Hyperactivity Disorder (ADHD).

| Mental health polygenic scores | Effect Size | Standard Error | *P*-value |
| --- | --- | --- | --- |
| Bipolar disorder | 0.00660 | 0.00415 | 0.11 |
| Schizophrenia | 0.00276 | 0.00402 | 0.49 |
| ADHD | -0.00388 | 0.00405 | 0.34 |

The polygenic scores for each trait were examined separately with age, sex, ancestry, and the first 10 genetic principal components fitted as covariates. Bold values indicate P-values that were significant after Bonferroni correction.

#### Low Self-Worth

Supplementary Table 175. Multivariable linear regression of individuals’ probability scores for Low Self-Worth on demographic variables.

| Demographic variables | Effect Size | Standard Error | *P*-value |
| --- | --- | --- | --- |
| Age | -0.00124 | 0.00087 | 0.16 |
| Sex | -0.03283 | 0.01384 | 0.018 |
| Ethnicity - Asian | -0.00923 | 0.06287 | 0.88 |
| Ethnicity - Black | 0.08243 | 0.07370 | 0.26 |
| Ethnicity - Chinese | 0.11911 | 0.16229 | 0.46 |
| Ethnicity - Mixed | -0.09504 | 0.07092 | 0.18 |
| Ethnicity - Other | -0.01484 | 0.08968 | 0.87 |
| Place of Birth | -0.02261 | 0.02864 | 0.43 |
| Townsend Deprivation Index | -0.02263 | 0.00677 | 8.41 × 10^-4^ |
| Smoking - Former | 0.02626 | 0.02138 | 0.22 |
| Smoking - Never | 0.02745 | 0.02047 | 0.18 |
| Body Mass Index | -0.01160 | 0.00629 | 0.07 |

Probability scores were calculated using Bernoulli-mixtures applied to currently depressed individuals at Q2. All variables were fitted simultaneously. Sex, ethnicity (European ethnicity as the reference), place of birth, and smoking (current smoking as the reference) were fitted as factors. Bold values indicate P-values that were significant after Bonferroni correction.

Supplementary Table 176. Multivariable linear regression of individuals’ probability scores for Low Self-Worth on health variables.

| Health variables | Effect Size | Standard Error | *P*-value |
| --- | --- | --- | --- |
| Myocardial infarction | -0.07465 | 0.03245 | 0.022 |
| Stroke | -0.04180 | 0.03606 | 0.25 |
| Asthma | -0.01365 | 0.01560 | 0.38 |
| COPD | -0.03057 | 0.02719 | 0.26 |
| Dementia | -0.21800 | 0.10270 | 0.034 |
| End stage renal disease | 0.27432 | 0.13493 | 0.042 |
| Parkinson’s disease | -0.17124 | 0.08520 | 0.045 |

Probability scores were calculated using Bernoulli-mixtures applied to currently depressed individuals at Q2. All variables were fitted simultaneously with age at questionnaire and sex fitted as covariates. Bold values indicate P-values that were significant after Bonferroni correction. COPD = chronic obstructive pulmonary disease.

Supplementary Table 177. Linear regressions of individuals’ probability scores for Low Self-Worth on depression recurrence and on treatment resistant depression.

| Depression | Effect Size | Standard Error | *P*-value |
| --- | --- | --- | --- |
| Recurrent | 0.02267 | 0.02231 | 0.31 |
| Treatment resistance | 0.04187 | 0.04090 | 0.31 |

Recurrence and treatment resistance were both fitted as factors and were examined separately with age at questionnaire and sex fitted as covariates. Bold values indicate P-values that were significant after Bonferroni correction.

Supplementary Table 178. Linear regressions of individuals’ probability scores for Low Self-Worth on polygenic scores for bipolar disorder, schizophrenia, and Attention-Deficit/Hyperactivity Disorder (ADHD).

| Mental health polygenic scores | Effect Size | Standard Error | *P*-value |
| --- | --- | --- | --- |
| Bipolar disorder | 0.00259 | 0.00675 | 0.70 |
| Schizophrenia | 0.00466 | 0.00654 | 0.48 |
| ADHD | -0.01504 | 0.00657 | 0.022 |

The polygenic scores for each trait were examined separately with age, sex, ancestry, and the first 10 genetic principal components fitted as covariates. Bold values indicate P-values that were significant after Bonferroni correction.

#### Psychomotor Changes and Preserved Self-Worth

Supplementary Table 179. Multivariable linear regression of individuals’ probability scores for Psychomotor Changes and Preserved Self-Worth on demographic variables.

| Demographic variables | Effect Size | Standard Error | *P*-value |
| --- | --- | --- | --- |
| Age | 0.00140 | 0.00047 | 0.0029 |
| Sex | 0.01757 | 0.00746 | 0.019 |
| Ethnicity - Asian | 0.09803 | 0.03388 | 0.0038 |
| Ethnicity - Black | 0.06831 | 0.03971 | 0.09 |
| Ethnicity - Chinese | -0.03211 | 0.08746 | 0.71 |
| Ethnicity - Mixed | 0.02623 | 0.03822 | 0.49 |
| Ethnicity - Other | 0.00044 | 0.04833 | 0.99 |
| Place of Birth | -0.02026 | 0.01543 | 0.19 |
| Townsend Deprivation Index | 0.00846 | 0.00365 | 0.020 |
| Smoking - Former | -0.00265 | 0.01152 | 0.82 |
| Smoking - Never | 0.00063 | 0.01103 | 0.95 |
| Body Mass Index | -0.00085 | 0.00339 | 0.80 |

Probability scores were calculated using Bernoulli-mixtures applied to currently depressed individuals at Q2. All variables were fitted simultaneously. Sex, ethnicity (European ethnicity as the reference), place of birth, and smoking (current smoking as the reference) were fitted as factors. Bold values indicate P-values that were significant after Bonferroni correction.

Supplementary Table 180. Multivariable linear regression of individuals’ probability scores for Psychomotor Changes and Preserved Self-Worth on health variables.

| Health variables | Effect Size | Standard Error | *P*-value |
| --- | --- | --- | --- |
| Myocardial infarction | 0.01147 | 0.01778 | 0.52 |
| Stroke | 0.07840 | 0.01975 | **7.38 × 10^-5^** |
| Asthma | 0.02285 | 0.00854 | 0.0075 |
| COPD | 0.00607 | 0.01490 | 0.68 |
| Dementia | 0.08402 | 0.05626 | 0.14 |
| End stage renal disease | -0.04977 | 0.07391 | 0.50 |
| Parkinson’s disease | 0.31020 | 0.04667 | **3.51 × 10^-11^** |

Probability scores were calculated using Bernoulli-mixtures applied to currently depressed individuals at Q2. All variables were fitted simultaneously with age at questionnaire and sex fitted as covariates. Bold values indicate P-values that were significant after Bonferroni correction. COPD = chronic obstructive pulmonary disease.

Supplementary Table 181. Linear regressions of individuals’ probability scores for Psychomotor Changes and Preserved Self-Worth on depression recurrence and on treatment resistant depression.

| Depression | Effect Size | Standard Error | *P*-value |
| --- | --- | --- | --- |
| Recurrent | 0.00232 | 0.01217 | 0.85 |
| Treatment resistance | 0.04308 | 0.02431 | 0.08 |

Recurrence and treatment resistance were both fitted as factors and were examined separately with age at questionnaire and sex fitted as covariates. Bold values indicate P-values that were significant after Bonferroni correction.

Supplementary Table 182. Linear regressions of individuals’ probability scores for Psychomotor Changes and Preserved Self-Worth on polygenic scores for bipolar disorder, schizophrenia, and Attention-Deficit/Hyperactivity Disorder (ADHD).

| Mental health polygenic scores | Effect Size | Standard Error | *P*-value |
| --- | --- | --- | --- |
| Bipolar disorder | -0.00622 | 0.00373 | 0.10 |
| Schizophrenia | 0.00746 | 0.00361 | 0.039 |
| ADHD | 0.00183 | 0.00364 | 0.61 |

The polygenic scores for each trait were examined separately with age, sex, ancestry, and the first 10 genetic principal components fitted as covariates. Bold values indicate P-values that were significant after Bonferroni correction.

#### Psychomotor Changes

Supplementary Table 183. Multivariable linear regression of individuals’ probability scores for Psychomotor Changes on demographic variables.

| Demographic variables | Effect Size | Standard Error | *P*-value |
| --- | --- | --- | --- |
| Age | 0.00085 | 0.00043 | 0.049 |
| Sex | 0.02384 | 0.00689 | 5.47 × 10^-4^ |
| Ethnicity - Asian | 0.04371 | 0.03129 | 0.16 |
| Ethnicity - Black | -0.03488 | 0.03667 | 0.34 |
| Ethnicity - Chinese | -0.06495 | 0.08076 | 0.42 |
| Ethnicity - Mixed | 0.06274 | 0.03529 | 0.08 |
| Ethnicity - Other | 0.00200 | 0.04462 | 0.96 |
| Place of Birth | 0.01879 | 0.01425 | 0.19 |
| Townsend Deprivation Index | 0.00716 | 0.00337 | 0.034 |
| Smoking - Former | -0.01151 | 0.01064 | 0.28 |
| Smoking - Never | -0.00319 | 0.01019 | 0.75 |
| Body Mass Index | -0.00248 | 0.00313 | 0.43 |

Probability scores were calculated using Bernoulli-mixtures applied to currently depressed individuals at Q2. All variables were fitted simultaneously. Sex, ethnicity (European ethnicity as the reference), place of birth, and smoking (current smoking as the reference) were fitted as factors. Bold values indicate P-values that were significant after Bonferroni correction.

Supplementary Table 184. Multivariable linear regression of individuals’ probability scores for Psychomotor Changes on health variables.

| Health variables | Effect Size | Standard Error | *P*-value |
| --- | --- | --- | --- |
| Myocardial infarction | 0.03988 | 0.01619 | 0.014 |
| Stroke | 0.02982 | 0.01799 | 0.10 |
| Asthma | -0.00434 | 0.00778 | 0.58 |
| COPD | 0.01676 | 0.01356 | 0.22 |
| Dementia | 0.15885 | 0.05122 | 0.0019 |
| End stage renal disease | 0.06631 | 0.06730 | 0.32 |
| Parkinson’s disease | 0.10347 | 0.04249 | 0.015 |

Probability scores were calculated using Bernoulli-mixtures applied to currently depressed individuals at Q2. All variables were fitted simultaneously with age at questionnaire and sex fitted as covariates. Bold values indicate P-values that were significant after Bonferroni correction. COPD = chronic obstructive pulmonary disease.

Supplementary Table 185. Linear regressions of individuals’ probability scores for Psychomotor Changes on depression recurrence and on treatment resistant depression.

| Depression | Effect Size | Standard Error | *P*-value |
| --- | --- | --- | --- |
| Recurrent | -0.00375 | 0.01049 | 0.72 |
| Treatment resistance | 0.00612 | 0.01934 | 0.75 |

Recurrence and treatment resistance were both fitted as factors and were examined separately with age at questionnaire and sex fitted as covariates. Bold values indicate P-values that were significant after Bonferroni correction.

Supplementary Table 186. Linear regressions of individuals’ probability scores for Psychomotor Changes on polygenic scores for bipolar disorder, schizophrenia, and Attention-Deficit/Hyperactivity Disorder (ADHD).

| Mental health polygenic scores | Effect Size | Standard Error | *P*-value |
| --- | --- | --- | --- |
| Bipolar disorder | -0.00279 | 0.00334 | 0.40 |
| Schizophrenia | 0.00082 | 0.00324 | 0.80 |
| ADHD | -0.00262 | 0.00326 | 0.42 |

The polygenic scores for each trait were examined separately with age, sex, ancestry, and the first 10 genetic principal components fitted as covariates. Bold values indicate P-values that were significant after Bonferroni correction.

#### Anhedonia and Low Self-Worth

Supplementary Table 187. Multivariable linear regression of individuals’ probability scores for Anhedonia and Low Self-Worth on demographic variables.

| Demographic variables | Effect Size | Standard Error | *P*-value |
| --- | --- | --- | --- |
| Age | 0.00009 | 0.00024 | 0.72 |
| Sex | -0.00235 | 0.00380 | 0.54 |
| Ethnicity - Asian | -0.01200 | 0.01726 | 0.49 |
| Ethnicity - Black | -0.01374 | 0.02023 | 0.50 |
| Ethnicity - Chinese | -0.00896 | 0.04455 | 0.84 |
| Ethnicity - Mixed | -0.01188 | 0.01947 | 0.54 |
| Ethnicity - Other | -0.01276 | 0.02462 | 0.60 |
| Place of Birth | 0.00177 | 0.00786 | 0.82 |
| Townsend Deprivation Index | 0.00083 | 0.00186 | 0.66 |
| Smoking - Former | -0.00170 | 0.00587 | 0.77 |
| Smoking - Never | -0.00435 | 0.00562 | 0.44 |
| Body Mass Index | 0.00125 | 0.00173 | 0.47 |

Probability scores were calculated using Bernoulli-mixtures applied to currently depressed individuals at Q2. All variables were fitted simultaneously. Sex, ethnicity (European ethnicity as the reference), place of birth, and smoking (current smoking as the reference) were fitted as factors. Bold values indicate P-values that were significant after Bonferroni correction.

Supplementary Table 188. Multivariable linear regression of individuals’ probability scores for Anhedonia and Low Self-Worth on health variables.

| Health variables | Effect Size | Standard Error | *P*-value |
| --- | --- | --- | --- |
| Myocardial infarction | 0.00257 | 0.00875 | 0.77 |
| Stroke | 0.01170 | 0.00972 | 0.23 |
| Asthma | -0.00018 | 0.00421 | 0.97 |
| COPD | 0.01115 | 0.00733 | 0.13 |
| Dementia | -0.01053 | 0.02769 | 0.70 |
| End stage renal disease | -0.01096 | 0.03638 | 0.76 |
| Parkinson’s disease | -0.00797 | 0.02297 | 0.73 |

Probability scores were calculated using Bernoulli-mixtures applied to currently depressed individuals at Q2. All variables were fitted simultaneously with age at questionnaire and sex fitted as covariates. Bold values indicate P-values that were significant after Bonferroni correction. COPD = chronic obstructive pulmonary disease.

Supplementary Table 189. Linear regressions of individuals’ probability scores for Anhedonia and Low Self-Worth on depression recurrence and on treatment resistant depression.

| Depression | Effect Size | Standard Error | *P*-value |
| --- | --- | --- | --- |
| Recurrent | -0.00501 | 0.00617 | 0.42 |
| Treatment resistance | 0.00854 | 0.00750 | 0.26 |

Recurrence and treatment resistance were both fitted as factors and were examined separately with age at questionnaire and sex fitted as covariates. Bold values indicate P-values that were significant after Bonferroni correction.

Supplementary Table 190. Linear regressions of individuals’ probability scores for Anhedonia and Low Self-Worth on polygenic scores for bipolar disorder, schizophrenia, and Attention-Deficit/Hyperactivity Disorder (ADHD).

| Mental health polygenic scores | Effect Size | Standard Error | *P*-value |
| --- | --- | --- | --- |
| Bipolar disorder | -0.00215 | 0.00182 | 0.24 |
| Schizophrenia | -0.00469 | 0.00177 | 0.0080 |
| ADHD | -0.00086 | 0.00178 | 0.63 |

The polygenic scores for each trait were examined separately with age, sex, ancestry, and the first 10 genetic principal components fitted as covariates. Bold values indicate P-values that were significant after Bonferroni correction.

#### General Anhedonia

Supplementary Table 191. Multivariable linear regression of individuals’ probability scores for General Anhedonia on demographic variables.

| Demographic variables | Effect Size | Standard Error | *P*-value |
| --- | --- | --- | --- |
| Age | -0.00163 | 0.00046 | 4.46 × 10^-4^ |
| Sex | 0.00235 | 0.00736 | 0.75 |
| Ethnicity - Asian | -0.04733 | 0.03345 | 0.16 |
| Ethnicity - Black | 0.02322 | 0.03921 | 0.55 |
| Ethnicity - Chinese | -0.04449 | 0.08634 | 0.61 |
| Ethnicity - Mixed | 0.01314 | 0.03773 | 0.73 |
| Ethnicity - Other | 0.00647 | 0.04771 | 0.89 |
| Place of Birth | 0.00245 | 0.01523 | 0.87 |
| Townsend Deprivation Index | 0.00355 | 0.00360 | 0.32 |
| Smoking - Former | -0.00088 | 0.01138 | 0.94 |
| Smoking - Never | 0.00030 | 0.01089 | 0.98 |
| Body Mass Index | 0.00605 | 0.00335 | 0.07 |

Probability scores were calculated using Bernoulli-mixtures applied to currently depressed individuals at Q2. All variables were fitted simultaneously. Sex, ethnicity (European ethnicity as the reference), place of birth, and smoking (current smoking as the reference) were fitted as factors. Bold values indicate P-values that were significant after Bonferroni correction.

Supplementary Table 192. Multivariable linear regression of individuals’ probability scores for General Anhedonia on health variables.

| Health variables | Effect Size | Standard Error | *P*-value |
| --- | --- | --- | --- |
| Myocardial infarction | 0.00618 | 0.01722 | 0.72 |
| Stroke | 0.00268 | 0.01913 | 0.89 |
| Asthma | 0.00396 | 0.00828 | 0.63 |
| COPD | 0.01107 | 0.01443 | 0.44 |
| Dementia | -0.04950 | 0.05449 | 0.36 |
| End stage renal disease | -0.03867 | 0.07159 | 0.59 |
| Parkinson’s disease | 0.03194 | 0.04520 | 0.48 |

Probability scores were calculated using Bernoulli-mixtures applied to currently depressed individuals at Q2. All variables were fitted simultaneously with age at questionnaire and sex fitted as covariates. Bold values indicate P-values that were significant after Bonferroni correction. COPD = chronic obstructive pulmonary disease.

Supplementary Table 193. Linear regressions of individuals’ probability scores for General Anhedonia on depression recurrence and on treatment resistant depression.

| Depression | Effect Size | Standard Error | *P*-value |
| --- | --- | --- | --- |
| Recurrent | -0.01584 | 0.01256 | 0.21 |
| Treatment resistance | -0.02543 | 0.02119 | 0.23 |

Recurrence and treatment resistance were both fitted as factors and were examined separately with age at questionnaire and sex fitted as covariates. Bold values indicate P-values that were significant after Bonferroni correction.

Supplementary Table 194. Linear regressions of individuals’ probability scores for General Anhedonia on polygenic scores for bipolar disorder, schizophrenia, and Attention-Deficit/Hyperactivity Disorder (ADHD).

| Mental health polygenic scores | Effect Size | Standard Error | *P*-value |
| --- | --- | --- | --- |
| Bipolar disorder | -0.00359 | 0.00354 | 0.31 |
| Schizophrenia | -0.00696 | 0.00343 | 0.042 |
| ADHD | 0.00378 | 0.00345 | 0.27 |

The polygenic scores for each trait were examined separately with age, sex, ancestry, and the first 10 genetic principal components fitted as covariates. Bold values indicate P-values that were significant after Bonferroni correction.

#### All Symptoms

Supplementary Table 195. Multivariable linear regression of individuals’ probability scores for All Symptoms on demographic variables.

| Demographic variables | Effect Size | Standard Error | *P*-value |
| --- | --- | --- | --- |
| Age | -0.00420 | 0.00068 | **6.78 × 10^-10^** |
| Sex | -0.01722 | 0.01076 | 0.11 |
| Ethnicity - Asian | 0.02307 | 0.04887 | 0.64 |
| Ethnicity - Black | -0.05619 | 0.05728 | 0.33 |
| Ethnicity - Chinese | 0.02648 | 0.12615 | 0.83 |
| Ethnicity - Mixed | 0.02175 | 0.05513 | 0.69 |
| Ethnicity - Other | 0.11788 | 0.06971 | 0.09 |
| Place of Birth | -0.01336 | 0.02226 | 0.55 |
| Townsend Deprivation Index | 0.01116 | 0.00526 | 0.034 |
| Smoking - Former | -0.02300 | 0.01662 | 0.17 |
| Smoking - Never | -0.02398 | 0.01591 | 0.13 |
| Body Mass Index | 0.01360 | 0.00489 | 0.0054 |

Probability scores were calculated using Bernoulli-mixtures applied to currently depressed individuals at Q2. All variables were fitted simultaneously. Sex, ethnicity (European ethnicity as the reference), place of birth, and smoking (current smoking as the reference) were fitted as factors. Bold values indicate P-values that were significant after Bonferroni correction.

Supplementary Table 196. Multivariable linear regression of individuals’ probability scores for All Symptoms on health variables.

| Health variables | Effect Size | Standard Error | *P*-value |
| --- | --- | --- | --- |
| Myocardial infarction | 0.05103 | 0.02523 | 0.043 |
| Stroke | 0.06619 | 0.02804 | 0.018 |
| Asthma | 0.00303 | 0.01213 | 0.80 |
| COPD | 0.06866 | 0.02114 | 0.0012 |
| Dementia | 0.04970 | 0.07985 | 0.53 |
| End stage renal disease | -0.10981 | 0.10490 | 0.30 |
| Parkinson’s disease | -0.00088 | 0.06624 | 0.99 |

Probability scores were calculated using Bernoulli-mixtures applied to currently depressed individuals at Q2. All variables were fitted simultaneously with age at questionnaire and sex fitted as covariates. Bold values indicate P-values that were significant after Bonferroni correction. COPD = chronic obstructive pulmonary disease.

Supplementary Table 197. Linear regressions of individuals’ probability scores for All Symptoms on depression recurrence and on treatment resistant depression.

| Depression | Effect Size | Standard Error | *P*-value |
| --- | --- | --- | --- |
| Recurrent | 0.01533 | 0.01507 | 0.31 |
| Treatment resistance | 0.09690 | 0.03394 | 0.0045 |

Recurrence and treatment resistance were both fitted as factors and were examined separately with age at questionnaire and sex fitted as covariates. Bold values indicate P-values that were significant after Bonferroni correction.

Supplementary Table 198. Linear regressions of individuals’ probability scores for All Symptoms on polygenic scores for bipolar disorder, schizophrenia, and Attention-Deficit/Hyperactivity Disorder (ADHD).

| Mental health polygenic scores | Effect Size | Standard Error | *P*-value |
| --- | --- | --- | --- |
| Bipolar disorder | 0.00798 | 0.00525 | 0.13 |
| Schizophrenia | 0.00905 | 0.00509 | 0.08 |
| ADHD | 0.01298 | 0.00511 | 0.011 |

The polygenic scores for each trait were examined separately with age, sex, ancestry, and the first 10 genetic principal components fitted as covariates. Bold values indicate P-values that were significant after Bonferroni correction.
